# A continental-scale scenario modelling framework for evaluating infant RSV immunisation strategies across Europe

**DOI:** 10.64898/2026.06.10.26355338

**Authors:** Elisa Viola, Mattia Mazzoli, Daniela Paolotti, Alessandro Rizzo, Lorenzo Zino, Nicolò Gozzi

## Abstract

**Background:** The recent approval of long-acting monoclonal antibodies (la-mAbs) and a maternal vaccine (MV) in the EU enables universal RSV prevention in infants. Modelling studies are widely used to quantify the population-level impact of alternative immunisation strategies. However, existing assessments of new RSV immunisation products focus on national or sub-national settings.

**Methods:** We developed an age-stratified, stochastic compartmental model of RSV transmission for 28 EU/EEA countries. It combines literature-based parameters on RSV natural history and product efficacy with country-specific demographic and contact patterns. After model calibration against age- and country-specific RSV hospitalisation rates, we designed scenarios for both la-mAbs and MV at four coverage levels, with and without catch-up immunisation for infants under six months at season onset. We then evaluated each scenario against a no-immunisation baseline.

**Results:** At 95% coverage, the cross-country median reduction in RSV hospitalisations over one season in infants under 12 months is 29.9% for la-mAbs (country median range: 27.7 *−* 33.9%) and 22.4% for MV (20.0 *−* 25.6%), scaling linearly with coverage. Out of all averted hospitalisations, 78.3% (90% CI: [67.3, 92.7]%) are concentrated in infants aged 0*−*2 months for la-mAbs and 72.7% (90% CI: [61.4, 88.6]%) for MV. A catch-up campaign nearly doubles the overall reduction in RSV hospitalisations.

**Conclusions:** Despite country-specific heterogeneities, impact of la-mAbs and MV is comparable across settings and herd-immunity effects are largely negligible. This supports harmonised European guidelines on coverage targets. Seasonal catch-up campaigns emerge as an effective lever to maximise the impact of immunisation programmes.

**Funding:** This publication is part of the project PNRR-NGEU which has received funding from the MUR-DM 630/2024.

## Introduction

Respiratory Syncytial Virus (RSV) is the leading pathogen responsible for acute lower respiratory infections worldwide, with the greatest burden falling on young children ^1^. Globally, an estimated 33 million RSV-associated acute lower respiratory infection episodes and 3.6 million hospitalisations occurred in children under five years of age in 2019 ^1^. In the European Union (EU), an estimated 245,000 RSV-associated hospitalisations occur annually among children under five years of age, with 75% of the burden affecting infants in their first year of life ^2^. The highest hospitalisation rate is observed for infants aged 0 *−*2 months, reaching 71.6 per 1,000 children ^2^. This substantial hospitalisation burden generates significant direct healthcare costs, making RSV prevention a critical public health priority. Until recently, the only available immunoprophylaxis against RSV in infants was palivizumab (Synagis^®^), a monoclonal antibody requiring multiple injections per season due to the short duration of protection, and as such was restricted to use in high-risk infants ^3,4^. The recent approval of new immunisation products by European authorities presents a valuable opportunity to advance RSV prevention through universal infant immunisation strategies. Nirsevimab (Beyfortus^®^) is a long-acting monoclonal antibody authorised in the EU in October 2022 ^5^. It provides protection against severe RSV up to six months with a single dose and is indicated for all newborns and infants during their first RSV season, as well as for children up to 24 months who remain vulnerable through their second season ^4^. In addition, a maternal RSV vaccine (Abrysvo^®^), authorised in August 2023, confers passive protection to infants from birth up to six months of age following a single maternal dose administered between weeks 24 and 36 of gestation ^4,6^. These products enabled for the first time universal prevention strategies against RSV-associated hospitalisations in infants, extending prophylaxis beyond the high-risk newborns previously covered by palivizumab.

Following these regulatory approvals, several European countries have rapidly moved to incorporate these new prevention tools into national immunisation programmes. As of the 2025*/*26 season, 16 EU/EEA countries have established fully funded universal infant immunisation programmes with long-acting monoclonal antibodies, offering them seasonally to all newborns ^4^. In parallel, 9 countries have introduced fully funded maternal RSV vaccination programmes, typically administered seasonally between September and March ^4^. Only 5 countries (Belgium, Cyprus, France, Greece, and Luxembourg) offer both strategies, allowing pregnant individuals to choose between maternal vaccination and infant monoclonal antibody administration at birth ^4^. As the decision-making on the integration of such products into national strategies evolve and European-level guidance is under development ^4,7^, epidemic modelling can play a key role in supporting these processes.

Mathematical models of infectious diseases are widely used to study epidemic dynamics and evaluate the potential impact of public health interventions. Within this landscape, scenario modelling has emerged as a valuable methodological framework, enabling the systematic comparison of multiple plausible futures under alternative assumptions about interventions and epidemiological uncertainties ^8^. In recent years, RSV has been the subject of a rich modelling literature, with dynamic transmission models that capture its key epidemiological features ^9^. Common modelling choices include the representation of maternally derived immunity in newborns and naturally acquired immunity from prior infection ^10–12^, and age-stratified population structure which allows for a more granular assessment of disease burden across the most vulnerable ^11–13^. A substantial body of this work has focused on the population-level impact of pharmaceutical interventions such as maternal vaccination and monoclonal antibodies, but these efforts have so far been confined to individual national contexts, such as France ^12^, Germany ^14^, England and Wales ^15^, or to subnational settings such as the Veneto region in Italy ^13^. A broader, European-level evaluation of these immunisation products at the population level is still missing, yet such evidence is critical to support countries in designing and calibrating their immunisation programmes on a comparable analytical basis.

This study addresses this gap presenting a data-driven, stochastic compartmental epidemic model calibrated across 28 EU/EEA countries (the 27 EU Member States and Norway) to evaluate RSV immunisation strategies at the continental level through systematic scenario modelling. In Europe, scenario modeling efforts are coordinated by the European Centre for Disease Prevention and Control (ECDC) through RespiCompass, a collaborative scenario modelling hub which brings together several modelling groups to evaluate the population-level impact of respiratory viruses at the European level ^16^. RespiCompass most recent round was dedicated to assessing the impact of new RSV immunisation products in infants, reflecting the priority of RSV prevention in European public health agendas. We leverage the harmonised burden estimates produced within this modeling exercise and we applya single coherent transmission model to all 28 countries under multiple scenarios. The model incorporates country-specific demographic structures, births, and social contact patterns ^17,18^, enabling a comparative analysis that accounts for the heterogeneity of European populations. We show that both long-acting monoclonal antibodies and maternal vaccination can substantially reduce infant hospitalisations and that seasonal immunisation catch-up campaigns for infants under six months can nearly double the hospitalisations averted. While the relative effectiveness of these strategies is broadly consistent across countries, the doses required to avert one hospitalisation show larger variation across countries, reflecting the differences in background RSV hospital burden. As European countries are actively introducing and refining universal RSV immunisation programmes, these results provide timely evidence on which elements of programme design can be meaningfully harmonised through European guidance.

## Methods

### Model description

The MSIRS_3_ model provides a detailed representation of RSV transmission dynamics that accounts for country-specific demographic characteristics and key epidemiological features of respiratory syncytial virus. The model acronym MSIRS_3_ denotes a Susceptible-Infected-Removed-Susceptible (SIRS) framework that incorporates maternal immunity (M) and three distinct levels of acquired immunity after successive infections ^9,12^. We subdivide each compartment X *∈*{M, S, I, R} into X_i,j,k_, where *i* denotes the age group, *j* the immunity level, and *k* the treatment status. The baseline system (*k* = 0) represents individuals without intervention (Figure 1a).

**Figure 1:**
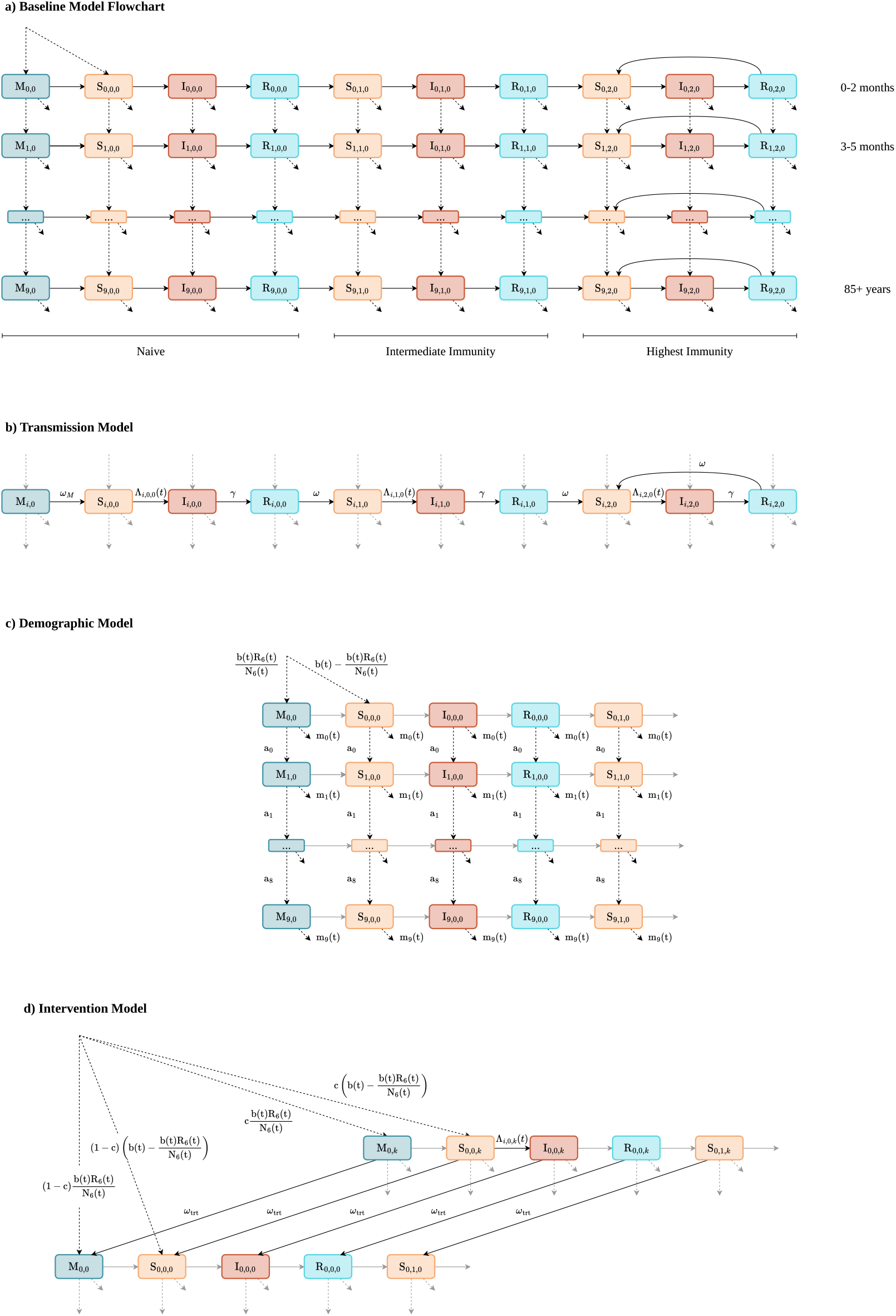
Schematic representation of the transmission model. Black arrows highlight the processes emphasised in each panel, grey arrows indicate the remaining model interactions shown for completeness. Solid lines denote transitions related to RSV transmission and immune dynamics, dashed lines represent changes driven by demographic processes. **(a)** Overview of the baseline model structure. **(b)** Detail of the RSV transmission dynamics. **(c)** Detail of the demographic processes. **(d)** Coupling between the baseline transmission system and the additional compartmental structure representing the immunised population.

Newborns enter either the maternally immune compartment M or the susceptible compartment S, depending on the immune status of the mother. Maternally protected infants lose their passive immunity at rate *ω*_*M*_ and transition to S. Susceptible individuals become infected according to the force of infection Λ_*i,j,k*_(*t*) (Supplementary Section S1.1) and move to the infected compartment I, where they remain infectious until recovering at rate *γ* into the removed compartment R. After recovery, individuals transition from *R*_*i,j,k*_ to *S*_*i,j*+1,*k*_ at rate *ω*_*R*_, with reduced susceptibility quantified by a multiplicative factor *λ*_*j*_ (*j* = 0, 1, 2), where *λ*_0_ *> λ*_1_ *> λ*_2_ and maximum immunity (*λ*_2_) is reached from the third infection onward (Figure 1b).

We compute hospitalisations as a fraction of the daily flow from I to R, using age-specific hospitalisation probabilities that reflect the age-dependent severity of RSV disease.

### Demographic structure

We stratified the population into ten age groups with enhanced resolution for the most vulnerable ages: 0–2 months, 3–5 months, 6–11 months, 1–2 years, 3–4 years, 5–17 years, 18–64 years, 65–74 years, 75–84 years, and 85+ years. Individuals age out of each group *i* at rate *a*_*i*_, defined as the inverse of the time spent in group *i*. All individuals, regardless of compartment and age group, are subject to age-specific mortality not linked to RSV disease. Newborns enter the system according to the time-varying birth rate *b*(*t*); we assign a fraction proportional to the share of recovered individuals in the fertile age group (18–64 years) to the maternally immune compartment M, while the remainder enters S directly (Figure 1c).

### Model implementation

We implemented the model as a chain binomial process. Disease-related transitions between compartments are stochastic: at each time step, the number of individuals moving from compartment *X* to compartment *Y* is sampled from a binomial distribution Bin(*X*_*i,j,k*_(*t*), *r*_*XY*_ Δ*t*), where *r*_*XY*_ is the corresponding transition rate.

### Model inputs

For each country, we initialise the model with their specific demographic and contact data. We draw population counts by age from the official Eurostat statistics ^19^. We obtain monthly birth and death rates from Eurostat demographic series ^20,21^; age-stratified mortality is only available as an annualdistribution, we therefore redistribute it proportionally across monthly death totals to derive daily mortality counts. Country-specific contact matrices inform social mixing patterns across age groups. For the majority of countries, we use the synthetic matrices from Ref. ^17^, which provide contact rates at one-year age resolution. For Belgium, Croatia, Malta, and Poland, where these matrices are not available, we rely on the matrices from Ref. ^18^, which offer a coarser five-year age resolution. Where model age groups are finer than the matrix resolution, we assume that all sub-groups within a given age bracket share the same contact behaviour.

### Model calibration

#### Parameters

We classified model parameters as either fixed across countries or calibrated independently for each country (Table 1). Fixed parameters, governing the duration of infectiousness, natural and maternal immunity, and the reduction in susceptibility after successive infections, were drawn from published estimates. Calibrated parameters comprised the three components of the seasonal transmission rate *β*(*t*) — baseline amplitude *β*_min_, peak amplitude *β*_max_, and peak timing *t*_peak_ — together with ten age-specific probabilities of hospitalisation given infection *ϕ*_*i*_. We informed the prior on *ϕ*_*i*_ with literature evidence indicating that severe RSV disease risk is highest in neonates, declines through the first years of life ^22^, and rises again after age 65 ^23^. Detailed prior specifications are provided in Table 1 and Supplementary Section S2.1.

**Table 1:** Model parameters. Fixed parameters (top) are shared across all countries. Calibrated parameters (bottom) are estimated independently for each country; prior ranges are reported. The subscript *i* indexes the ten model age groups: 0–2 m, 3–5 m, 6–11 m, 1–2 y, 3–4 y, 5–17 y, 18–64 y, 65–74 y, 75–84 y, and 85+.

| Parameter | Symbol | Value / Prior | Source |
| --- | --- | --- | --- |
| *Fixed parameters* |  |  |  |
| Duration of infectiousness | $1/\gamma$ | 7 days | <sup>12</sup> |
| Waning of natural immunity | $1/\omega$ | 9 months | <sup>11,12</sup> |
| Waning of maternal immunity | $1/\omega_M$ | 3 months | <sup>12</sup> |
| Susceptibility reduction<br>(1st infection) | $\lambda_1$ | $7.6 \times 10^{-1}$ | <sup>12</sup> |
| Susceptibility reduction<br>(2nd infection) | $\lambda_2$ | $8.8 \times 10^{-1}$ | <sup>12</sup> |
| *Calibrated parameters* |  |  |  |
| Seasonal transmission rate | $(\beta_{\min}, \beta_{\max})$ | Uniform($\mathcal{D}$)<br>$\mathcal{D} \subseteq \{(\beta_{\min}, \beta_{\max}) : 4.9 \times 10^{-3} \leq \beta_{\min} \leq 3.5 \times 10^{-2}, \beta_{\min} + 1.7 \times 10^{-3} \leq \beta_{\max} \leq 5.0 \times 10^{-2}\}$ | <sup>12</sup> |
| Peak timing | $t_{\text{peak}}$ | Uniform over 1 Sep–1 Mar<br>$\cup \{244, \dots, 365\}$ | |
| Hospitalisation probability<br>after infection (0–2 m) | $\phi_{0-2\text{m}}$ | Uniform($10^{-4}, 10^{-1}$) | |
| Hospitalisation probability<br>after infection (3 m–64 y) | $\phi_i$ | Uniform($10^{-4}, \phi_{i-1}$) | <sup>22</sup> |
| Hospitalisation probability<br>after infection (65 y+) | $\phi_i$ | Uniform($\phi_{i-1}, \phi_{0-2\text{m}}$) | <sup>23</sup> |

#### Calibration data

We calibrated the model against RSV-associated hospitalisation data provided by the ECDC Respi-Compass collaborative modelling hub for the 2025–2026 round ^16^, covering all 28 European countries included in our analysis. These data represent a realistic epidemic season ^24^ spanning from 1 September to 31 May, generated by recombining age-specific hospitalisation burden estimates for infants ^2^ and the elderly ^25^ into a single seasonal trajectory. The resulting datasets provide two calibration targets: the cumulative RSV hospitalisation burden by age group and the weekly hospitalisation curve over the season. They follow a coarser age stratification than the model (0–2 months, 3–5 months, 6–11 months, 1–4 years, 5–64 years, and 65+ years).

#### Calibration algorithm

We calibrated the model to each country using an Approximate Bayesian Computation (ABC) approach with a simulation-budget strategy ^26,27^. To enable efficient sampling of initial conditions at equilibrium, we drew each simulation’s starting state from a precomputed set of long-run model outputs indexed by transmission parameters (Supplementary Section S2.2 details the sampling strategy and filtering criteria). We then sampled the calibrated parameters from the priors defined in Table 1, producing 100,000 simulations per country. For each simulation, we computed the root mean square error against the weekly hospitalisation curve and the mean absolute percentage error against the cumulative age-stratified burden. We combined the two rankings via a Borda score and retained the 300 best-performing simulations (Supplementary Section S2.3 for the metric definitions and full ranking procedure).

#### Scenario definition

We consider a set of scenarios to enable a systematic comparison of the two immunisation strategies in Europe. In this context, a scenario is a conditional projection that describes what would be expected to happen if a specific set of conditions were met. Two types of factors guide the construction of scenarios: interventions, which represent decision options under the control of policy makers, and uncertainties, which capture key unknowns that may affect outcomes but lie beyond direct control ^8^. Figure 2 shows a summary of all scenarios evaluated in this study. The rows of the table explore the intervention dimension.

**Figure 2:**
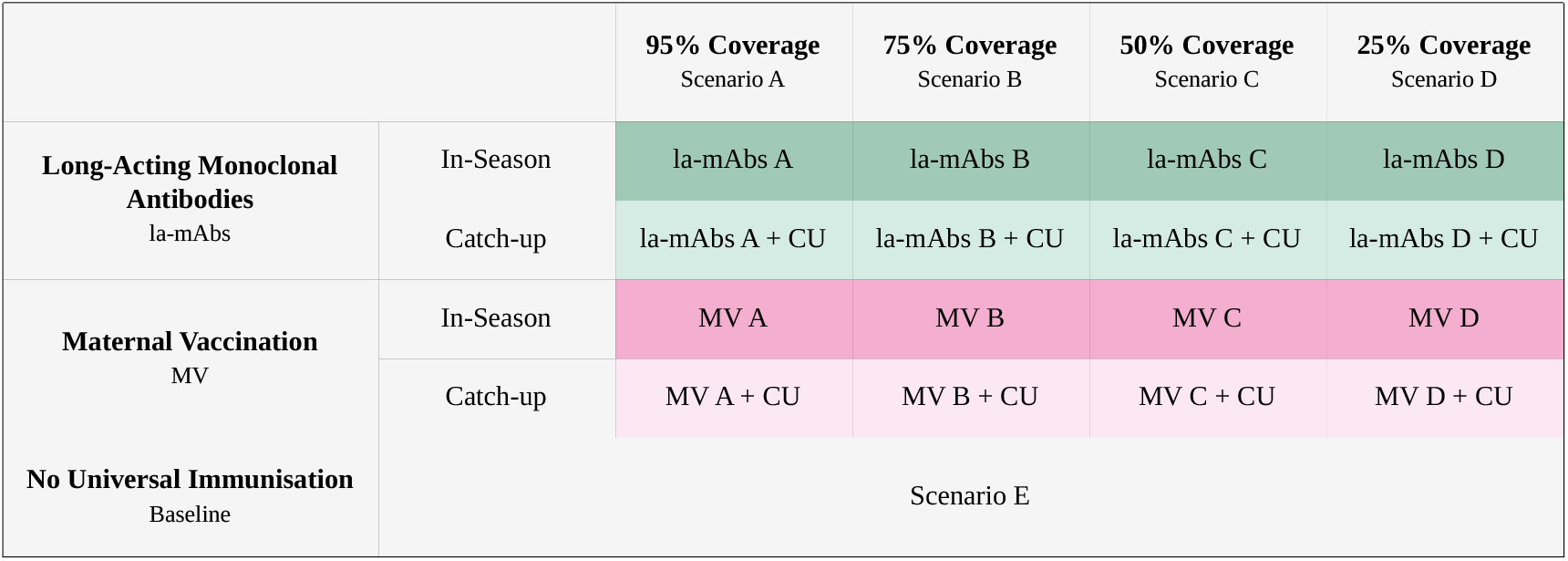
Summary of immunisation scenarios explored. La-mAbs refers to long-acting monoclonal antibodies administered to newborns at birth during the RSV season. MV refers to maternal vaccination administered during antenatal care visits between 32 and 36 weeks of gestation, timed so that immunised infants are born within the same seasonal window. “In-season” scenarios assume immunisation of newborns only; “+ Catch-up” scenarios additionally include la-mAb administration at season onset to infants born up to six months. Four coverage levels are evaluated: 95% (A), 75% (B), 50% (C), and 25% (D). Scenario E represents the baseline with no universal RSV immunisation programme. Equal coverage is assumed for at birth and catch-up cohorts.

Specifically, the rows contrast the two immunisation strategies, each further differentiated by their immunisation target, resulting in four intervention configurations. Two of them represent in-season immunisation only, through long-acting monoclonal antibodies (Intervention la-mAbs) or maternal vaccination (Intervention MV). The other two add a catch-up component (Intervention la-mAbs+CU and Intervention MV+CU, respectively), in which infants born up to six months before the season start also receive immunisation at season onset, corresponding to the first day of simulation (1 September) ^4^. Catch-up doses are administered as monoclonal antibodies, regardless of the standard strategy, reflecting the fact that only monoclonal antibodies can be administered after birth. Early data from Spain indicate that catch-up uptake is comparable to birth-dose uptake ^28,29^; we therefore assumed equal coverage for catch-up and in-season administrations. The columns capture the uncertainty dimension, represented here by the level of vaccination coverage achieved during the campaign. To capture a wide range of possible uptake, we evaluated four coverage levels: 25% (Scenario A), 50% (Scenario B), 75% (Scenario C), and 95% (Scenario D) for each intervention type. Regional assessments from Spain’s 2024 campaign and Italy’s 2025 campaign provide early evidence that la-mAb uptake at birth as high as 95% is achievable in practice ^28–30^. We compare all intervention scenarios against a baseline (Scenario E) representing the current status quo, in which neither strategy is universally implemented. Where applicable, immunisation of high-risk newborns is assumed to continue according to existing country practice.

We aligned intervention timing with current national programmes and clinical recommendations.

For long-acting monoclonal antibodies, we assume administration at birth during the RSV season, from September 1^*st*^ to March 31^*st*^. For maternal vaccination, we assume administration between 32 and 36 weeks of gestation during antenatal care visits, timed so that all immunised infants are born within the same seasonal window as for la-mAbs. These assumptions reflect both existing national implementation strategies and manufacturer recommendations ^4,7^.

### Modelling interventions

For both interventions, the constant *c* determines the proportion of newborns entering the treated system, which mirrors the baseline model structure (Figure 1d). For maternal vaccination, we additionally model protection in vaccinated mothers, who enter the treated compartmental layer with the same vaccine effectiveness as their newborns.

In our model, both interventions provide dual protection by reducing the risk of infection and the probability of hospitalisation given breakthrough infection. We parameterise effectiveness using clinical trial estimates (Table 2). For each simulation, we sample vaccine effectiveness against hospitalisation from a Beta distribution fitted to the reported 95% confidence interval and derive effectiveness against infection assuming a fixed ratio between the two endpoints (Supplementary Section S1.2). Effectiveness against infection modifies the force of infection in the treated system, while the conditional effectiveness against hospitalisation given infection reduces the age-specific probability of hospitalisation in treated individuals who experience breakthrough infection.

**Table 2:** Intervention effectiveness parameters. Literature-based vaccine efficacy (VE) and duration of protection for the two RSV immunisation products considered in the scenarios: long-acting monoclonal antibodies (la-mAbs) administered at birth and maternal vaccination (MV) administered to pregnant women. VE against hospitalisation is reported as the median with 95% confidence interval.

| Intervention | VE hospitalisation<br>(median %, 95% CI) | VE infection<br>(median%) | Duration<br>(months) |
| --- | --- | --- | --- |
| Monoclonal antibodies | 87 [81 – 91]<sup>31</sup> | 73<sup>31</sup> | 6<sup>15,31</sup> |
| Maternal vaccination | 71 [53 – 82]<sup>32</sup> | 57<sup>33</sup> | 6<sup>15</sup> |

Protection from both interventions wanes over an average period of 1*/ω*_*trt*_. We model this waning through a three-stage Erlang distribution ^15^. As immunity declines, individuals transition from the treated compartments back to the baseline system (*k* = 0).

### Sensitivity analysis

We assessed the sensitivity of our results to the modelling of waning protection using Italy as an illustrative case study. We re-ran all intervention scenarios under three alternative specifications of the waning process: no waning, exponential decay, and the three-stage Erlang distribution adopted in the main analysis. We then compared scenario-level outcomes across the three specifications to quantify the influence of this modelling choice on the conclusions.

### Scenario evaluation

The 300 calibrated simulations retained from the calibration procedure correspond to the baseline scenario (Scenario E), in which no immunisation programme is in place. To generate projections under each intervention scenario, we re-run the model using exactly the same parameter sets and stochastic seeds, modifying only the immunisation strategy and coverage level.

For each intervention scenario *S*, we computed the number of *averted hospitalisations* as the paired simulation-level difference from the baseline, and the corresponding *percentage reduction* over the first year of life as our primary measure of intervention impact. To characterise the distribution of benefits across age groups at the European level, we computed the *age-group share* of averted hospitalisations, defined as the fraction attributable to each age group after aggregating across all 28 countries at the simulation level. Finally, to evaluate programmatic efficiency, we computed the *doses per averted hospitalisation (DPA)* as the ratio between the total doses administered and the number of averted hospitalisations in the first year of life. All quantities are summarised at the country level as the median across the retained simulations, with 90% credible intervals of the corresponding distributions. Formal definitions are provided in Supplementary Section S3.

## Results

### European heterogeneities in demographic and RSV hospitalisation burden

In European countries, the population of up to one year at the beginning of the considered season ranged from 0.7% of the total population in Italy to 1.1% in Cyprus. Similarly, births that occurred between September and May ranged from 0.5% of the total population at the start of the season in Italy to 0.8% in Ireland and Cyprus (Figure 3, top).

**Figure 3:**
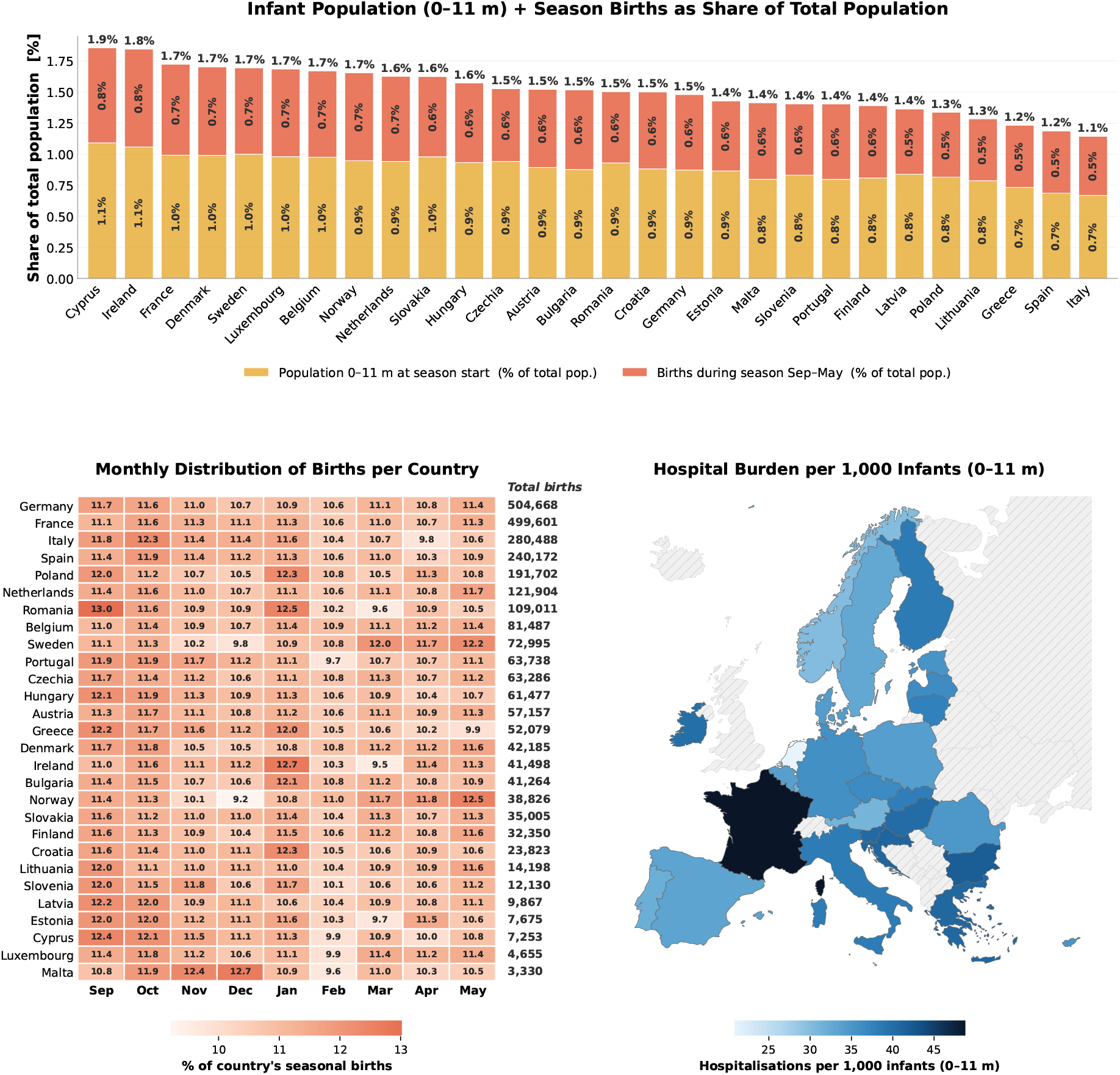
Demographic and clinical burden of RSV-relevant infant populations across 28 European countries over a realistic respiratory season (September-May). **(Top)** Stacked bar chart showing, for each country, the size of the at-risk infant population as a share of total population: the mustard segment represents infants aged 0 *−* 11 months alive at the season start, and the coral segment represents additional infants born during the season; the value on top of each bar reports the combined share. Countries are ranked by combined share in decreasing order. **(Bottom left)** Heatmap of the monthly distribution of births within the season for each country, expressed as a percentage of that country’s total seasonal births (rows sum to 100%); the absolute number of births during the season is reported on the right of each row. Countries are ranked by total seasonal births in decreasing order. **(Bottom right)** Choropleth map showing the observed hospitalisation burden among infants aged 0 *−* 11 months, expressed as hospitalisations per 1,000 infants over the season. Countries shown with diagonal hatching are those for which hospitalisation data were not available.

The bottom-left panel of Figure 3 shows that the seasonality of births varied across countries. The peak birth quarter was March–May in 3 of the 28 countries (Netherlands, Sweden, Norway), and December–February in 5 countries (Malta, Poland, Ireland, Bulgaria, and Croatia), while the majority of the countries reported a birth peak in the first quarter of the season, between September and November.

The epidemiological burden data on RSV used in this study, summarised in the bottom-right panel of Figure 3, highlighted that in the age group of 0–11 months, the median number of RSV-related hospitalisations across the 28 European countries was 35.6 per 1,000 infants, ranging from 20.8 hospitalisations per 1,000 in the Netherlands to a peak of 48.8 per 1,000 in France, with five additional countries (Hungary, Slovenia, Greece, Croatia, and Bulgaria) reporting more than 40 cases per 1,000 over the course of the season.

### Model fit and estimated parameters

In this section, we report calibration results for two countries with contrasting demographic and epidemiological profiles, France and Sweden. Complete calibration outputs for all 28 countries are provided in Supplementary Section S4.

For both countries, the baseline scenario reproduces well the dual calibration targets (Figure 4). For the weekly hospitalisation curve, the 50% credible interval of the simulated distributions captured 30.8% of the observed data points in both countries, while the 90% interval captured 100.0% for France and 94.9% for Sweden. For the cumulative age-stratified burden, the 50% and 90% intervals captured 83.3% and 100.0% of the data for France, and 100.0% at both levels for Sweden. A pointwise comparison between observed and simulated values is provided in Supplementary Tables S1-S2 together with equivalent performance metrics for the remaining countries.

**Figure 4:**
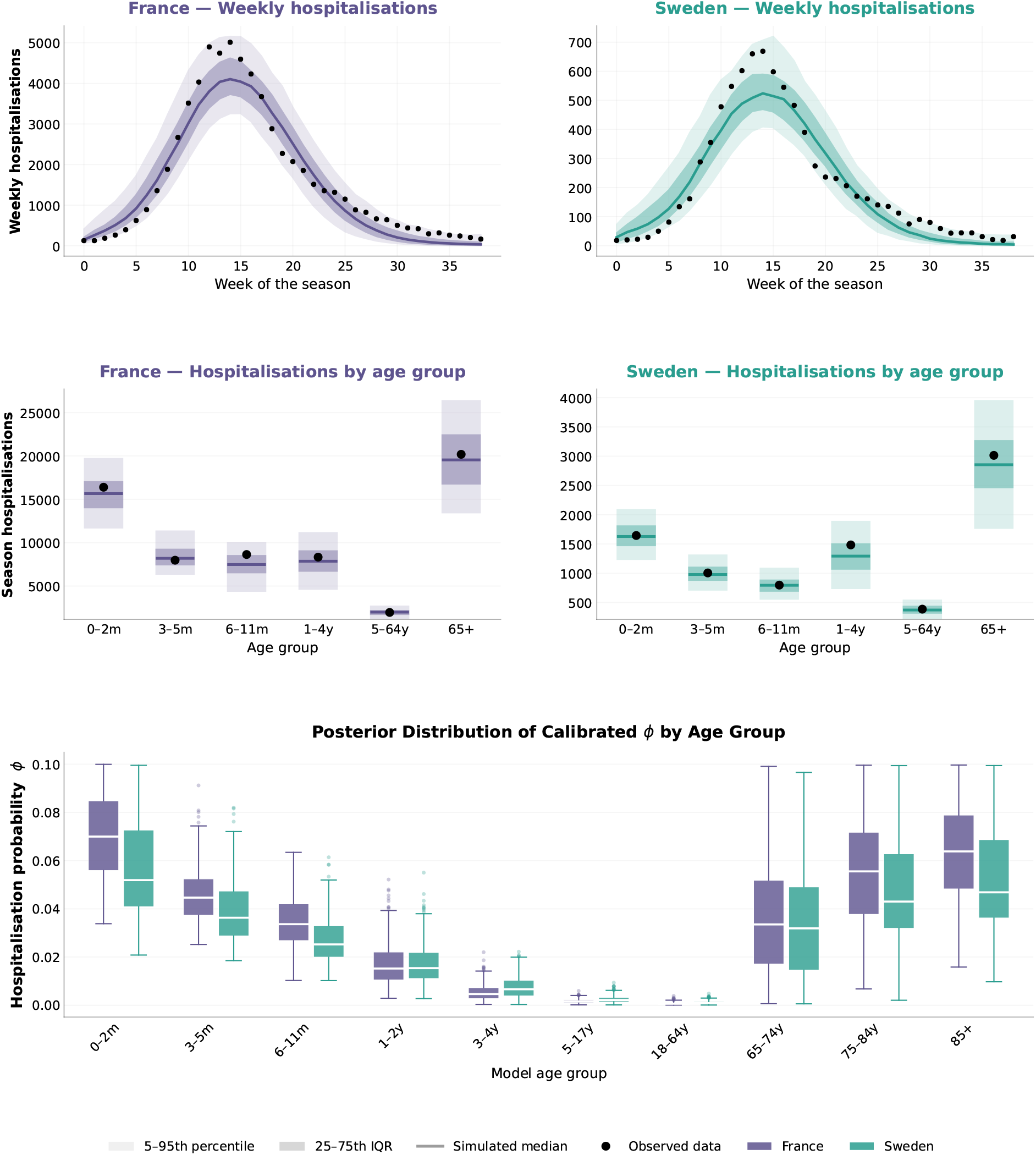
Model calibration results for France (violet) and Sweden (teal) over a realistic respiratory season. **(Top row)** Weekly hospitalisations: black dots represent observed data, while shaded bands and solid lines show the simulated 5–95th percentile (lightest band), 25–75th interquartile range (darker band), and median (solid line) across the calibrated simulation ensemble. **(Middle row)** Total seasonal hospitalisations stratified by age group, expressed in the RespiCompass age stratification: black dots indicate observed values, while bars and horizontal segments report the simulated 5–95th percentile, 25–75th interquartile range, and median, respectively. **(Bottom row)** Posterior distributions of the calibrated age-specific hospitalisation probability *ϕ* across the 10 model age groups, summarised over the top-300 best-fitting simulations selected via Borda ranking; boxes span the interquartile range, whiskers extend to 1.5*×*IQR, and outliers are shown as individual points.

The estimated post-infection hospitalisation probabilities *ϕ*_*i*_ reflect the expected age-dependent pattern of severe RSV disease (Figure 4). In the 0–2 month age group, the median *ϕ* was 0.070 (90% CI: [0.041, 0.098]) for France and 0.052 (90% CI: [0.029, 0.095]) for Sweden, approximately halving by the 6 *−* 11 month age group in both countries. From 65 years onward, the hospitalisation probabilityrose again, reaching a median of 0.064 (90% CI: [0.030, 0.094]) in the 85+ group for France and 0.047(90% CI: [0.024, 0.089]) for Sweden.

### Averted RSV-associated hospitalizations at continental-scale

For each scenario, Figure 5 shows the distribution of the median percentage of averted hospitalisations in infants under one year of age across 28 European countries. At 95% coverage, the monoclonal antibody strategy yields a median reduction of 29.9%, with country-level medians ranging from 27.7% in Norway to 33.9% in Malta. At 25% coverage, the median reduction is 7.8%, ranging from 6.4% in Luxembourg to 8.8% in Croatia. For maternal vaccination, the corresponding median reductions are 22.4% at 95% coverage, spanning from 20.0% in Norway to 25.6% in the Netherlands, and 6.0% at 25% coverage, spanning from 4.8% in Estonia to 6.9% in Cyprus (Figure 5 and Table 3). Across the four coverage levels considered, the percentage of hospitalisations averted scales linearly with coverage, characterised by a proportionality factor 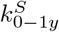whose median across countries is 0.3 for S = la-mAbs and 0.2 for MV, with low inter-country variability.

**Table 3:**
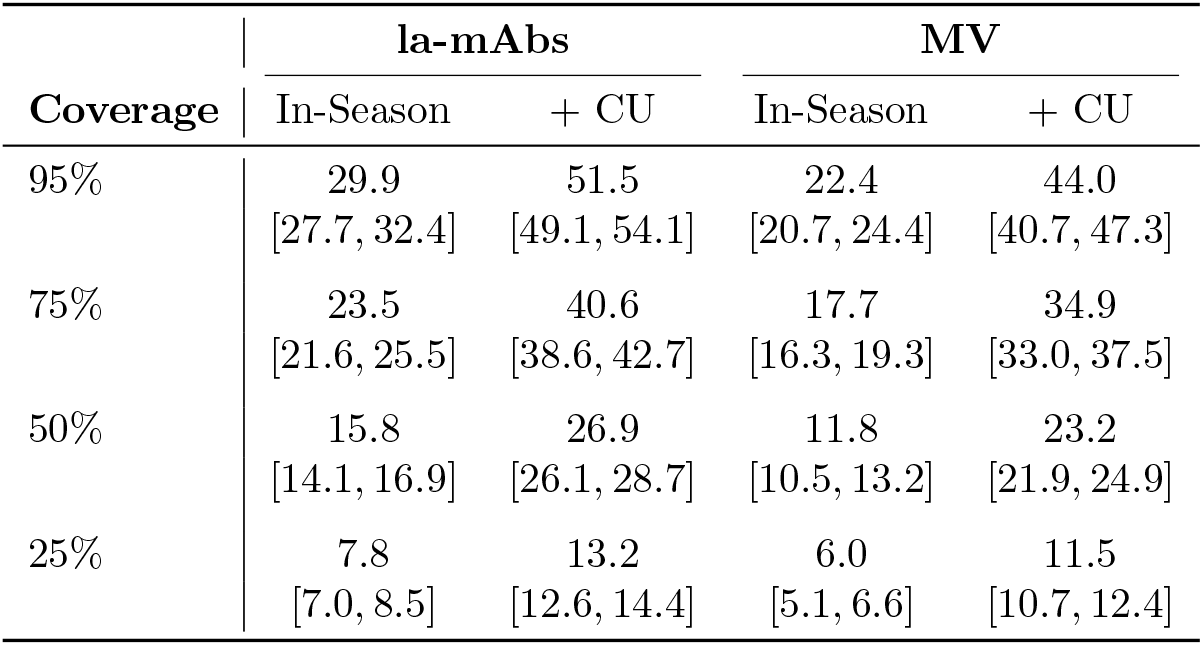
Median percentage reduction in hospitalisations. Intervention 1 refers to monoclonal antibodies (mAbs); Intervention 2 refers to maternal vaccination (MV). The suffix +CU denotes catchup scenarios. Coverage levels are labelled A (95%), B (75%), C (50%), and D (25%).

**Figure 5:**
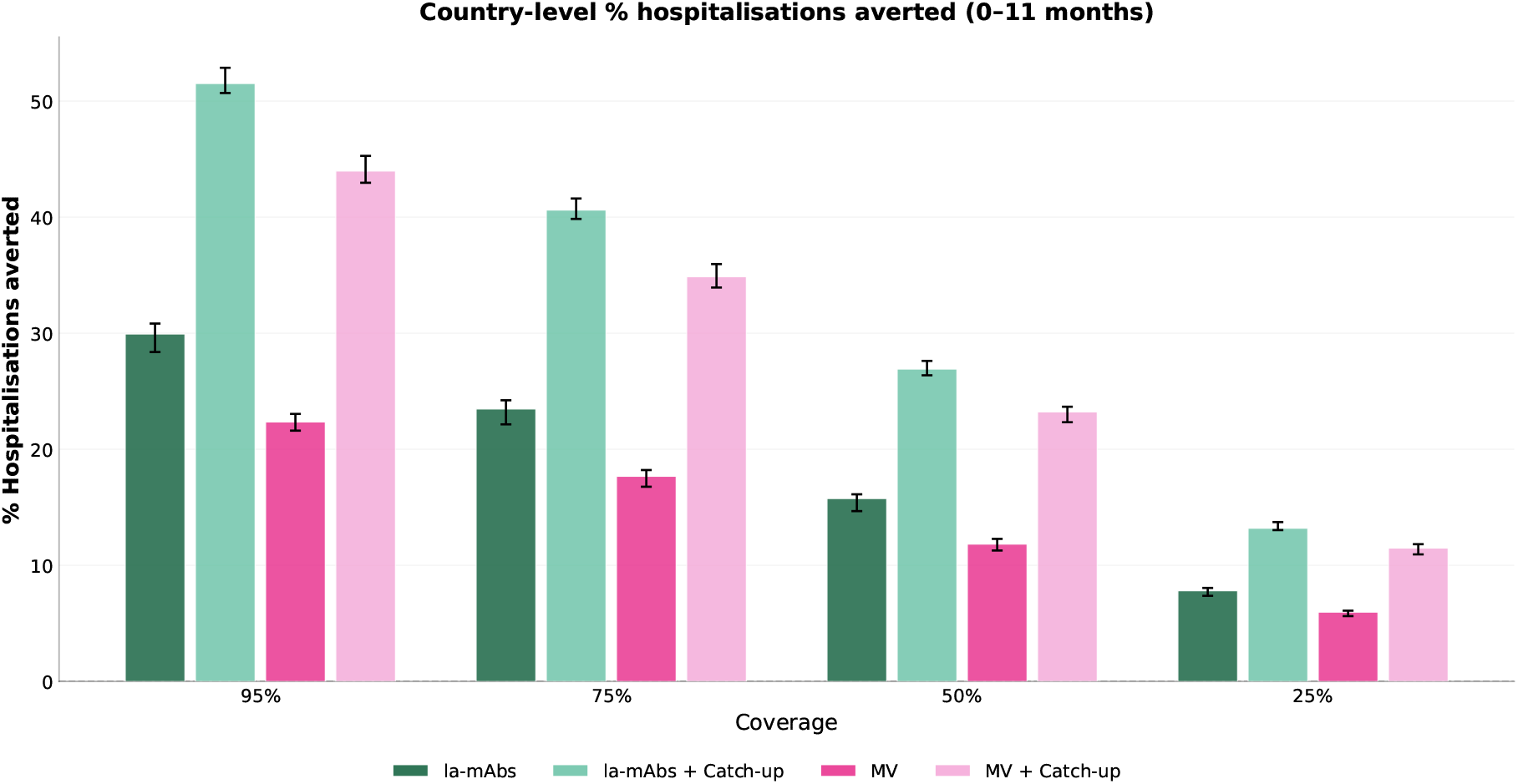
Distribution of country-level median percentage of hospitalisations averted among infants aged 0–11 months, across 28 European countries. For each country, the median percentage of averted hospitalisations is computed over the calibrated simulations; bar heights represent the median of these country-level medians, and whiskers indicate the interquartile range (Q25–Q75) of their distribution across countries. Results are shown by coverage (corresponding to 95%, 75%, 50%, and 25% coverage, respectively) and immunisation strategy. Green bars correspond to long-acting monoclonal antibodies; pink bars correspond to maternal vaccination. Lighter shades denote strategies that include a catch-up campaign (+CU).

The inclusion of a catch-up campaign increases the median percentage of hospitalisations averted across countries to 51.5% at 95% coverage, with country-level medians ranging from 47.3% in Malta to 55.5% in the Netherlands, and 13.2% at 25% coverage, ranging from 12.4% in Malta to 14.5% in the Netherlands. For maternal vaccination with catch-up, the corresponding median reductions are 44.0% at 95% coverage, spanning from 38.8% in Malta to 47.8% in the Netherlands, and 11.5% at 25% coverage, spanning from 10.2% in Estonia to 13.5% in Ireland (Figure 5 and Table 3). Country-specific absolute numbers and percentages of averted hospitalisations for each intervention scenario and coverage level are reported in Supplementary Section S5.

As shown in Figure 6, at 95% coverage, the share of averted hospitalisations falling within the first three months of life is 78.2% (90% CI: [67.3, 82.9]%) for la-mAbs and 72.6% (90% CI: [61.4, 82.6]%) for maternal vaccination, and 61.7% (90% CI: [55.5, 66.3]%) and 57.1% (90% CI: [51.1, 62.3]%) for the corresponding strategies with catch-up. Looking at children aged 1 *−* 4 years, the share of averted hospitalisations is 0.7% (90% CI: [0.0, 3.0]%) for la-mAbs, 1.9% (90% CI: [0.0, 4.6]%) for maternal vaccination, 1.3% (90% CI: [0.0, 3.0]%) for la-mAbs with catch-up, and 2.0% (90% CI: [0.3, 3.5]%) for maternal vaccination with catch-up. Finally, for individuals over 65 years, the corresponding shares are 1.3% (90% CI: [0.0, 10.8]%) for la-mAbs, 7.0% (90% CI: [0.0, 16.4]%) for maternal vaccination, 2.9% (90% CI: [0.0, 8.1]%) for la-mAbs with catch-up, and 5.5% (90% CI: [0.0, 11.1]%) for maternal vaccination with catch-up. The full age-specific distribution of averted hospitalisations for all intervention scenarios and coverage levels is reported in Supplementary Section S6.

**Figure 6:**
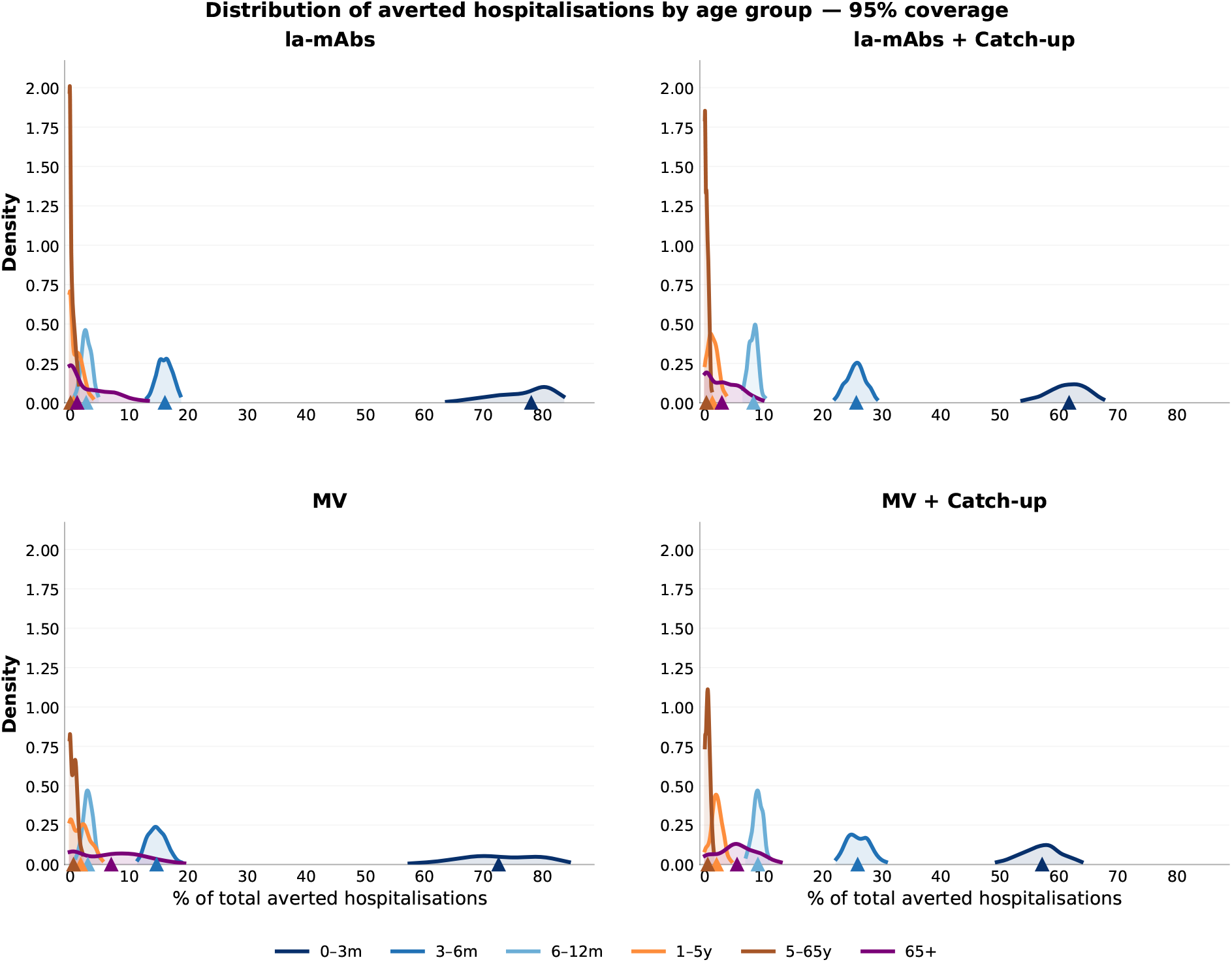
Distribution of averted hospitalisations by age group at 95% coverage. Each panel corresponds to one intervention strategy. Curves represent kernel density estimates of the age-group share of total European averted hospitalisations. Triangles on the horizontal axis mark the median of each distribution.

At 95% of coverage, the median number of doses needed to avert one hospitalisation in the 0–11 month age group across 28 European countries is 48 for monoclonal antibody administration and 50 for maternal vaccination. Cross-country values ranging from 38 for France to 77 for the Netherlands and from 41 for France to 89 for the Netherlands, respectively. When a catch-up programme is added, these medians increase to 54 and 57, with corresponding ranges from 41 for France to 89 for Malta and from 42 for France to 99 for Malta (Figure 7). Country-specific distributions of doses per averted hospitalisation for all intervention scenarios and coverage levels are provided in Supplementary Section S7.

**Figure 7:**
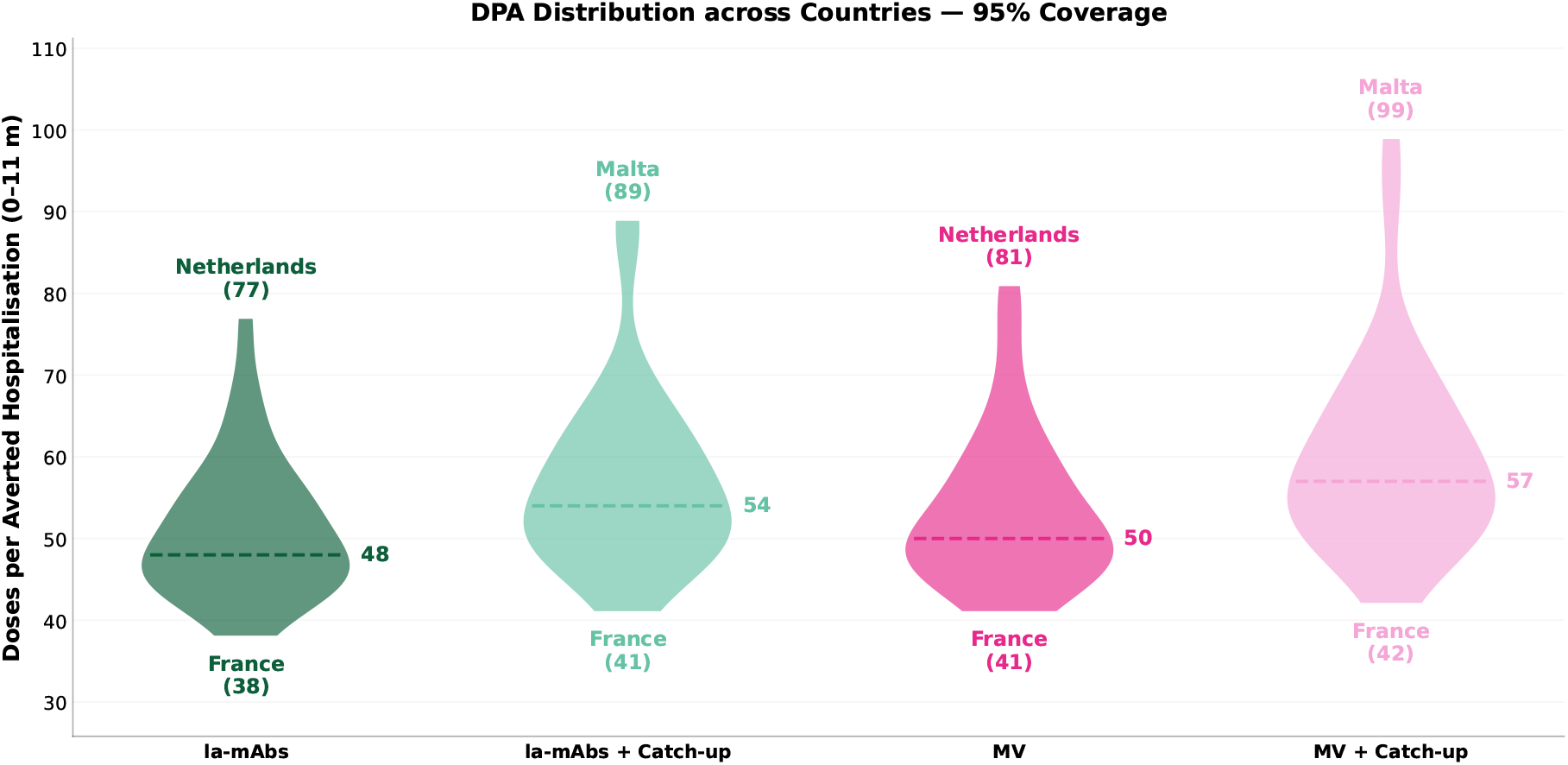
Distribution of Doses per Averted Hospitalisation (DPA) in the 0 *−* 11 month age group across European countries, at 95% coverage. Each violin represents the spread of country-level median DPA values for a given intervention strategy. The dashed horizontal line indicates the median across countries. Labels at the top and bottom of each violin identify the worst-performing (highest DPA) and best-performing (lowest DPA) country, respectively, with their corresponding values.

## Discussion

Using a data-driven dynamic transmission model calibrated independently for 28 European countries, we evaluated the population-level impact of two universal RSV prevention strategies in infants: longacting monoclonal antibodies and maternal vaccination. To our knowledge, this is the first continental-scale evaluation of these strategies built on a harmonised modelling framework with country-specific calibration.

Three main findings emerge. First, both strategies substantially reduce infant hospitalisations in every country examined. At 95% coverage of newborns delivered between September and March, monoclonal antibodies achieved a cross-country median reduction of 29.9% in hospitalisations under one year of age, compared to 22.4% for maternal vaccination. Adding a seasonal catch-up campaign at season onset, targeting infants up to six months old, nearly doubled the impact of both strategies. Second, the relative effectiveness of these strategies is remarkably uniform across countries despite substantial demographic and epidemiological heterogeneity. Country profiles vary widely in the share of infants under one year of age, in baseline hospitalisation burden, and in the seasonal distribution of births. Yet country-level median reductions fell within a narrow window (from 27.7% in Norway to 33.9% in Malta for monoclonal antibodies, and from 20.0% in Norway to 25.6% in the Netherlands for maternal vaccination). Across countries, coverage and impact followed an approximately country-invariant linear relationship, with every 10 percentage points of coverage averting roughly 3% of hospitalisations for monoclonal antibodies and 2% for maternal vaccination. No saturation effects emerged within the range of coverage explored, suggesting that each additional administered dose contributes a comparable marginal benefit. Despite accounting for indirect protection through reduced susceptibility to infection, the benefits also remained concentrated in the directly targeted age group across all countries and scenarios, with modest indirect effects on older children and the elderly. Taken together, these patterns indicate that the relative impact of these interventions is primarily governed by the intrinsic effectiveness rather than by country-specific contextual factors, and is therefore comparable across European settings. Third, a different picture emerges when impact is expressed as doses required per averted hospitalisation. At 95% coverage, the cross-country median is 47 doses for monoclonal antibodies and 49 for maternal vaccination, with France at one extreme and the Netherlands and Malta at the opposite end. The number of doses needed per averted hospitalisation is lower in countries with a higher baseline burden of infant hospitalisations and, unlike the relative reduction, depends substantially on country-specific epidemiology.

The methodological contribution of this work lies in providing a harmonised framework for cross-country assessment, enabling comparisons across settings under a consistent set of assumptions while complementing existing country-specific analyses. Interestingly, our country-level estimates are broadly consistent with previous studies where direct comparisons are possible. For example, Krauer et al. estimated that seasonal immunisation of infants younger than 6 months at 70% coverage would reduce RSV-related hospitalisations and ICU admissions during the first year of life by 39.3% ^14^. A similar effect was estimated by our model for Germany, where long-acting monoclonal antibodies with catch-up at 75% coverage coverage reduced these outcomes by 41.8% (Supplementary Information, Table S64). Likewise, their maternal vaccination scenario yielded a median reduction of 12.9% at 40% coverage ^14^, compared with 12.1% in our corresponding scenario at 50% coverage (Supplementary Information, Table S68). The approximately linear increase in impact with coverage observed in our analysis is also in agreement with their results. Furthermore, the limited indirect effects in non-targeted age groups are consistent with previous national and sub-national modelling studies ^12–14^ and reflect the limited role of infants in onward RSV transmission ^34^. Finally, the estimated number of doses required to avert one hospitalisation is broadly consistent with both clinical trial evidence and previous national modelling studies. Our population-level estimates align with those reported in the MELODY trial (65, 95% CI [41, 158]) ^31^, while our estimates for France closely match the value of 39 reported by Brault et al. ^12^.

Several limitations should be considered. First, the model relies on the burden estimates from RespiCompass 2024*/*2025, themselves informed by estimates drawn predominantly from north-western European countries ^2,25^; extrapolating these patterns to settings with different demographic, climatic, or healthcare profiles may attenuate the true cross-country heterogeneity. Second, our scenario design assumes catch-up doses are administered simultaneously at season onset, whereas real-world programmes deliver catch-up gradually ^7^; this simplification likely underestimates its impact, since immunisation closer to the epidemic peak would confer greater protection during peak transmission. Finally, we assume uniform product efficacy across countries in line with pivotal trials, while real-world effectiveness may differ. In particular, recent evidence indicates that maternal vaccination effectiveness depends substantially on gestational age at vaccination, with longer intervals to delivery yielding higher transplacental antibody transfer and stronger infant protection ^35^.

Our work provides the epidemiological foundation for country-specific economic evaluations, which represent the natural next step in this line of research. Further extensions could explore the population-level effectiveness of immunisation strategies currently emerging in European programmes, such as the combined use of maternal vaccination and monoclonal antibodies ^7^, and identify optimal immunisation timing patterns.

These findings have direct implications for how RSV prevention policies can be coordinated across Europe. The marked homogeneity in relative effectiveness and the country-invariant coverage-impact relationship support harmonised European guidelines when policy aims to achieve a defined level of reduction in RSV burden across the continent. The introduction of a seasonal catch-up campaign emerges as a particularly relevant element for such coordinated guidance, given its potential to substantially increase impact regardless of national context.

## Data Availability

All data produced in the present work are contained in the manuscript

## Author contributions

All authors contributed to the design of the project and the model. EV implemented the model, the scenario pipeline, and data analyses. EV wrote the first draft of the manuscript with guidance from NG and critical revisions from MM and DP. All authors contributed to interpreting the results, and read, reviewed, and approved the final version and the submission of the manuscript.

## Data sharing

All code and direct links to the input data are available at https://github.com/Eli-13/RSV_Europe, and the project output data are reported in Supplementary Sections S3 to S8.

## Acknowledgments

This publication is part of the project PNRR-NGEU which has received funding from the MUR-DM 630/2024.

