## SupplementaryInformation for "A continental-scale scenario modelling framework for evaluating infant RSV immunisation strategies across Europe"

**S8 Sensitivity analysis to the waning immunity assumptions** **89**

### S1 Model equations

#### S1.1 Force of infection and seasonal transmission rate

The force of infection acting on susceptible individuals in age group $i$ at immunity level $j$ in the baseline system ($k = 0$) is:

$$\Lambda_{i,j,0}(t) = \lambda_j \beta(t) \sum_{i'} c_{ii'} \frac{\sum_{j',k'} I_{i',j',k'}(t)}{N_{i'}(t)} \tag{1}$$

where $c_{ii'}$ is the contact rate between age groups $i$ and $i'$, $N_{i'}(t)$ the total population in age group $i'$ at time $t$, and $\beta(t)$ the time-varying seasonal transmission rate, defined as:

$$\beta(t) = \beta_{\text{min}} + \frac{\beta_{\text{max}} - \beta_{\text{min}}}{2} \left[1 + \cos\left((t - t_{\text{peak}}) \frac{2\pi}{365}\right)\right] \tag{2}$$

where $\beta_{\text{min}}$ and $\beta_{\text{max}}$ are the minimum and maximum values of the transmission rate, and $t_{\text{peak}}$ is the day at which transmission peaks.

#### S1.2 Intervention effectiveness

For each intervention, we compute the ratio $\rho$ between the median effectiveness against hospitalisation and that against infection. For each simulation $s$, we sample vaccine effectiveness against hospitalisation $\text{VE}_s^{(\text{hosp})}$ from a Beta distribution fitted to the reported 95% confidence interval, then derive effectiveness against infection using the fixed ratio:

$$\text{VE}_s^{(\text{inf})} = \frac{\text{VE}_s^{(\text{hosp})}}{\rho} \tag{3}$$

Effectiveness against infection modifies the force of infection in the treated system:

$$\Lambda_{i,j,k}(t) = \lambda_j \left(1 - \text{VE}_k^{(\text{inf})}\right) \beta(t) \sum_{i'} c_{ii'} \frac{\sum_{j',k'} I_{i',j',k'}(t)}{N_{i'}(t)} \tag{4}$$

where $\text{VE}_k^{(\text{inf})}$ denotes the effectiveness against infection for treatment status $k \neq 0$. For untreated individuals ($k = 0$), $\text{VE}_k^{(\text{inf})} = 0$ and the expression reduces to the baseline force of infection in Equation (1). The conditional effectiveness against hospitalisation given infection is:

$$\text{VE}_s^{(\text{hosp}|\text{inf})} = \frac{\text{VE}_s^{(\text{hosp})} - \text{VE}_s^{(\text{inf})}}{1 - \text{VE}_s^{(\text{inf})}} \tag{5}$$

This quantity reduces the age-specific probability of hospitalisation for treated individuals who experience breakthrough infection.

### S2 Detailed Calibration Procedure

We calibrated the model independently for each country over the period covered by the calibration data. The procedure described below applies to each country separately.

#### S2.1 Prior specifications

##### Transmission parameters

We conditioned the prior for $\beta_{\max}$ on the sampled value of $\beta_{\min}$ in each simulation, ensuring that $\beta_{\min} < \beta_{\max}$. The joint constraint defines the support $\mathcal{D}$ reported in Table 1 of the main text:

$$\mathcal{D} = \{(\beta_{\min}, \beta_{\max}) : 4.9\times10^{-3} \leq \beta_{\min} \leq 3.5\times10^{-2},\ \beta_{\min} + 1.7\times10^{-3} \leq \beta_{\max} \leq 5.0\times10^{-2}\}.$$

The peak timing $t_{\text{peak}}$ was sampled uniformly over the period from 1 September to 1 March, with day indexing reflecting the seasonal window of analysis.

##### Hierarchical priors on hospitalisation probabilities

To encode the well-documented age dependence of severe RSV disease, we constructed hierarchical priors on the ten $\phi_i$. We assigned a broad uniform prior to the youngest age group (0–2 months):

$$\phi_{0\text{–}2\text{m}} \sim \text{Uniform}(10^{-4},\ 10^{-1}).$$

For each subsequent age group up to 64 years, the prior was constrained to lie below the previously sampled value:

$$\phi_i \sim \text{Uniform}(10^{-4},\ \phi_{i-1}), \quad i \in \{3\text{–}5\text{m}, 6\text{–}11\text{m}, 1\text{–}2\text{y}, 3\text{–}4\text{y}, 5\text{–}17\text{y}, 18\text{–}64\text{y}\}.$$

From 65 years onward, the direction was reversed, with each subsequent age group constrained to lie above the previous one but below the maximum value of the youngest group:

$$\phi_i \sim \text{Uniform}(\phi_{i-1},\ \phi_{0\text{–}2\text{m}}), \quad i \in \{65\text{–}74\text{y}, 75\text{–}84\text{y}, 85+\text{y}\}.$$

This hierarchical structure enforces the empirically observed pattern of high hospitalisation risk among neonates and the elderly, with low risk in school-age and adult age groups, while allowing the data to inform the precise values.

#### S2.2 Model initialisation and reference database

To initialise the model with equilibrium compartment values, we ran the model for 100 years using 2023 demographic data, saving the population state on 31 August of each year. We sampled 10,000 combinations of $(\beta_{\min}, \beta_{\max})$ using a Poisson disk algorithm [1] to achieve a uniform exploration of the constrained parameter space, and independently sampled $t_{\text{peak}}$ from its prior. We then filtered the population states from the last 10 years of simulation, retaining only those corresponding to non-extinct epidemics, defined as simulations in which the infected and recovered compartments did not remain identically zero throughout the period. We stored the corresponding parameter values and age-stratified population distributions across compartments in a reference database.

#### S2.3 ABC simulation-budget approach

We fitted the calibrated parameters using a simulation-budget approach within the Approximate Bayesian Computation (ABC) framework [2, 3]. For each country, we drew 100,000 parameter sets $(\beta_{\min}, \beta_{\max}, t_{\text{peak}})$ from the same prior. For each sampled triplet, we retrieved initial conditions from the reference database by selecting the entry with the smallest Euclidean distance in $(\beta_{\min}, \beta_{\max})$ among those whose $t_{\text{peak}}$ fell within 5 days of the sampled value, and distributed the population across compartments at $t_0$ accordingly. We then ran each simulation over the calibration window.

For each of the 100,000 simulations, we drew 100 independent sets of age-specific hospitalisation probabilities $\phi_i$ from their hierarchical priors. For each set, we estimated the hospitalisation curve by age group, aggregated the results over the full season to obtain cumulative hospitalisations per age class, and compared them against the calibration targets using the Mean Absolute Percentage Error (MAPE):

$$\text{MAPE} = \frac{1}{n} \sum_{i=1}^{n} \left| \frac{H_i^{\text{sim}} - H_i^{\text{obs}}}{H_i^{\text{obs}}} \right| \tag{6}$$

where $n$ denotes the number of age groups, $H_i^{\text{sim}}$ the simulated cumulative hospitalisations in age group $i$, and $H_i^{\text{obs}}$ the corresponding calibration target. We retained the $\phi_i$ set that minimised the MAPE for each simulation. We ranked each simulation separately by two criteria: the Root Mean Square Error (RMSE) between the simulated and observed weekly hospitalisation curves,

$$\text{RMSE} = \sqrt{\frac{1}{W} \sum_{w=1}^{W} \left(h_w^{\text{sim}} - h_w^{\text{obs}}\right)^2} \tag{7}$$

where $W$ denotes the number of weeks and $h_w$ the weekly hospitalisation count; and the MAPE across age groups defined above. We summed the two ranks for each simulation and retained the 300 with the lowest combined Borda score.

### S3 Scenario Evaluation: Formal Definitions

This section provides the formal definitions of the outcome metrics described in the main text.

For each country $c$, simulation $s$, and intervention scenario $S$, we denote by $H_i^{E,s,c}$ the cumulative hospitalisations in age group $i$ under the baseline scenario E (summed over all weeks of the season), and analogously $H_i^{S,s,c}$ under intervention scenario $S$, which identifies a specific combination of immunisation strategy and coverage level $l_S$.

##### Averted hospitalisations

The number of averted hospitalisations in age group $i$ for simulation $s$ is defined as

$$\delta_i^{S,s,c} = H_i^{E,s,c} - H_i^{S,s,c}, \tag{8}$$

where positive values indicate a reduction in hospitalisations due to the intervention. Repeating this paired calculation over all retained simulations yields an empirical distribution, summarised at the country level as the median, $\widetilde{\delta}_i^{S,c} = \text{median}_s(\delta_i^{S,s,c})$.

##### Percentage reduction in hospitalisations

To enable comparison across countries with different baseline burdens, we computed a simulation-level percentage of hospitalisations averted over the first year of life (age groups $i \in \{0, 1, 2\}$):

$$p^{S,s,c} = \frac{\sum_{i=0}^{2} \delta_i^{S,s,c}}{\sum_{i=0}^{2} H_i^{E,s,c}} \times 100, \tag{9}$$

summarised at the country level as $\widetilde{p}^{S,c} = \text{median}_s(p^{S,s,c})$.

##### Proportionality constant

To assess the relationship between coverage and intervention impact, we defined the proportionality constant

$$k^{S,c} = \frac{\widetilde{p}_{l_S}^{S,c}}{l_S}, \tag{10}$$

where $\widetilde{p}_{l_S}^{S,c}$ is the median percentage of hospitalisations averted at coverage level $l_S$.

##### Age-group share of averted hospitalisations

To quantify how averted hospitalisations distribute across age groups at the European level, we aggregated the averted hospitalisations across all 28 countries at the simulation level, and then computed the share attributable to each age group. For a given simulation $s$ and intervention scenario $S$, the fraction of averted hospitalisations attributable to age group $i$ is defined as

$$w_i^{S,s} = \frac{\sum_c \delta_i^{S,s,c}}{\sum_c \sum_k \delta_k^{S,s,c}}. \tag{11}$$

Repeating this calculation across all retained simulations yields an empirical distribution of age-group shares.

##### Doses per averted hospitalisation

To assess the efficiency of each immunisation strategy, we computed the number of doses per averted hospitalisation (DPA). Letting $D^{S,c}$ denote the total doses administered (birth doses plus catch-up doses, where applicable) for country $c$ under scenario $S$, for each simulation $s$ in which at least one hospitalisation is averted, we defined

$$\mathrm{DPA}^{S,s,c} = \frac{D^{S,c}}{\sum_{i=0}^{2} \delta_i^{S,s,c}}, \tag{12}$$

with country-level point estimate $\widetilde{\mathrm{DPA}}^{S,c} = \mathrm{median}_s(\mathrm{DPA}^{S,s,c})$.

### S4 Calibration results

This section provides the complete calibration outputs for all 28 countries included in the analysis, complementing the country-specific results for France and Sweden presented in the main text. It is organised in three parts: (i) two summary tables reporting calibration performance metrics across countries; (ii) detailed country-level tables for the weekly hospitalisation target; and (iii) detailed country-level tables for the age-stratified target.

#### S4.1 Calibration performance metrics

To summarise the calibration performance across countries, we report two synthetic tables, one for each calibration target. For each country, we report the percentage of observed data points falling within the 50% and 90% credible intervals of the simulated distributions, together with standard error metrics: the Root Mean Square Error (RMSE), the Mean Absolute Error (MAE), and the Weighted Mean Absolute

Percentage Error (WMAPE). Table S1 reports these metrics against the weekly hospitalisation curve, and Table S2 against the cumulative age-stratified hospitalisation burden.

Table S1: **Calibration performance against the weekly hospitalisation curve.** For each country, we report the percentage of weekly observed data points falling within the 50% and 90% credible intervals of the simulated distributions, and the corresponding error metrics across the 39 weeks of the calibration window: Root Mean Square Error (RMSE), Mean Absolute Error (MAE), and Weighted Mean Absolute Percentage Error (WMAPE).

| **Country** | **Cov.** 50% | **Cov.** 90% | **RMSE** | **MAE** | **WMAPE** |
|---|---|---|---|---|---|
| Austria | 28.2% | 97.4% | 53.15 | 43.26 | 0.264 |
| Belgium | 25.6% | 74.4% | 79.04 | 64.69 | 0.269 |
| Bulgaria | 25.6% | 84.6% | 40.66 | 33.91 | 0.235 |
| Croatia | 38.5% | 89.7% | 26.14 | 19.92 | 0.256 |
| Cyprus | 38.5% | 94.9% | 7.68 | 6.09 | 0.318 |
| Czechia | 25.6% | 92.3% | 60.96 | 52.71 | 0.241 |
| Denmark | 23.1% | 87.2% | 38.44 | 33.47 | 0.270 |
| Estonia | 46.2% | 97.4% | 9.17 | 7.05 | 0.274 |
| Finland | 43.6% | 84.6% | 33.64 | 24.56 | 0.267 |
| France | 30.8% | 100.0% | 387.90 | 319.90 | 0.197 |
| Germany | 28.2% | 100.0% | 364.07 | 295.37 | 0.185 |
| Greece | 30.8% | 92.3% | 53.53 | 44.35 | 0.217 |
| Hungary | 28.2% | 89.7% | 53.96 | 45.81 | 0.234 |
| Ireland | 28.2% | 84.6% | 37.98 | 32.45 | 0.263 |
| Italy | 25.6% | 82.1% | 239.99 | 207.97 | 0.200 |
| Latvia | 41.0% | 100.0% | 13.01 | 10.50 | 0.292 |
| Lithuania | 43.6% | 87.2% | 16.08 | 12.44 | 0.242 |
| Luxembourg | 48.7% | 92.3% | 6.45 | 4.64 | 0.416 |
| Malta | 51.3% | 84.6% | 5.00 | 3.60 | 0.374 |
| Netherlands | 28.2% | 92.3% | 61.05 | 49.54 | 0.211 |
| Norway | 28.2% | 89.7% | 33.39 | 26.85 | 0.241 |
| Poland | 38.5% | 87.2% | 231.30 | 173.31 | 0.270 |
| Portugal | 25.6% | 97.4% | 53.37 | 45.41 | 0.237 |
| Romania | 38.5% | 97.4% | 89.12 | 75.31 | 0.200 |
| Slovakia | 30.8% | 92.3% | 28.94 | 24.17 | 0.223 |
| Slovenia | 35.9% | 89.7% | 15.37 | 12.08 | 0.301 |
| Spain | 33.3% | 97.4% | 173.95 | 146.14 | 0.212 |
| Sweden | 30.8% | 94.9% | 58.17 | 46.54 | 0.218 |

Table S2: **Calibration performance against the cumulative age-stratified hospitalisation burden.** For each country, we report the percentage of age-stratified observed data points falling within the 50% and 90% credible intervals of the simulated distributions, and the corresponding error metrics: Root Mean Square Error (RMSE), Mean Absolute Error (MAE), and Weighted Mean Absolute Percentage Error (WMAPE).

| **Country** | **Cov.** 50% | **Cov.** 90% | **RMSE** | **MAE** | **WMAPE** |
|---|---|---|---|---|---|
| Austria | 100.0% | 100.0% | 75.98 | 50.17 | 0.047 |
| Belgium | 100.0% | 100.0% | 67.40 | 56.92 | 0.036 |
| Bulgaria | 100.0% | 100.0% | 56.07 | 43.17 | 0.046 |
| Croatia | 100.0% | 100.0% | 45.32 | 37.67 | 0.074 |
| Cyprus | 100.0% | 100.0% | 11.60 | 9.25 | 0.074 |
| Czechia | 100.0% | 100.0% | 94.83 | 69.92 | 0.049 |
| Denmark | 83.3% | 100.0% | 97.17 | 54.58 | 0.068 |
| Estonia | 100.0% | 100.0% | 15.26 | 11.67 | 0.070 |
| Finland | 83.3% | 100.0% | 51.77 | 31.25 | 0.052 |
| France | 83.3% | 100.0% | 654.11 | 543.58 | 0.051 |
| Germany | 100.0% | 100.0% | 878.27 | 596.08 | 0.058 |
| Greece | 100.0% | 100.0% | 70.41 | 46.33 | 0.035 |
| Hungary | 100.0% | 100.0% | 68.03 | 46.50 | 0.037 |
| Ireland | 100.0% | 100.0% | 74.72 | 49.33 | 0.062 |
| Italy | 100.0% | 100.0% | 779.46 | 459.33 | 0.068 |
| Latvia | 100.0% | 100.0% | 25.22 | 16.67 | 0.071 |
| Lithuania | 100.0% | 100.0% | 18.18 | 15.42 | 0.046 |
| Luxembourg | 100.0% | 100.0% | 10.01 | 8.83 | 0.122 |
| Malta | 66.7% | 83.3% | 20.20 | 14.92 | 0.238 |
| Netherlands | 100.0% | 100.0% | 118.84 | 71.83 | 0.047 |
| Norway | 100.0% | 100.0% | 62.71 | 53.75 | 0.074 |
| Poland | 83.3% | 100.0% | 641.70 | 486.75 | 0.117 |
| Portugal | 100.0% | 100.0% | 43.69 | 39.00 | 0.031 |
| Romania | 100.0% | 100.0% | 176.08 | 119.17 | 0.049 |
| Slovakia | 100.0% | 100.0% | 48.91 | 36.67 | 0.052 |
| Slovenia | 100.0% | 100.0% | 19.42 | 15.08 | 0.058 |
| Spain | 83.3% | 100.0% | 498.36 | 294.58 | 0.066 |
| Sweden | 100.0% | 100.0% | 102.88 | 68.83 | 0.050 |

#### S4.2 Country-level detailed calibration outputs

For each of the 28 countries, we report two tables corresponding to the two calibration targets: the weekly hospitalisation curve (Tables S3–S30) and the cumulative age-stratified hospitalisation burden over the season (Tables S31–S58). Each table reports the observed values alongside the 5th, 25th, 50th (median), 75th, and 95th percentiles of the simulated distributions across the 300 retained simulations, enabling a pointwise comparison between observations and model output. Tables are ordered alphabetically by country, with weekly tables presented first followed by age-stratified tables.

##### S4.2.1 Weekly hospitalisation calibration tables

Table S3: **Weekly calibration – Austria.** Observed weekly hospitalisation counts and the 5th, 25th, 50th (median), 75th, and 95th percentiles of the simulated distributions across the 300 retained simulations.

| Week | Observed | Sim Q05 | Sim Q25 | Sim Median | Sim Q75 | Sim Q95 |
|---|---|---|---|---|---|---|
| 0 | 12.0 | 3.0 | 13.0 | 24.0 | 34.0 | 51.0 |
| 1 | 17.0 | 5.0 | 24.0 | 42.0 | 60.2 | 84.1 |
| 2 | 18.0 | 9.9 | 32.0 | 55.0 | 81.0 | 110.0 |
| 3 | 28.0 | 12.0 | 46.5 | 73.0 | 104.0 | 139.0 |
| 4 | 41.0 | 23.0 | 59.0 | 97.0 | 132.2 | 171.0 |
| 5 | 60.0 | 33.0 | 76.8 | 124.0 | 161.0 | 199.1 |
| 6 | 76.0 | 49.9 | 103.0 | 155.0 | 200.2 | 247.1 |
| 7 | 149.0 | 74.7 | 133.8 | 189.5 | 238.2 | 295.1 |
| 8 | 187.0 | 106.0 | 167.0 | 230.0 | 281.2 | 352.1 |
| 9 | 268.0 | 139.0 | 208.0 | 270.5 | 327.8 | 384.0 |
| 10 | 370.0 | 188.8 | 252.8 | 306.0 | 362.0 | 446.1 |
| 11 | 397.0 | 225.9 | 288.0 | 340.0 | 389.5 | 476.1 |
| 12 | 469.0 | 266.9 | 318.8 | 363.5 | 416.0 | 502.1 |
| 13 | 509.0 | 289.9 | 341.0 | 379.0 | 432.8 | 513.0 |
| 14 | 529.0 | 302.9 | 352.0 | 388.5 | 437.0 | 530.0 |
| 15 | 465.0 | 292.9 | 342.0 | 380.0 | 427.0 | 532.0 |
| 16 | 412.0 | 284.0 | 327.8 | 361.0 | 418.5 | 510.5 |
| 17 | 384.0 | 259.9 | 301.0 | 340.5 | 394.0 | 495.1 |
| 18 | 281.0 | 221.8 | 271.0 | 313.5 | 360.2 | 445.1 |
| 19 | 214.0 | 188.9 | 233.8 | 279.0 | 325.5 | 397.3 |
| 20 | 189.0 | 149.0 | 198.0 | 239.0 | 287.0 | 370.0 |
| 21 | 168.0 | 115.0 | 163.8 | 202.5 | 244.2 | 309.1 |
| 22 | 163.0 | 84.9 | 127.8 | 169.0 | 203.2 | 264.4 |
| 23 | 128.0 | 61.0 | 102.0 | 139.0 | 171.0 | 216.1 |
| 24 | 135.0 | 46.0 | 77.8 | 108.5 | 138.2 | 183.1 |
| 25 | 123.0 | 27.0 | 56.0 | 84.0 | 113.2 | 148.2 |
| 26 | 81.0 | 20.0 | 42.0 | 64.0 | 89.0 | 130.1 |
| 27 | 102.0 | 13.9 | 29.8 | 50.0 | 70.0 | 106.0 |
| 28 | 70.0 | 9.0 | 22.0 | 34.5 | 56.2 | 93.0 |
| 29 | 74.0 | 5.0 | 15.0 | 27.5 | 45.0 | 73.0 |
| 30 | 44.0 | 3.0 | 9.0 | 19.0 | 33.2 | 68.1 |
| 31 | 42.0 | 2.0 | 6.0 | 14.0 | 26.2 | 58.1 |
| 32 | 34.0 | 1.0 | 5.0 | 10.0 | 22.2 | 47.0 |
| 33 | 36.0 | 0.0 | 3.0 | 8.0 | 17.2 | 43.1 |
| 34 | 25.0 | 0.0 | 2.0 | 6.0 | 14.0 | 34.1 |
| 35 | 28.0 | 0.0 | 2.0 | 4.0 | 13.0 | 33.0 |
| 36 | 15.0 | 0.0 | 1.0 | 4.0 | 11.0 | 32.1 |
| 37 | 26.0 | 0.0 | 1.0 | 3.0 | 9.0 | 33.0 |
| 38 | 11.0 | 0.0 | 0.0 | 2.0 | 8.0 | 29.0 |

Table S4: **Weekly calibration – Belgium.** Observed weekly hospitalisation counts and the 5th, 25th, 50th (median), 75th, and 95th percentiles of the simulated distributions across the 300 retained simulations.

| Week | Observed | Sim Q05 | Sim Q25 | Sim Median | Sim Q75 | Sim Q95 |
|---|---|---|---|---|---|---|
| 0 | 9.0 | 45.9 | 69.0 | 95.0 | 136.2 | 171.0 |
| 1 | 15.0 | 42.0 | 59.0 | 84.5 | 123.2 | 166.0 |
| 2 | 24.0 | 42.0 | 61.0 | 89.5 | 125.2 | 176.0 |
| 3 | 45.0 | 53.0 | 81.0 | 109.0 | 154.2 | 213.1 |
| 4 | 44.0 | 70.9 | 108.0 | 142.0 | 192.2 | 256.1 |
| 5 | 92.0 | 95.9 | 146.8 | 190.5 | 253.0 | 314.3 |
| 6 | 133.0 | 136.7 | 194.2 | 241.0 | 306.2 | 390.1 |
| 7 | 196.0 | 166.8 | 248.5 | 304.5 | 368.0 | 445.3 |
| 8 | 290.0 | 220.0 | 308.8 | 370.5 | 440.2 | 520.3 |
| 9 | 394.0 | 271.9 | 366.0 | 424.5 | 500.5 | 599.1 |
| 10 | 518.0 | 331.9 | 417.8 | 479.0 | 549.2 | 677.0 |
| 11 | 600.0 | 381.9 | 452.8 | 516.5 | 593.0 | 704.2 |
| 12 | 711.0 | 408.7 | 489.0 | 549.5 | 619.0 | 737.2 |
| 13 | 797.0 | 427.9 | 506.5 | 563.0 | 635.0 | 754.2 |
| 14 | 705.0 | 431.9 | 509.8 | 566.0 | 641.0 | 758.0 |
| 15 | 689.0 | 427.9 | 503.0 | 559.0 | 621.0 | 747.2 |
| 16 | 645.0 | 393.9 | 478.0 | 541.0 | 601.5 | 731.1 |
| 17 | 546.0 | 374.9 | 445.0 | 504.5 | 572.0 | 669.2 |
| 18 | 404.0 | 328.8 | 399.8 | 460.5 | 525.0 | 614.7 |
| 19 | 337.0 | 285.9 | 353.5 | 403.5 | 479.2 | 564.2 |
| 20 | 306.0 | 235.9 | 300.5 | 359.5 | 413.2 | 504.3 |
| 21 | 251.0 | 190.9 | 245.8 | 295.5 | 358.2 | 454.1 |
| 22 | 221.0 | 143.9 | 200.0 | 241.5 | 294.0 | 381.0 |
| 23 | 191.0 | 115.8 | 158.0 | 194.5 | 247.2 | 333.4 |
| 24 | 219.0 | 80.0 | 121.8 | 155.5 | 207.5 | 291.1 |
| 25 | 159.0 | 58.9 | 91.8 | 121.0 | 166.5 | 238.3 |
| 26 | 123.0 | 43.0 | 68.0 | 95.5 | 131.2 | 199.1 |
| 27 | 115.0 | 29.0 | 51.0 | 72.0 | 104.5 | 163.2 |
| 28 | 99.0 | 17.0 | 36.0 | 54.0 | 85.0 | 139.1 |
| 29 | 82.0 | 13.0 | 26.0 | 41.0 | 62.5 | 110.2 |
| 30 | 87.0 | 8.0 | 18.8 | 32.0 | 52.0 | 93.0 |
| 31 | 58.0 | 5.0 | 13.0 | 25.0 | 39.0 | 74.1 |
| 32 | 43.0 | 5.0 | 8.8 | 18.0 | 34.0 | 62.1 |
| 33 | 61.0 | 2.0 | 7.0 | 14.0 | 25.2 | 57.0 |
| 34 | 41.0 | 1.0 | 5.0 | 12.5 | 22.0 | 47.1 |
| 35 | 41.0 | 1.0 | 4.0 | 9.5 | 18.0 | 38.1 |
| 36 | 38.0 | 0.0 | 3.0 | 7.5 | 15.0 | 40.0 |
| 37 | 23.0 | 0.0 | 2.0 | 7.0 | 14.0 | 35.1 |
| 38 | 18.0 | 0.0 | 2.0 | 6.0 | 13.0 | 31.0 |

Table S5: **Weekly calibration – Bulgaria.** Observed weekly hospitalisation counts and the 5th, 25th, 50th (median), 75th, and 95th percentiles of the simulated distributions across the 300 retained simulations.

| Week | Observed | Sim Q05 | Sim Q25 | Sim Median | Sim Q75 | Sim Q95 |
|---|---|---|---|---|---|---|
| 0 | 13.0 | 7.0 | 12.0 | 18.0 | 24.0 | 41.0 |
| 1 | 13.0 | 13.0 | 23.0 | 32.0 | 44.0 | 67.1 |
| 2 | 18.0 | 21.0 | 32.0 | 43.5 | 57.2 | 95.0 |
| 3 | 19.0 | 26.0 | 44.0 | 59.0 | 75.0 | 110.0 |
| 4 | 33.0 | 38.0 | 58.0 | 76.5 | 97.2 | 138.1 |
| 5 | 71.0 | 55.0 | 77.0 | 99.5 | 124.2 | 173.3 |
| 6 | 93.0 | 70.0 | 99.0 | 128.0 | 158.0 | 205.1 |
| 7 | 131.0 | 94.0 | 128.0 | 159.0 | 197.0 | 249.4 |
| 8 | 191.0 | 119.0 | 161.8 | 196.0 | 232.2 | 290.1 |
| 9 | 223.0 | 154.9 | 195.0 | 230.0 | 274.2 | 338.2 |
| 10 | 296.0 | 178.9 | 230.0 | 269.0 | 314.0 | 403.1 |
| 11 | 340.0 | 208.0 | 259.0 | 303.0 | 351.0 | 428.0 |
| 12 | 410.0 | 220.0 | 287.2 | 330.0 | 371.2 | 461.1 |
| 13 | 424.0 | 251.8 | 309.0 | 341.0 | 388.0 | 482.1 |
| 14 | 459.0 | 249.9 | 315.0 | 346.0 | 396.8 | 480.0 |
| 15 | 396.0 | 254.9 | 308.8 | 345.0 | 393.5 | 466.2 |
| 16 | 393.0 | 246.0 | 294.0 | 335.0 | 378.0 | 437.0 |
| 17 | 344.0 | 221.9 | 271.8 | 314.5 | 355.0 | 416.3 |
| 18 | 249.0 | 201.9 | 250.0 | 284.0 | 327.2 | 379.0 |
| 19 | 191.0 | 175.0 | 216.8 | 251.0 | 288.0 | 349.0 |
| 20 | 163.0 | 141.0 | 180.0 | 215.0 | 255.0 | 306.2 |
| 21 | 130.0 | 107.0 | 148.0 | 180.5 | 220.0 | 267.0 |
| 22 | 143.0 | 83.0 | 116.8 | 149.0 | 184.0 | 232.0 |
| 23 | 105.0 | 60.0 | 91.0 | 120.5 | 151.5 | 187.0 |
| 24 | 141.0 | 41.0 | 71.0 | 96.0 | 121.0 | 160.1 |
| 25 | 122.0 | 30.0 | 52.8 | 74.0 | 100.0 | 136.1 |
| 26 | 80.0 | 20.0 | 37.0 | 59.0 | 79.0 | 111.1 |
| 27 | 57.0 | 13.9 | 27.0 | 42.5 | 61.0 | 91.0 |
| 28 | 63.0 | 7.0 | 18.0 | 30.0 | 46.2 | 72.0 |
| 29 | 51.0 | 5.0 | 12.0 | 22.0 | 36.0 | 61.1 |
| 30 | 47.0 | 4.0 | 9.0 | 16.0 | 28.0 | 52.1 |
| 31 | 33.0 | 2.0 | 6.0 | 11.0 | 21.0 | 41.0 |
| 32 | 32.0 | 1.0 | 4.0 | 8.0 | 19.0 | 38.1 |
| 33 | 41.0 | 0.0 | 2.0 | 6.0 | 13.2 | 32.0 |
| 34 | 29.0 | 0.0 | 2.0 | 5.0 | 11.0 | 28.0 |
| 35 | 21.0 | 0.0 | 1.0 | 4.0 | 10.0 | 23.1 |
| 36 | 16.0 | 0.0 | 1.0 | 3.0 | 8.0 | 22.1 |
| 37 | 15.0 | 0.0 | 1.0 | 2.0 | 8.0 | 21.1 |
| 38 | 21.0 | 0.0 | 0.0 | 2.0 | 7.0 | 19.1 |

Table S6: **Weekly calibration – Croatia.** Observed weekly hospitalisation counts and the 5th, 25th, 50th (median), 75th, and 95th percentiles of the simulated distributions across the 300 retained simulations.

| Week | Observed | Sim Q05 | Sim Q25 | Sim Median | Sim Q75 | Sim Q95 |
|---|---|---|---|---|---|---|
| 0 | 4.0 | 11.0 | 17.0 | 24.0 | 35.0 | 48.1 |
| 1 | 7.0 | 11.0 | 18.0 | 26.0 | 36.0 | 53.0 |
| 2 | 6.0 | 12.9 | 20.0 | 28.0 | 37.0 | 61.0 |
| 3 | 14.0 | 13.0 | 23.0 | 32.5 | 44.0 | 68.1 |
| 4 | 15.0 | 18.0 | 27.0 | 39.0 | 56.2 | 86.1 |
| 5 | 26.0 | 24.0 | 36.0 | 52.0 | 68.2 | 98.0 |
| 6 | 49.0 | 28.0 | 45.0 | 60.0 | 84.0 | 119.0 |
| 7 | 68.0 | 38.0 | 57.0 | 74.0 | 103.0 | 140.2 |
| 8 | 97.0 | 47.0 | 70.0 | 92.0 | 121.2 | 160.1 |
| 9 | 117.0 | 56.0 | 84.0 | 111.0 | 141.5 | 189.1 |
| 10 | 175.0 | 69.0 | 99.0 | 126.0 | 160.2 | 209.1 |
| 11 | 203.0 | 76.0 | 113.8 | 144.0 | 177.5 | 235.1 |
| 12 | 225.0 | 90.0 | 127.0 | 160.5 | 194.0 | 256.1 |
| 13 | 224.0 | 96.0 | 139.0 | 173.0 | 204.0 | 267.3 |
| 14 | 238.0 | 107.0 | 147.0 | 179.0 | 216.0 | 282.1 |
| 15 | 231.0 | 109.0 | 149.8 | 181.0 | 215.5 | 287.2 |
| 16 | 193.0 | 112.0 | 146.8 | 176.0 | 214.0 | 282.1 |
| 17 | 190.0 | 106.0 | 143.0 | 172.0 | 207.2 | 272.0 |
| 18 | 145.0 | 101.0 | 129.8 | 160.5 | 199.2 | 261.1 |
| 19 | 113.0 | 90.0 | 119.0 | 147.0 | 180.0 | 229.3 |
| 20 | 104.0 | 76.0 | 105.0 | 131.0 | 160.5 | 214.1 |
| 21 | 92.0 | 60.0 | 88.8 | 113.0 | 143.2 | 184.0 |
| 22 | 73.0 | 47.0 | 72.0 | 96.0 | 124.2 | 158.0 |
| 23 | 66.0 | 36.0 | 58.0 | 78.5 | 103.0 | 133.2 |
| 24 | 58.0 | 26.0 | 44.0 | 64.0 | 83.0 | 114.0 |
| 25 | 52.0 | 17.0 | 34.0 | 52.0 | 68.2 | 91.0 |
| 26 | 47.0 | 12.0 | 25.0 | 38.0 | 53.0 | 71.3 |
| 27 | 37.0 | 6.0 | 18.0 | 28.0 | 42.0 | 58.0 |
| 28 | 35.0 | 5.0 | 13.0 | 22.0 | 32.0 | 49.1 |
| 29 | 27.0 | 4.0 | 8.0 | 16.0 | 25.0 | 41.0 |
| 30 | 18.0 | 2.0 | 6.0 | 11.0 | 19.0 | 34.0 |
| 31 | 17.0 | 1.0 | 3.0 | 8.0 | 14.2 | 26.1 |
| 32 | 15.0 | 0.0 | 2.8 | 5.0 | 11.0 | 22.0 |
| 33 | 13.0 | 0.0 | 1.0 | 4.0 | 8.0 | 19.1 |
| 34 | 13.0 | 0.0 | 1.0 | 3.0 | 6.0 | 15.0 |
| 35 | 13.0 | 0.0 | 1.0 | 2.0 | 6.0 | 14.1 |
| 36 | 5.0 | 0.0 | 0.0 | 1.0 | 4.0 | 13.0 |
| 37 | 8.0 | 0.0 | 0.0 | 1.0 | 3.0 | 11.0 |
| 38 | 5.0 | 0.0 | 0.0 | 1.0 | 3.0 | 9.0 |

Table S7: **Weekly calibration – Cyprus.** Observed weekly hospitalisation counts and the 5th, 25th, 50th (median), 75th, and 95th percentiles of the simulated distributions across the 300 retained simulations.

| Week | Observed | Sim Q05 | Sim Q25 | Sim Median | Sim Q75 | Sim Q95 |
|---|---|---|---|---|---|---|
| 0 | 1.0 | 0.0 | 2.0 | 3.0 | 5.0 | 10.1 |
| 1 | 2.0 | 0.0 | 2.8 | 5.0 | 8.0 | 16.0 |
| 2 | 5.0 | 1.0 | 4.0 | 6.0 | 10.0 | 18.1 |
| 3 | 4.0 | 2.0 | 5.0 | 8.0 | 12.0 | 19.0 |
| 4 | 3.0 | 3.0 | 7.0 | 10.0 | 14.0 | 23.1 |
| 5 | 9.0 | 4.0 | 9.0 | 13.0 | 18.0 | 29.0 |
| 6 | 9.0 | 7.0 | 11.0 | 17.0 | 23.0 | 32.0 |
| 7 | 14.0 | 9.0 | 15.0 | 20.0 | 27.0 | 38.0 |
| 8 | 16.0 | 9.9 | 19.0 | 25.5 | 32.0 | 47.1 |
| 9 | 37.0 | 12.0 | 23.0 | 30.0 | 38.0 | 55.0 |
| 10 | 39.0 | 15.0 | 26.0 | 34.0 | 45.2 | 63.0 |
| 11 | 45.0 | 18.0 | 31.0 | 40.0 | 50.0 | 70.0 |
| 12 | 63.0 | 18.0 | 33.0 | 42.0 | 52.0 | 72.0 |
| 13 | 50.0 | 20.0 | 35.0 | 46.0 | 55.0 | 77.1 |
| 14 | 57.0 | 24.9 | 35.0 | 45.0 | 56.0 | 78.1 |
| 15 | 66.0 | 25.0 | 36.0 | 45.0 | 53.2 | 75.1 |
| 16 | 57.0 | 21.0 | 35.0 | 43.0 | 53.0 | 68.0 |
| 17 | 43.0 | 22.0 | 33.0 | 41.0 | 50.0 | 66.1 |
| 18 | 29.0 | 20.0 | 30.8 | 38.0 | 46.0 | 65.1 |
| 19 | 25.0 | 16.0 | 25.0 | 33.0 | 41.0 | 56.0 |
| 20 | 17.0 | 13.0 | 22.0 | 28.0 | 36.0 | 48.0 |
| 21 | 16.0 | 11.0 | 17.8 | 23.0 | 31.0 | 43.1 |
| 22 | 15.0 | 8.0 | 14.0 | 20.0 | 27.0 | 35.1 |
| 23 | 18.0 | 6.0 | 11.0 | 16.0 | 22.0 | 31.0 |
| 24 | 17.0 | 5.0 | 9.0 | 12.0 | 18.0 | 26.0 |
| 25 | 17.0 | 2.0 | 6.0 | 10.0 | 14.0 | 21.0 |
| 26 | 10.0 | 2.0 | 4.0 | 7.0 | 11.0 | 17.1 |
| 27 | 6.0 | 1.0 | 3.0 | 5.0 | 9.0 | 14.0 |
| 28 | 6.0 | 0.0 | 2.0 | 4.0 | 7.0 | 13.0 |
| 29 | 9.0 | 0.0 | 1.0 | 3.0 | 6.0 | 10.1 |
| 30 | 11.0 | 0.0 | 1.0 | 2.0 | 4.0 | 10.0 |
| 31 | 5.0 | 0.0 | 0.0 | 2.0 | 4.0 | 9.0 |
| 32 | 9.0 | 0.0 | 0.0 | 1.0 | 3.0 | 7.0 |
| 33 | 4.0 | 0.0 | 0.0 | 1.0 | 2.0 | 5.0 |
| 34 | 5.0 | 0.0 | 0.0 | 0.0 | 2.0 | 5.0 |
| 35 | 1.0 | 0.0 | 0.0 | 0.0 | 1.0 | 5.0 |
| 36 | 3.0 | 0.0 | 0.0 | 0.0 | 1.0 | 5.0 |
| 37 | 2.0 | 0.0 | 0.0 | 0.0 | 1.0 | 4.0 |
| 38 | 2.0 | 0.0 | 0.0 | 0.0 | 1.0 | 4.0 |

Table S8: **Weekly calibration – Czechia.** Observed weekly hospitalisation counts and the 5th, 25th, 50th (median), 75th, and 95th percentiles of the simulated distributions across the 300 retained simulations.

| Week | Observed | Sim Q05 | Sim Q25 | Sim Median | Sim Q75 | Sim Q95 |
|---|---|---|---|---|---|---|
| 0 | 16.0 | 10.0 | 21.0 | 32.0 | 48.5 | 73.1 |
| 1 | 17.0 | 20.0 | 38.0 | 55.0 | 79.2 | 120.2 |
| 2 | 20.0 | 29.0 | 51.0 | 77.5 | 104.0 | 157.0 |
| 3 | 41.0 | 44.0 | 70.0 | 98.0 | 132.0 | 186.1 |
| 4 | 68.0 | 53.0 | 90.0 | 126.5 | 169.0 | 233.1 |
| 5 | 86.0 | 71.0 | 116.8 | 161.0 | 211.0 | 276.1 |
| 6 | 132.0 | 101.0 | 155.5 | 208.0 | 258.0 | 330.1 |
| 7 | 179.0 | 134.9 | 190.8 | 254.5 | 324.5 | 383.1 |
| 8 | 257.0 | 168.7 | 236.8 | 309.0 | 380.0 | 460.0 |
| 9 | 362.0 | 208.0 | 285.8 | 372.0 | 441.2 | 523.0 |
| 10 | 496.0 | 263.9 | 341.0 | 426.5 | 487.0 | 595.0 |
| 11 | 533.0 | 312.8 | 384.8 | 465.5 | 525.0 | 635.0 |
| 12 | 605.0 | 352.8 | 417.0 | 492.0 | 562.2 | 651.0 |
| 13 | 629.0 | 375.9 | 448.0 | 513.5 | 579.0 | 697.1 |
| 14 | 652.0 | 384.9 | 455.0 | 515.5 | 578.0 | 686.2 |
| 15 | 628.0 | 391.9 | 447.8 | 507.5 | 568.2 | 685.3 |
| 16 | 573.0 | 357.9 | 435.0 | 490.5 | 547.2 | 650.3 |
| 17 | 515.0 | 345.0 | 406.0 | 456.0 | 520.0 | 620.5 |
| 18 | 409.0 | 303.9 | 365.8 | 413.0 | 485.0 | 584.1 |
| 19 | 308.0 | 259.0 | 323.0 | 372.0 | 435.2 | 530.0 |
| 20 | 272.0 | 209.8 | 278.0 | 332.0 | 388.2 | 469.0 |
| 21 | 220.0 | 155.9 | 224.5 | 280.5 | 343.5 | 423.1 |
| 22 | 193.0 | 118.9 | 188.0 | 235.5 | 297.0 | 358.1 |
| 23 | 156.0 | 89.6 | 147.0 | 194.0 | 245.2 | 312.1 |
| 24 | 170.0 | 64.8 | 113.8 | 153.0 | 206.2 | 272.0 |
| 25 | 161.0 | 49.0 | 88.0 | 127.5 | 168.0 | 225.1 |
| 26 | 138.0 | 32.0 | 62.0 | 95.0 | 132.0 | 193.1 |
| 27 | 108.0 | 20.0 | 44.8 | 73.5 | 108.0 | 165.1 |
| 28 | 85.0 | 15.0 | 34.0 | 54.5 | 87.0 | 138.1 |
| 29 | 109.0 | 9.0 | 22.0 | 39.0 | 67.0 | 118.2 |
| 30 | 81.0 | 7.0 | 16.0 | 31.0 | 56.0 | 102.0 |
| 31 | 54.0 | 4.0 | 11.8 | 23.0 | 44.5 | 86.1 |
| 32 | 47.0 | 3.0 | 8.8 | 17.0 | 37.2 | 78.1 |
| 33 | 37.0 | 2.0 | 5.8 | 13.5 | 31.2 | 67.1 |
| 34 | 51.0 | 1.0 | 4.0 | 10.0 | 26.0 | 58.1 |
| 35 | 31.0 | 0.0 | 3.0 | 8.0 | 24.0 | 54.0 |
| 36 | 38.0 | 0.0 | 2.0 | 6.0 | 19.2 | 53.1 |
| 37 | 39.0 | 0.0 | 2.0 | 5.0 | 18.0 | 49.2 |
| 38 | 19.0 | 0.0 | 1.0 | 5.0 | 15.0 | 49.0 |

Table S9: **Weekly calibration – Denmark.** Observed weekly hospitalisation counts and the 5th, 25th, 50th (median), 75th, and 95th percentiles of the simulated distributions across the 300 retained simulations.

| Week | Observed | Sim Q05 | Sim Q25 | Sim Median | Sim Q75 | Sim Q95 |
|---|---|---|---|---|---|---|
| 0 | 9.0 | 8.0 | 13.0 | 20.0 | 28.0 | 42.1 |
| 1 | 5.0 | 14.9 | 23.0 | 32.5 | 43.0 | 62.0 |
| 2 | 18.0 | 20.0 | 31.0 | 43.5 | 57.0 | 82.0 |
| 3 | 18.0 | 29.0 | 43.0 | 57.0 | 73.2 | 102.0 |
| 4 | 28.0 | 36.0 | 55.8 | 73.5 | 95.0 | 122.0 |
| 5 | 51.0 | 50.9 | 72.8 | 96.0 | 119.2 | 152.2 |
| 6 | 68.0 | 67.0 | 92.8 | 123.0 | 149.8 | 189.1 |
| 7 | 92.0 | 87.9 | 116.0 | 151.0 | 188.0 | 226.0 |
| 8 | 136.0 | 108.7 | 145.8 | 181.5 | 220.2 | 270.1 |
| 9 | 194.0 | 131.9 | 176.8 | 214.0 | 252.0 | 314.0 |
| 10 | 277.0 | 150.9 | 201.0 | 242.5 | 285.0 | 355.1 |
| 11 | 323.0 | 169.9 | 223.0 | 263.5 | 309.5 | 377.1 |
| 12 | 382.0 | 198.9 | 245.0 | 283.0 | 331.2 | 397.1 |
| 13 | 339.0 | 210.9 | 261.0 | 299.0 | 340.0 | 397.2 |
| 14 | 366.0 | 219.9 | 265.0 | 302.0 | 338.0 | 411.1 |
| 15 | 363.0 | 216.0 | 259.0 | 292.5 | 326.2 | 387.1 |
| 16 | 320.0 | 204.0 | 246.5 | 282.0 | 316.0 | 376.1 |
| 17 | 305.0 | 184.9 | 221.0 | 256.0 | 292.0 | 346.0 |
| 18 | 228.0 | 168.0 | 200.0 | 234.5 | 269.2 | 319.1 |
| 19 | 173.0 | 137.0 | 172.0 | 206.0 | 241.2 | 291.1 |
| 20 | 150.0 | 110.0 | 144.0 | 177.0 | 209.2 | 260.1 |
| 21 | 119.0 | 82.0 | 120.8 | 152.0 | 182.0 | 231.3 |
| 22 | 104.0 | 60.0 | 98.0 | 126.0 | 151.2 | 201.1 |
| 23 | 88.0 | 43.0 | 76.0 | 104.5 | 128.2 | 170.2 |
| 24 | 115.0 | 34.0 | 58.5 | 80.0 | 104.0 | 144.2 |
| 25 | 85.0 | 22.9 | 43.0 | 64.5 | 87.2 | 121.2 |
| 26 | 77.0 | 16.0 | 31.0 | 50.0 | 70.2 | 101.0 |
| 27 | 70.0 | 10.0 | 24.0 | 38.5 | 53.0 | 87.1 |
| 28 | 52.0 | 6.0 | 17.0 | 28.0 | 43.2 | 67.1 |
| 29 | 56.0 | 4.0 | 11.0 | 20.0 | 33.2 | 60.1 |
| 30 | 47.0 | 3.0 | 8.0 | 18.0 | 26.2 | 51.0 |
| 31 | 30.0 | 2.0 | 5.0 | 11.0 | 22.0 | 43.1 |
| 32 | 42.0 | 1.0 | 4.0 | 9.0 | 17.0 | 37.1 |
| 33 | 24.0 | 0.0 | 3.0 | 6.0 | 14.2 | 34.0 |
| 34 | 19.0 | 0.0 | 2.0 | 5.0 | 12.0 | 27.1 |
| 35 | 18.0 | 0.0 | 1.0 | 4.0 | 11.0 | 24.0 |
| 36 | 18.0 | 0.0 | 1.0 | 3.0 | 9.0 | 22.1 |
| 37 | 14.0 | 0.0 | 1.0 | 3.0 | 8.0 | 21.0 |
| 38 | 15.0 | 0.0 | 1.0 | 2.0 | 7.2 | 19.0 |

Table S10: **Weekly calibration – Estonia.** Observed weekly hospitalisation counts and the 5th, 25th, 50th (median), 75th, and 95th percentiles of the simulated distributions across the 300 retained simulations.

| **Week** | **Observed** | **Sim Q05** | **Sim Q25** | **Sim Median** | **Sim Q75** | **Sim Q95** |
|---|---|---|---|---|---|---|
| 0 | 2.0 | 1.0 | 2.0 | 4.0 | 8.0 | 13.0 |
| 1 | 2.0 | 1.0 | 4.0 | 7.0 | 12.0 | 22.0 |
| 2 | 5.0 | 2.0 | 6.0 | 10.0 | 15.0 | 26.0 |
| 3 | 4.0 | 4.0 | 7.0 | 11.0 | 18.0 | 28.0 |
| 4 | 3.0 | 5.0 | 10.0 | 14.0 | 21.0 | 32.0 |
| 5 | 11.0 | 8.0 | 14.0 | 19.0 | 26.0 | 38.0 |
| 6 | 18.0 | 9.0 | 17.0 | 24.0 | 32.0 | 43.0 |
| 7 | 25.0 | 13.0 | 21.0 | 28.0 | 36.0 | 50.0 |
| 8 | 29.0 | 15.0 | 25.0 | 33.0 | 43.0 | 56.1 |
| 9 | 38.0 | 19.0 | 28.0 | 39.0 | 49.0 | 61.1 |
| 10 | 58.0 | 23.0 | 35.0 | 44.0 | 54.0 | 71.0 |
| 11 | 60.0 | 26.0 | 39.0 | 48.0 | 59.0 | 77.1 |
| 12 | 65.0 | 28.0 | 40.0 | 52.0 | 64.2 | 83.0 |
| 13 | 80.0 | 31.0 | 44.8 | 54.0 | 68.0 | 84.1 |
| 14 | 80.0 | 33.0 | 46.0 | 56.0 | 69.0 | 88.1 |
| 15 | 74.0 | 31.0 | 45.0 | 58.0 | 68.0 | 87.1 |
| 16 | 58.0 | 32.0 | 44.0 | 55.5 | 67.0 | 86.1 |
| 17 | 63.0 | 31.0 | 42.0 | 54.0 | 65.0 | 81.1 |
| 18 | 43.0 | 26.0 | 39.0 | 50.0 | 59.0 | 78.0 |
| 19 | 32.0 | 25.0 | 35.0 | 45.0 | 57.0 | 73.1 |
| 20 | 30.0 | 21.0 | 31.0 | 41.0 | 51.0 | 64.0 |
| 21 | 27.0 | 16.0 | 26.0 | 36.0 | 44.0 | 57.1 |
| 22 | 30.0 | 12.0 | 22.0 | 29.0 | 38.0 | 52.0 |
| 23 | 23.0 | 9.0 | 16.0 | 24.0 | 32.0 | 44.1 |
| 24 | 27.0 | 7.0 | 13.0 | 20.0 | 28.0 | 39.1 |
| 25 | 22.0 | 5.0 | 9.0 | 16.0 | 23.0 | 33.0 |
| 26 | 22.0 | 3.0 | 8.0 | 12.0 | 18.2 | 28.0 |
| 27 | 9.0 | 2.0 | 5.0 | 10.0 | 16.0 | 23.0 |
| 28 | 13.0 | 1.0 | 4.0 | 7.5 | 11.0 | 21.1 |
| 29 | 4.0 | 0.0 | 2.0 | 5.0 | 10.0 | 17.1 |
| 30 | 12.0 | 0.0 | 1.0 | 4.0 | 8.2 | 16.0 |
| 31 | 8.0 | 0.0 | 1.0 | 3.0 | 6.0 | 13.0 |
| 32 | 10.0 | 0.0 | 1.0 | 2.0 | 5.0 | 11.0 |
| 33 | 3.0 | 0.0 | 0.0 | 2.0 | 4.0 | 10.0 |
| 34 | 5.0 | 0.0 | 0.0 | 1.0 | 3.0 | 8.0 |
| 35 | 3.0 | 0.0 | 0.0 | 1.0 | 3.0 | 7.0 |
| 36 | 3.0 | 0.0 | 0.0 | 1.0 | 2.0 | 6.0 |
| 37 | 1.0 | 0.0 | 0.0 | 0.0 | 2.0 | 7.0 |
| 38 | 3.0 | 0.0 | 0.0 | 0.0 | 2.0 | 6.0 |

Table S11: **Weekly calibration – Finland.** Observed weekly hospitalisation counts and the 5th, 25th, 50th (median), 75th, and 95th percentiles of the simulated distributions across the 300 retained simulations.

| Week | Observed | Sim Q05 | Sim Q25 | Sim Median | Sim Q75 | Sim Q95 |
|---|---|---|---|---|---|---|
| 0 | 8.0 | 6.0 | 12.0 | 18.0 | 26.0 | 38.1 |
| 1 | 7.0 | 11.0 | 19.0 | 30.0 | 43.0 | 63.1 |
| 2 | 9.0 | 14.0 | 25.0 | 38.0 | 56.0 | 80.1 |
| 3 | 17.0 | 19.0 | 29.0 | 47.0 | 66.0 | 95.0 |
| 4 | 21.0 | 26.0 | 38.0 | 59.0 | 82.0 | 115.0 |
| 5 | 26.0 | 28.0 | 50.0 | 72.0 | 98.2 | 132.1 |
| 6 | 49.0 | 35.0 | 62.0 | 87.0 | 115.0 | 156.0 |
| 7 | 92.0 | 53.0 | 78.8 | 103.0 | 136.0 | 178.1 |
| 8 | 95.0 | 64.0 | 97.0 | 124.0 | 155.2 | 204.0 |
| 9 | 156.0 | 78.0 | 112.8 | 144.0 | 172.0 | 226.1 |
| 10 | 189.0 | 90.9 | 132.8 | 159.5 | 191.0 | 256.2 |
| 11 | 234.0 | 109.8 | 147.0 | 174.0 | 209.2 | 271.1 |
| 12 | 273.0 | 123.9 | 163.0 | 188.0 | 220.0 | 276.1 |
| 13 | 268.0 | 135.0 | 171.0 | 196.0 | 230.0 | 285.1 |
| 14 | 299.0 | 143.9 | 174.8 | 199.0 | 230.0 | 290.1 |
| 15 | 250.0 | 143.9 | 176.0 | 202.0 | 230.2 | 287.0 |
| 16 | 241.0 | 142.0 | 173.0 | 195.0 | 227.0 | 273.2 |
| 17 | 218.0 | 134.0 | 159.8 | 190.0 | 216.0 | 265.2 |
| 18 | 164.0 | 120.0 | 148.0 | 175.0 | 205.2 | 246.7 |
| 19 | 138.0 | 108.0 | 134.0 | 157.0 | 190.2 | 241.1 |
| 20 | 112.0 | 93.0 | 113.0 | 147.0 | 174.0 | 217.0 |
| 21 | 111.0 | 71.0 | 97.8 | 128.0 | 156.0 | 192.0 |
| 22 | 103.0 | 56.9 | 83.0 | 109.5 | 136.0 | 170.1 |
| 23 | 77.0 | 42.0 | 67.8 | 92.0 | 115.0 | 145.1 |
| 24 | 72.0 | 32.0 | 54.0 | 76.0 | 96.0 | 122.0 |
| 25 | 63.0 | 22.0 | 43.0 | 61.0 | 80.0 | 107.0 |
| 26 | 40.0 | 15.9 | 34.0 | 50.0 | 67.2 | 93.0 |
| 27 | 44.0 | 10.0 | 23.8 | 38.0 | 52.0 | 75.0 |
| 28 | 34.0 | 7.0 | 19.0 | 30.0 | 45.0 | 65.0 |
| 29 | 41.0 | 5.0 | 13.0 | 22.5 | 37.0 | 55.1 |
| 30 | 31.0 | 3.0 | 9.0 | 17.0 | 28.2 | 46.0 |
| 31 | 27.0 | 1.0 | 6.8 | 13.0 | 24.0 | 40.1 |
| 32 | 22.0 | 1.0 | 4.8 | 9.5 | 20.0 | 35.1 |
| 33 | 18.0 | 1.0 | 3.0 | 8.0 | 16.0 | 33.0 |
| 34 | 12.0 | 0.0 | 2.0 | 6.0 | 13.2 | 27.1 |
| 35 | 12.0 | 0.0 | 1.0 | 5.0 | 12.0 | 25.1 |
| 36 | 11.0 | 0.0 | 1.0 | 4.0 | 10.0 | 23.0 |
| 37 | 5.0 | 0.0 | 1.0 | 3.0 | 9.2 | 22.0 |
| 38 | 5.0 | 0.0 | 0.0 | 2.0 | 8.0 | 21.1 |

Table S12: **Weekly calibration – France.** Observed weekly hospitalisation counts and the 5th, 25th, 50th (median), 75th, and 95th percentiles of the simulated distributions across the 300 retained simulations.

| Week | Observed | Sim Q05 | Sim Q25 | Sim Median | Sim Q75 | Sim Q95 |
|---|---|---|---|---|---|---|
| 0 | 126.0 | 65.0 | 104.8 | 148.0 | 209.0 | 413.0 |
| 1 | 125.0 | 109.8 | 183.8 | 259.5 | 377.2 | 655.6 |
| 2 | 185.0 | 167.9 | 263.8 | 372.5 | 510.5 | 881.2 |
| 3 | 263.0 | 248.1 | 368.8 | 508.5 | 699.5 | 1,097.4 |
| 4 | 393.0 | 354.9 | 518.2 | 679.0 | 949.5 | 1,391.0 |
| 5 | 622.0 | 499.8 | 712.8 | 918.5 | 1,259.0 | 1,727.0 |
| 6 | 888.0 | 693.7 | 962.5 | 1,242.0 | 1,652.0 | 2,202.4 |
| 7 | 1,356.0 | 981.6 | 1,276.2 | 1,615.0 | 2,103.5 | 2,646.6 |
| 8 | 1,882.0 | 1,295.2 | 1,674.0 | 2,079.5 | 2,554.5 | 3,227.3 |
| 9 | 2,668.0 | 1,692.3 | 2,115.8 | 2,527.5 | 3,026.0 | 3,771.7 |
| 10 | 3,514.0 | 2,128.6 | 2,572.8 | 3,018.0 | 3,526.0 | 4,312.9 |
| 11 | 4,033.0 | 2,548.1 | 3,061.8 | 3,479.5 | 3,980.5 | 4,696.4 |
| 12 | 4,899.0 | 2,912.9 | 3,394.8 | 3,803.5 | 4,290.5 | 4,984.1 |
| 13 | 4,744.0 | 3,140.3 | 3,619.5 | 4,036.5 | 4,518.2 | 5,125.8 |
| 14 | 5,012.0 | 3,253.4 | 3,728.2 | 4,104.0 | 4,622.5 | 5,158.2 |
| 15 | 4,596.0 | 3,256.8 | 3,672.5 | 4,042.5 | 4,537.8 | 5,151.1 |
| 16 | 4,231.0 | 3,058.8 | 3,490.5 | 3,927.0 | 4,302.0 | 4,973.8 |
| 17 | 3,673.0 | 2,706.8 | 3,247.0 | 3,659.5 | 4,042.5 | 4,674.1 |
| 18 | 2,883.0 | 2,392.3 | 2,925.5 | 3,274.0 | 3,716.0 | 4,249.5 |
| 19 | 2,274.0 | 1,992.8 | 2,525.5 | 2,914.5 | 3,306.2 | 3,930.1 |
| 20 | 2,074.0 | 1,675.8 | 2,151.0 | 2,513.5 | 2,904.5 | 3,414.8 |
| 21 | 1,855.0 | 1,296.8 | 1,765.2 | 2,109.0 | 2,478.0 | 2,993.2 |
| 22 | 1,513.0 | 970.6 | 1,429.5 | 1,743.0 | 2,093.0 | 2,574.6 |
| 23 | 1,355.0 | 716.9 | 1,117.2 | 1,398.0 | 1,729.5 | 2,193.4 |
| 24 | 1,317.0 | 510.9 | 843.2 | 1,103.5 | 1,400.8 | 1,836.3 |
| 25 | 1,148.0 | 343.9 | 630.5 | 866.0 | 1,132.8 | 1,534.4 |
| 26 | 883.0 | 245.8 | 471.0 | 662.0 | 893.2 | 1,257.5 |
| 27 | 823.0 | 175.8 | 333.5 | 500.5 | 709.0 | 1,088.3 |
| 28 | 662.0 | 115.8 | 227.8 | 380.0 | 552.8 | 884.7 |
| 29 | 639.0 | 78.8 | 160.5 | 276.5 | 442.5 | 714.6 |
| 30 | 503.0 | 51.0 | 110.0 | 203.0 | 361.0 | 620.5 |
| 31 | 438.0 | 32.9 | 73.8 | 151.5 | 287.0 | 509.5 |
| 32 | 419.0 | 22.0 | 55.2 | 116.5 | 228.2 | 475.3 |
| 33 | 295.0 | 12.9 | 38.0 | 89.5 | 191.2 | 395.4 |
| 34 | 317.0 | 9.0 | 27.0 | 68.0 | 152.5 | 349.1 |
| 35 | 261.0 | 6.0 | 21.0 | 58.0 | 130.5 | 292.3 |
| 36 | 241.0 | 5.0 | 15.8 | 44.0 | 113.2 | 266.5 |
| 37 | 204.0 | 2.0 | 12.8 | 38.5 | 99.0 | 260.7 |
| 38 | 164.0 | 2.0 | 10.0 | 33.0 | 90.0 | 275.3 |

Table S13: **Weekly calibration – Germany.** Observed weekly hospitalisation counts and the 5th, 25th, 50th (median), 75th, and 95th percentiles of the simulated distributions across the 300 retained simulations.

| Week | Observed | Sim Q05 | Sim Q25 | Sim Median | Sim Q75 | Sim Q95 |
|---|---|---|---|---|---|---|
| 0 | 122.0 | 52.0 | 72.8 | 109.0 | 194.5 | 391.2 |
| 1 | 122.0 | 100.0 | 136.8 | 199.0 | 349.5 | 686.0 |
| 2 | 170.0 | 146.9 | 210.8 | 289.0 | 498.8 | 911.2 |
| 3 | 258.0 | 223.9 | 309.0 | 424.5 | 675.2 | 1,179.1 |
| 4 | 427.0 | 330.9 | 443.8 | 583.0 | 914.2 | 1,472.7 |
| 5 | 623.0 | 495.9 | 646.8 | 826.5 | 1,179.5 | 1,798.8 |
| 6 | 906.0 | 676.9 | 912.8 | 1,154.5 | 1,525.5 | 2,156.6 |
| 7 | 1,395.0 | 963.0 | 1,239.2 | 1,545.0 | 1,966.8 | 2,547.8 |
| 8 | 1,876.0 | 1,288.7 | 1,670.2 | 1,999.5 | 2,425.2 | 3,025.0 |
| 9 | 2,574.0 | 1,695.7 | 2,096.5 | 2,502.5 | 2,906.5 | 3,578.3 |
| 10 | 3,389.0 | 2,155.2 | 2,569.0 | 2,979.0 | 3,397.5 | 4,120.8 |
| 11 | 3,979.0 | 2,556.0 | 3,029.8 | 3,402.0 | 3,858.0 | 4,480.2 |
| 12 | 4,602.0 | 2,920.8 | 3,346.8 | 3,735.0 | 4,234.5 | 4,960.3 |
| 13 | 4,815.0 | 3,092.2 | 3,574.8 | 3,976.0 | 4,425.2 | 5,128.4 |
| 14 | 4,919.0 | 3,268.7 | 3,672.0 | 4,085.0 | 4,522.0 | 5,137.4 |
| 15 | 4,542.0 | 3,175.1 | 3,634.5 | 4,050.5 | 4,504.2 | 5,134.0 |
| 16 | 4,081.0 | 2,988.1 | 3,474.8 | 3,845.5 | 4,300.2 | 4,927.3 |
| 17 | 3,593.0 | 2,736.8 | 3,221.2 | 3,585.5 | 4,045.8 | 4,639.7 |
| 18 | 2,886.0 | 2,440.7 | 2,872.8 | 3,238.5 | 3,653.0 | 4,256.9 |
| 19 | 2,211.0 | 2,083.6 | 2,480.0 | 2,865.5 | 3,257.8 | 3,860.5 |
| 20 | 2,038.0 | 1,675.1 | 2,129.8 | 2,443.5 | 2,849.8 | 3,442.7 |
| 21 | 1,701.0 | 1,302.7 | 1,730.8 | 2,053.0 | 2,452.5 | 2,966.7 |
| 22 | 1,489.0 | 1,011.7 | 1,379.2 | 1,657.5 | 2,070.0 | 2,525.5 |
| 23 | 1,281.0 | 736.2 | 1,059.5 | 1,341.0 | 1,723.2 | 2,170.2 |
| 24 | 1,245.0 | 519.5 | 791.8 | 1,045.5 | 1,402.5 | 1,816.2 |
| 25 | 1,183.0 | 362.9 | 583.8 | 822.0 | 1,119.0 | 1,527.6 |
| 26 | 929.0 | 244.9 | 432.2 | 614.5 | 905.5 | 1,253.3 |
| 27 | 836.0 | 163.0 | 309.2 | 468.0 | 718.0 | 1,036.5 |
| 28 | 661.0 | 105.0 | 220.5 | 341.5 | 567.2 | 890.2 |
| 29 | 587.0 | 71.0 | 148.0 | 256.0 | 451.0 | 748.2 |
| 30 | 504.0 | 48.0 | 102.8 | 193.5 | 347.5 | 624.5 |
| 31 | 491.0 | 33.0 | 70.8 | 141.5 | 281.2 | 545.0 |
| 32 | 389.0 | 20.0 | 51.8 | 110.0 | 224.5 | 458.4 |
| 33 | 318.0 | 14.0 | 34.0 | 84.0 | 180.0 | 400.1 |
| 34 | 320.0 | 9.0 | 22.0 | 67.0 | 149.0 | 331.5 |
| 35 | 241.0 | 7.0 | 19.0 | 51.0 | 124.2 | 299.1 |
| 36 | 207.0 | 4.0 | 15.0 | 44.5 | 108.0 | 266.2 |
| 37 | 177.0 | 3.0 | 11.0 | 37.5 | 92.0 | 251.0 |
| 38 | 154.0 | 2.0 | 9.0 | 29.0 | 85.2 | 235.3 |

Table S14: **Weekly calibration – Greece.** Observed weekly hospitalisation counts and the 5th, 25th, 50th (median), 75th, and 95th percentiles of the simulated distributions across the 300 retained simulations.

| Week | Observed | Sim Q05 | Sim Q25 | Sim Median | Sim Q75 | Sim Q95 |
|---|---|---|---|---|---|---|
| 0 | 15.0 | 4.0 | 14.0 | 23.0 | 34.0 | 54.0 |
| 1 | 14.0 | 8.0 | 26.0 | 43.0 | 59.0 | 90.1 |
| 2 | 26.0 | 16.0 | 40.0 | 61.5 | 82.0 | 125.2 |
| 3 | 40.0 | 29.0 | 54.8 | 82.0 | 106.0 | 151.1 |
| 4 | 50.0 | 41.0 | 74.0 | 108.5 | 137.0 | 197.1 |
| 5 | 75.0 | 70.9 | 105.0 | 140.0 | 181.2 | 237.0 |
| 6 | 128.0 | 99.0 | 138.0 | 185.0 | 232.0 | 287.2 |
| 7 | 195.0 | 138.0 | 179.5 | 236.0 | 283.0 | 342.1 |
| 8 | 242.0 | 171.9 | 228.0 | 293.0 | 339.0 | 414.0 |
| 9 | 330.0 | 218.9 | 285.8 | 350.5 | 402.0 | 472.1 |
| 10 | 421.0 | 256.9 | 341.5 | 399.5 | 461.2 | 528.0 |
| 11 | 538.0 | 310.9 | 387.8 | 444.5 | 513.0 | 600.3 |
| 12 | 605.0 | 340.9 | 418.8 | 481.5 | 547.5 | 656.2 |
| 13 | 623.0 | 375.8 | 450.0 | 503.5 | 568.2 | 687.0 |
| 14 | 632.0 | 385.9 | 459.0 | 516.0 | 576.0 | 703.2 |
| 15 | 562.0 | 381.0 | 450.0 | 505.5 | 571.0 | 694.0 |
| 16 | 492.0 | 370.9 | 431.0 | 481.5 | 558.2 | 673.2 |
| 17 | 450.0 | 342.0 | 403.5 | 446.5 | 511.0 | 621.5 |
| 18 | 323.0 | 309.9 | 365.8 | 408.0 | 461.0 | 566.2 |
| 19 | 296.0 | 259.0 | 315.0 | 361.0 | 413.5 | 503.1 |
| 20 | 265.0 | 214.9 | 267.0 | 311.5 | 362.2 | 441.3 |
| 21 | 228.0 | 169.0 | 216.8 | 259.5 | 308.0 | 376.1 |
| 22 | 189.0 | 121.9 | 166.0 | 215.0 | 261.2 | 316.2 |
| 23 | 178.0 | 94.0 | 131.5 | 172.5 | 216.2 | 267.2 |
| 24 | 133.0 | 68.0 | 98.0 | 133.5 | 172.0 | 220.0 |
| 25 | 153.0 | 45.0 | 70.0 | 104.0 | 139.0 | 186.1 |
| 26 | 113.0 | 31.0 | 50.0 | 79.5 | 106.2 | 161.1 |
| 27 | 107.0 | 20.0 | 34.0 | 55.5 | 83.2 | 133.1 |
| 28 | 100.0 | 13.0 | 24.0 | 39.0 | 64.2 | 103.0 |
| 29 | 87.0 | 8.0 | 16.0 | 32.0 | 52.2 | 84.1 |
| 30 | 61.0 | 5.0 | 11.0 | 22.0 | 40.0 | 81.1 |
| 31 | 55.0 | 2.0 | 7.0 | 16.0 | 32.0 | 65.1 |
| 32 | 35.0 | 1.0 | 6.0 | 11.5 | 25.0 | 52.0 |
| 33 | 41.0 | 0.0 | 3.0 | 8.0 | 20.0 | 46.1 |
| 34 | 44.0 | 0.0 | 2.0 | 6.0 | 16.2 | 43.0 |
| 35 | 32.0 | 0.0 | 2.0 | 5.0 | 14.0 | 35.0 |
| 36 | 32.0 | 0.0 | 1.0 | 4.0 | 13.0 | 30.1 |
| 37 | 19.0 | 0.0 | 1.0 | 3.0 | 10.2 | 32.1 |
| 38 | 27.0 | 0.0 | 1.0 | 3.0 | 10.0 | 30.0 |

Table S15: **Weekly calibration – Hungary.** Observed weekly hospitalisation counts and the 5th, 25th, 50th (median), 75th, and 95th percentiles of the simulated distributions across the 300 retained simulations.

| Week | Observed | Sim Q05 | Sim Q25 | Sim Median | Sim Q75 | Sim Q95 |
|---|---|---|---|---|---|---|
| 0 | 5.0 | 6.0 | 13.0 | 23.0 | 32.2 | 49.0 |
| 1 | 17.0 | 13.0 | 25.8 | 40.0 | 57.0 | 81.3 |
| 2 | 14.0 | 21.0 | 37.0 | 54.0 | 78.0 | 109.0 |
| 3 | 29.0 | 27.0 | 53.0 | 74.0 | 100.2 | 133.1 |
| 4 | 50.0 | 45.0 | 71.0 | 101.0 | 129.0 | 170.2 |
| 5 | 89.0 | 61.0 | 102.0 | 133.0 | 166.8 | 214.1 |
| 6 | 106.0 | 87.0 | 132.0 | 172.0 | 213.0 | 269.1 |
| 7 | 145.0 | 118.0 | 172.0 | 222.0 | 263.0 | 333.1 |
| 8 | 213.0 | 158.9 | 222.8 | 273.5 | 315.5 | 403.1 |
| 9 | 334.0 | 205.9 | 275.0 | 330.0 | 381.2 | 466.1 |
| 10 | 444.0 | 263.9 | 327.0 | 379.0 | 439.2 | 534.4 |
| 11 | 487.0 | 315.9 | 374.8 | 430.0 | 493.0 | 602.2 |
| 12 | 592.0 | 345.0 | 411.8 | 466.5 | 533.0 | 638.0 |
| 13 | 578.0 | 369.9 | 423.0 | 492.0 | 547.2 | 657.1 |
| 14 | 617.0 | 389.0 | 444.0 | 493.0 | 552.2 | 669.1 |
| 15 | 597.0 | 383.6 | 435.0 | 486.5 | 548.2 | 639.2 |
| 16 | 517.0 | 356.0 | 418.8 | 468.5 | 529.0 | 616.5 |
| 17 | 441.0 | 325.9 | 385.0 | 432.5 | 487.5 | 578.5 |
| 18 | 332.0 | 278.0 | 344.0 | 396.0 | 441.0 | 524.0 |
| 19 | 273.0 | 244.8 | 298.8 | 337.0 | 388.2 | 471.0 |
| 20 | 240.0 | 197.9 | 247.0 | 293.0 | 344.0 | 419.0 |
| 21 | 216.0 | 150.9 | 201.2 | 247.0 | 293.0 | 358.0 |
| 22 | 166.0 | 119.0 | 156.0 | 203.5 | 249.2 | 307.0 |
| 23 | 150.0 | 83.9 | 124.0 | 162.0 | 206.5 | 259.1 |
| 24 | 165.0 | 60.0 | 95.0 | 127.0 | 166.2 | 218.0 |
| 25 | 125.0 | 38.9 | 67.8 | 101.0 | 140.0 | 178.2 |
| 26 | 96.0 | 28.0 | 50.0 | 75.0 | 111.0 | 149.1 |
| 27 | 80.0 | 18.0 | 34.8 | 56.0 | 91.0 | 126.0 |
| 28 | 88.0 | 12.0 | 25.0 | 44.0 | 74.0 | 104.0 |
| 29 | 72.0 | 7.0 | 17.0 | 31.0 | 56.2 | 90.0 |
| 30 | 66.0 | 5.0 | 11.0 | 22.0 | 44.2 | 71.1 |
| 31 | 66.0 | 3.0 | 8.0 | 17.0 | 34.0 | 64.2 |
| 32 | 55.0 | 1.0 | 5.0 | 13.0 | 29.0 | 51.1 |
| 33 | 44.0 | 1.0 | 3.0 | 10.0 | 23.2 | 48.1 |
| 34 | 34.0 | 0.0 | 3.0 | 7.0 | 20.0 | 43.1 |
| 35 | 31.0 | 0.0 | 2.0 | 6.0 | 17.2 | 42.1 |
| 36 | 27.0 | 0.0 | 1.0 | 4.0 | 15.0 | 36.2 |
| 37 | 25.0 | 0.0 | 1.0 | 4.0 | 12.2 | 31.2 |
| 38 | 18.0 | 0.0 | 1.0 | 3.5 | 12.0 | 33.1 |

Table S16: **Weekly calibration – Ireland.** Observed weekly hospitalisation counts and the 5th, 25th, 50th (median), 75th, and 95th percentiles of the simulated distributions across the 300 retained simulations.

| Week | Observed | Sim Q05 | Sim Q25 | Sim Median | Sim Q75 | Sim Q95 |
|---|---|---|---|---|---|---|
| 0 | 8.0 | 9.0 | 15.0 | 22.0 | 33.0 | 54.1 |
| 1 | 11.0 | 14.0 | 26.0 | 35.0 | 51.0 | 75.1 |
| 2 | 10.0 | 19.0 | 35.8 | 48.0 | 65.2 | 92.0 |
| 3 | 26.0 | 27.0 | 43.0 | 59.0 | 82.0 | 111.0 |
| 4 | 27.0 | 36.0 | 57.8 | 78.0 | 104.2 | 136.1 |
| 5 | 53.0 | 46.9 | 71.0 | 97.0 | 121.5 | 158.1 |
| 6 | 78.0 | 59.0 | 92.8 | 119.0 | 150.0 | 195.0 |
| 7 | 102.0 | 78.0 | 116.8 | 146.0 | 178.2 | 233.1 |
| 8 | 145.0 | 100.0 | 135.8 | 170.0 | 211.0 | 277.0 |
| 9 | 219.0 | 120.0 | 163.0 | 206.0 | 245.2 | 295.4 |
| 10 | 274.0 | 137.9 | 186.0 | 227.5 | 273.0 | 336.1 |
| 11 | 301.0 | 167.0 | 216.8 | 254.5 | 296.0 | 357.1 |
| 12 | 347.0 | 192.9 | 233.0 | 272.0 | 312.0 | 387.1 |
| 13 | 346.0 | 208.0 | 248.8 | 286.0 | 331.2 | 389.2 |
| 14 | 394.0 | 206.8 | 255.8 | 286.5 | 328.0 | 395.0 |
| 15 | 340.0 | 203.8 | 250.0 | 287.5 | 326.0 | 398.2 |
| 16 | 297.0 | 200.0 | 243.0 | 271.5 | 309.2 | 375.3 |
| 17 | 298.0 | 185.9 | 217.8 | 258.0 | 295.0 | 366.0 |
| 18 | 201.0 | 161.0 | 200.8 | 237.0 | 271.0 | 338.1 |
| 19 | 164.0 | 141.0 | 178.8 | 208.0 | 243.2 | 302.1 |
| 20 | 140.0 | 118.0 | 149.0 | 182.0 | 214.0 | 269.0 |
| 21 | 136.0 | 93.9 | 122.8 | 155.0 | 187.0 | 233.4 |
| 22 | 117.0 | 67.0 | 102.0 | 134.0 | 163.0 | 198.1 |
| 23 | 100.0 | 51.0 | 82.0 | 108.0 | 136.2 | 176.0 |
| 24 | 100.0 | 36.0 | 60.8 | 86.0 | 113.2 | 145.0 |
| 25 | 107.0 | 24.0 | 44.8 | 66.0 | 91.2 | 126.1 |
| 26 | 69.0 | 17.0 | 32.0 | 52.0 | 72.0 | 111.1 |
| 27 | 62.0 | 9.0 | 24.0 | 39.0 | 57.0 | 89.0 |
| 28 | 64.0 | 7.0 | 17.0 | 30.0 | 47.0 | 73.2 |
| 29 | 36.0 | 4.0 | 12.0 | 22.0 | 37.0 | 65.0 |
| 30 | 41.0 | 2.0 | 9.0 | 16.0 | 30.0 | 51.0 |
| 31 | 51.0 | 2.0 | 6.0 | 12.5 | 23.2 | 48.1 |
| 32 | 34.0 | 1.0 | 4.0 | 9.0 | 19.0 | 38.0 |
| 33 | 25.0 | 0.0 | 3.0 | 7.0 | 15.0 | 36.1 |
| 34 | 21.0 | 0.0 | 2.0 | 5.0 | 13.0 | 31.0 |
| 35 | 25.0 | 0.0 | 2.0 | 4.5 | 11.0 | 29.1 |
| 36 | 19.0 | 0.0 | 1.0 | 3.0 | 9.0 | 25.1 |
| 37 | 14.0 | 0.0 | 1.0 | 3.0 | 9.0 | 23.1 |
| 38 | 8.0 | 0.0 | 0.0 | 2.0 | 8.0 | 24.1 |

Table S17: **Weekly calibration – Italy.** Observed weekly hospitalisation counts and the 5th, 25th, 50th (median), 75th, and 95th percentiles of the simulated distributions across the 300 retained simulations.

| Week | Observed | Sim Q05 | Sim Q25 | Sim Median | Sim Q75 | Sim Q95 |
|---|---|---|---|---|---|---|
| 0 | 65.0 | 24.0 | 46.0 | 73.5 | 100.5 | 163.1 |
| 1 | 94.0 | 52.0 | 96.8 | 150.5 | 198.2 | 297.7 |
| 2 | 109.0 | 84.9 | 153.0 | 225.0 | 290.2 | 410.1 |
| 3 | 172.0 | 132.9 | 224.0 | 328.0 | 406.5 | 564.4 |
| 4 | 240.0 | 199.0 | 331.0 | 463.5 | 564.0 | 748.3 |
| 5 | 396.0 | 305.9 | 449.8 | 628.0 | 781.5 | 974.4 |
| 6 | 581.0 | 439.0 | 639.8 | 840.5 | 1,042.2 | 1,283.2 |
| 7 | 873.0 | 619.5 | 857.0 | 1,103.5 | 1,345.8 | 1,666.4 |
| 8 | 1,193.0 | 857.0 | 1,143.8 | 1,403.5 | 1,686.8 | 2,085.3 |
| 9 | 1,711.0 | 1,193.1 | 1,455.8 | 1,724.0 | 2,047.5 | 2,443.0 |
| 10 | 2,189.0 | 1,494.5 | 1,794.8 | 2,049.0 | 2,367.2 | 2,813.2 |
| 11 | 2,652.0 | 1,722.7 | 2,069.0 | 2,356.5 | 2,636.2 | 3,061.7 |
| 12 | 3,057.0 | 2,005.8 | 2,286.2 | 2,555.5 | 2,837.5 | 3,214.8 |
| 13 | 3,167.0 | 2,131.8 | 2,438.2 | 2,704.5 | 2,964.0 | 3,310.7 |
| 14 | 3,177.0 | 2,193.3 | 2,503.5 | 2,748.5 | 2,988.5 | 3,299.5 |
| 15 | 2,982.0 | 2,184.8 | 2,477.2 | 2,702.0 | 2,953.5 | 3,323.1 |
| 16 | 2,624.0 | 2,064.8 | 2,338.2 | 2,574.0 | 2,829.2 | 3,166.9 |
| 17 | 2,438.0 | 1,902.5 | 2,137.5 | 2,385.5 | 2,647.5 | 2,995.0 |
| 18 | 1,814.0 | 1,644.2 | 1,915.8 | 2,152.0 | 2,394.0 | 2,749.6 |
| 19 | 1,428.0 | 1,342.0 | 1,645.5 | 1,870.0 | 2,120.0 | 2,458.2 |
| 20 | 1,296.0 | 1,091.0 | 1,358.0 | 1,593.0 | 1,823.2 | 2,145.2 |
| 21 | 1,130.0 | 821.8 | 1,085.8 | 1,332.0 | 1,544.0 | 1,834.1 |
| 22 | 948.0 | 615.9 | 862.8 | 1,070.5 | 1,289.0 | 1,532.0 |
| 23 | 903.0 | 442.9 | 645.2 | 853.5 | 1,029.0 | 1,295.6 |
| 24 | 818.0 | 311.0 | 473.2 | 643.5 | 817.2 | 1,035.2 |
| 25 | 675.0 | 217.7 | 343.2 | 485.0 | 638.2 | 853.2 |
| 26 | 559.0 | 139.9 | 237.5 | 356.0 | 504.2 | 700.9 |
| 27 | 551.0 | 95.0 | 168.2 | 259.0 | 389.0 | 562.3 |
| 28 | 445.0 | 64.0 | 116.0 | 186.5 | 295.5 | 449.0 |
| 29 | 440.0 | 42.0 | 78.8 | 134.5 | 225.0 | 372.3 |
| 30 | 347.0 | 28.9 | 52.8 | 96.0 | 177.2 | 308.2 |
| 31 | 313.0 | 18.0 | 37.0 | 71.0 | 141.5 | 252.2 |
| 32 | 289.0 | 11.0 | 26.0 | 51.5 | 112.0 | 220.1 |
| 33 | 191.0 | 8.0 | 19.0 | 40.0 | 96.0 | 187.4 |
| 34 | 205.0 | 5.0 | 13.0 | 30.5 | 75.0 | 172.1 |
| 35 | 179.0 | 3.0 | 10.0 | 24.0 | 65.0 | 165.3 |
| 36 | 143.0 | 2.0 | 8.0 | 22.0 | 54.2 | 153.1 |
| 37 | 118.0 | 2.0 | 7.0 | 18.0 | 53.0 | 150.2 |
| 38 | 104.0 | 1.0 | 5.8 | 16.0 | 49.0 | 164.2 |

Table S18: **Weekly calibration – Latvia.** Observed weekly hospitalisation counts and the 5th, 25th, 50th (median), 75th, and 95th percentiles of the simulated distributions across the 300 retained simulations.

| Week | Observed | Sim Q05 | Sim Q25 | Sim Median | Sim Q75 | Sim Q95 |
|---|---|---|---|---|---|---|
| 0 | 3.0 | 0.0 | 2.0 | 5.0 | 10.0 | 14.0 |
| 1 | 3.0 | 1.0 | 4.0 | 10.0 | 16.0 | 22.1 |
| 2 | 4.0 | 1.0 | 6.0 | 13.0 | 19.0 | 28.1 |
| 3 | 10.0 | 2.0 | 9.8 | 17.5 | 24.0 | 33.1 |
| 4 | 9.0 | 4.0 | 12.0 | 22.0 | 29.0 | 39.0 |
| 5 | 18.0 | 6.0 | 17.0 | 28.0 | 37.0 | 48.1 |
| 6 | 22.0 | 8.0 | 22.0 | 34.0 | 44.0 | 59.1 |
| 7 | 30.0 | 12.0 | 28.0 | 41.0 | 50.2 | 66.0 |
| 8 | 32.0 | 15.9 | 38.0 | 49.0 | 60.0 | 77.1 |
| 9 | 55.0 | 19.0 | 44.0 | 56.5 | 72.0 | 91.0 |
| 10 | 79.0 | 27.0 | 50.8 | 63.0 | 76.2 | 104.0 |
| 11 | 89.0 | 33.0 | 56.8 | 71.0 | 91.0 | 120.0 |
| 12 | 111.0 | 34.9 | 61.0 | 77.0 | 92.2 | 124.0 |
| 13 | 119.0 | 39.0 | 65.8 | 82.0 | 101.0 | 126.0 |
| 14 | 94.0 | 40.0 | 66.0 | 82.0 | 100.0 | 127.1 |
| 15 | 101.0 | 46.0 | 66.0 | 79.5 | 98.0 | 127.0 |
| 16 | 92.0 | 40.0 | 65.0 | 78.5 | 95.2 | 121.1 |
| 17 | 86.0 | 41.0 | 62.0 | 74.0 | 90.0 | 116.0 |
| 18 | 55.0 | 38.0 | 56.0 | 68.0 | 80.0 | 103.0 |
| 19 | 51.0 | 32.0 | 49.0 | 60.0 | 73.2 | 93.0 |
| 20 | 49.0 | 28.0 | 40.0 | 51.5 | 63.0 | 80.1 |
| 21 | 32.0 | 21.0 | 34.8 | 44.0 | 55.0 | 69.0 |
| 22 | 30.0 | 16.0 | 28.0 | 37.0 | 45.0 | 63.0 |
| 23 | 39.0 | 12.0 | 21.0 | 29.0 | 38.0 | 51.0 |
| 24 | 37.0 | 8.0 | 16.0 | 23.0 | 32.0 | 46.1 |
| 25 | 29.0 | 6.0 | 12.0 | 18.0 | 26.0 | 38.0 |
| 26 | 27.0 | 4.0 | 8.0 | 13.0 | 20.0 | 33.0 |
| 27 | 25.0 | 2.0 | 6.0 | 10.0 | 16.2 | 26.0 |
| 28 | 12.0 | 1.0 | 4.0 | 7.0 | 13.0 | 21.1 |
| 29 | 7.0 | 1.0 | 3.0 | 6.0 | 11.0 | 22.0 |
| 30 | 11.0 | 0.0 | 2.0 | 4.0 | 8.0 | 17.0 |
| 31 | 7.0 | 0.0 | 1.0 | 3.0 | 6.0 | 14.0 |
| 32 | 10.0 | 0.0 | 1.0 | 2.0 | 5.0 | 11.0 |
| 33 | 6.0 | 0.0 | 0.0 | 1.0 | 4.0 | 10.0 |
| 34 | 6.0 | 0.0 | 0.0 | 1.0 | 3.0 | 10.1 |
| 35 | 7.0 | 0.0 | 0.0 | 1.0 | 3.0 | 7.0 |
| 36 | 4.0 | 0.0 | 0.0 | 1.0 | 2.0 | 7.0 |
| 37 | 1.0 | 0.0 | 0.0 | 0.0 | 2.0 | 7.0 |
| 38 | 3.0 | 0.0 | 0.0 | 0.0 | 2.0 | 6.0 |

Table S19: **Weekly calibration – Lithuania.** Observed weekly hospitalisation counts and the 5th, 25th, 50th (median), 75th, and 95th percentiles of the simulated distributions across the 300 retained simulations.

| Week | Observed | Sim Q05 | Sim Q25 | Sim Median | Sim Q75 | Sim Q95 |
|---|---|---|---|---|---|---|
| 0 | 6.0 | 3.0 | 4.0 | 7.0 | 11.0 | 15.0 |
| 1 | 4.0 | 5.0 | 10.0 | 14.0 | 18.0 | 28.0 |
| 2 | 2.0 | 9.0 | 14.0 | 18.0 | 24.0 | 35.0 |
| 3 | 5.0 | 11.9 | 18.0 | 24.0 | 30.0 | 42.0 |
| 4 | 17.0 | 14.0 | 23.0 | 31.0 | 39.0 | 53.0 |
| 5 | 26.0 | 20.0 | 31.0 | 38.5 | 48.0 | 64.1 |
| 6 | 24.0 | 25.0 | 38.0 | 49.0 | 60.2 | 77.0 |
| 7 | 43.0 | 31.0 | 48.0 | 60.0 | 75.0 | 95.2 |
| 8 | 69.0 | 40.0 | 58.8 | 71.0 | 88.0 | 114.0 |
| 9 | 85.0 | 49.0 | 67.0 | 84.0 | 104.0 | 131.0 |
| 10 | 141.0 | 58.0 | 78.0 | 95.0 | 111.2 | 148.1 |
| 11 | 133.0 | 63.0 | 85.8 | 105.0 | 129.2 | 161.2 |
| 12 | 148.0 | 71.0 | 93.0 | 110.0 | 132.0 | 167.1 |
| 13 | 136.0 | 72.0 | 99.0 | 119.5 | 144.0 | 175.1 |
| 14 | 150.0 | 77.0 | 101.0 | 120.0 | 140.2 | 177.0 |
| 15 | 132.0 | 79.0 | 101.0 | 123.0 | 139.0 | 171.1 |
| 16 | 121.0 | 75.0 | 99.0 | 118.5 | 136.0 | 164.0 |
| 17 | 128.0 | 75.0 | 94.0 | 111.0 | 129.0 | 149.1 |
| 18 | 91.0 | 66.0 | 85.0 | 101.0 | 117.0 | 137.0 |
| 19 | 68.0 | 57.0 | 76.0 | 88.0 | 104.0 | 128.1 |
| 20 | 77.0 | 46.0 | 62.0 | 78.5 | 92.0 | 114.0 |
| 21 | 46.0 | 38.0 | 53.8 | 66.0 | 79.0 | 95.0 |
| 22 | 56.0 | 30.0 | 42.0 | 54.0 | 67.0 | 84.1 |
| 23 | 37.0 | 21.0 | 32.8 | 44.0 | 55.0 | 72.1 |
| 24 | 39.0 | 14.0 | 24.0 | 35.0 | 45.0 | 60.0 |
| 25 | 35.0 | 10.0 | 18.0 | 27.0 | 37.0 | 55.0 |
| 26 | 24.0 | 7.0 | 14.0 | 20.0 | 29.0 | 44.1 |
| 27 | 32.0 | 4.0 | 9.0 | 15.0 | 24.0 | 37.0 |
| 28 | 23.0 | 2.0 | 6.0 | 11.0 | 19.0 | 31.0 |
| 29 | 24.0 | 1.0 | 4.0 | 8.0 | 14.0 | 25.1 |
| 30 | 16.0 | 1.0 | 3.0 | 6.0 | 11.0 | 21.0 |
| 31 | 9.0 | 0.0 | 2.0 | 4.0 | 9.2 | 19.0 |
| 32 | 7.0 | 0.0 | 1.0 | 3.0 | 8.0 | 16.0 |
| 33 | 11.0 | 0.0 | 1.0 | 2.0 | 5.0 | 14.0 |
| 34 | 8.0 | 0.0 | 0.0 | 2.0 | 4.0 | 11.1 |
| 35 | 6.0 | 0.0 | 0.0 | 1.0 | 4.0 | 11.0 |
| 36 | 12.0 | 0.0 | 0.0 | 1.0 | 3.0 | 10.0 |
| 37 | 8.0 | 0.0 | 0.0 | 1.0 | 2.2 | 8.0 |
| 38 | 2.0 | 0.0 | 0.0 | 0.0 | 2.0 | 8.1 |

Table S20: **Weekly calibration – Luxembourg.** Observed weekly hospitalisation counts and the 5th, 25th, 50th (median), 75th, and 95th percentiles of the simulated distributions across the 300 retained simulations.

| Week | Observed | Sim Q05 | Sim Q25 | Sim Median | Sim Q75 | Sim Q95 |
|---|---|---|---|---|---|---|
| 0 | 2.0 | 1.0 | 2.0 | 4.0 | 6.0 | 11.0 |
| 1 | 1.0 | 2.0 | 4.0 | 6.0 | 9.0 | 15.0 |
| 2 | 0.0 | 2.0 | 5.0 | 8.0 | 11.0 | 17.0 |
| 3 | 3.0 | 3.0 | 7.0 | 10.0 | 12.0 | 18.0 |
| 4 | 6.0 | 4.0 | 7.0 | 11.0 | 15.0 | 21.0 |
| 5 | 5.0 | 4.0 | 8.8 | 13.0 | 16.0 | 22.1 |
| 6 | 7.0 | 6.0 | 11.0 | 15.0 | 19.0 | 25.0 |
| 7 | 9.0 | 6.0 | 12.0 | 16.0 | 21.0 | 30.0 |
| 8 | 21.0 | 8.0 | 13.0 | 18.0 | 22.2 | 34.0 |
| 9 | 18.0 | 9.0 | 15.0 | 20.0 | 26.0 | 33.1 |
| 10 | 18.0 | 10.0 | 16.0 | 21.0 | 27.0 | 36.1 |
| 11 | 26.0 | 9.9 | 16.0 | 22.0 | 28.0 | 38.0 |
| 12 | 31.0 | 11.0 | 18.0 | 24.0 | 30.0 | 38.1 |
| 13 | 37.0 | 11.0 | 17.0 | 23.0 | 31.0 | 38.1 |
| 14 | 38.0 | 13.0 | 19.0 | 24.0 | 29.0 | 38.0 |
| 15 | 46.0 | 12.0 | 19.0 | 23.0 | 29.0 | 36.1 |
| 16 | 27.0 | 10.0 | 17.0 | 22.0 | 27.0 | 35.1 |
| 17 | 15.0 | 11.0 | 16.0 | 20.0 | 25.0 | 33.1 |
| 18 | 13.0 | 8.0 | 14.8 | 19.0 | 24.0 | 31.0 |
| 19 | 8.0 | 7.0 | 13.0 | 17.0 | 21.0 | 29.1 |
| 20 | 14.0 | 6.0 | 10.0 | 14.0 | 19.0 | 25.1 |
| 21 | 12.0 | 3.0 | 9.0 | 13.0 | 16.2 | 23.0 |
| 22 | 10.0 | 3.0 | 7.0 | 11.0 | 14.0 | 22.0 |
| 23 | 10.0 | 2.0 | 5.0 | 8.0 | 12.0 | 18.0 |
| 24 | 5.0 | 1.0 | 4.0 | 7.0 | 10.2 | 17.0 |
| 25 | 11.0 | 0.0 | 3.0 | 6.0 | 9.0 | 14.0 |
| 26 | 10.0 | 0.0 | 2.0 | 5.0 | 7.0 | 12.0 |
| 27 | 4.0 | 0.0 | 2.0 | 4.0 | 6.0 | 11.0 |
| 28 | 5.0 | 0.0 | 1.0 | 3.0 | 5.0 | 9.1 |
| 29 | 5.0 | 0.0 | 1.0 | 2.0 | 4.0 | 8.0 |
| 30 | 3.0 | 0.0 | 0.0 | 1.0 | 3.0 | 6.0 |
| 31 | 2.0 | 0.0 | 0.0 | 1.0 | 3.0 | 6.0 |
| 32 | 5.0 | 0.0 | 0.0 | 1.0 | 2.0 | 5.0 |
| 33 | 1.0 | 0.0 | 0.0 | 0.0 | 2.0 | 4.1 |
| 34 | 2.0 | 0.0 | 0.0 | 0.0 | 1.0 | 5.0 |
| 35 | 2.0 | 0.0 | 0.0 | 0.0 | 1.0 | 4.0 |
| 36 | 1.0 | 0.0 | 0.0 | 0.0 | 1.0 | 3.1 |
| 37 | 1.0 | 0.0 | 0.0 | 0.0 | 1.0 | 4.0 |
| 38 | 1.0 | 0.0 | 0.0 | 0.0 | 1.0 | 3.0 |

Table S21: **Weekly calibration – Malta.** Observed weekly hospitalisation counts and the 5th, 25th, 50th (median), 75th, and 95th percentiles of the simulated distributions across the 300 retained simulations.

| Week | Observed | Sim Q05 | Sim Q25 | Sim Median | Sim Q75 | Sim Q95 |
|---|---|---|---|---|---|---|
| 0 | 1.0 | 1.0 | 2.0 | 4.0 | 6.0 | 10.0 |
| 1 | 1.0 | 1.0 | 3.0 | 5.0 | 8.0 | 12.0 |
| 2 | 4.0 | 2.0 | 4.0 | 6.0 | 8.2 | 13.1 |
| 3 | 2.0 | 2.0 | 5.0 | 7.0 | 10.0 | 15.0 |
| 4 | 0.0 | 3.0 | 6.0 | 8.0 | 11.0 | 17.0 |
| 5 | 5.0 | 4.0 | 7.0 | 10.0 | 13.0 | 21.0 |
| 6 | 4.0 | 5.0 | 8.0 | 12.0 | 15.0 | 24.0 |
| 7 | 7.0 | 6.0 | 10.0 | 13.0 | 17.0 | 26.0 |
| 8 | 10.0 | 7.0 | 11.0 | 15.0 | 21.0 | 29.0 |
| 9 | 21.0 | 8.0 | 13.0 | 17.0 | 22.0 | 29.1 |
| 10 | 27.0 | 8.0 | 14.0 | 18.0 | 25.0 | 34.1 |
| 11 | 21.0 | 8.0 | 15.0 | 20.0 | 26.0 | 36.0 |
| 12 | 32.0 | 10.0 | 15.0 | 20.0 | 26.0 | 36.1 |
| 13 | 24.0 | 10.0 | 17.0 | 22.0 | 28.0 | 39.0 |
| 14 | 38.0 | 10.0 | 16.0 | 21.0 | 27.0 | 37.1 |
| 15 | 28.0 | 10.0 | 17.0 | 22.0 | 27.0 | 36.0 |
| 16 | 23.0 | 9.0 | 15.0 | 20.0 | 26.0 | 36.0 |
| 17 | 19.0 | 9.0 | 14.0 | 18.5 | 23.2 | 32.0 |
| 18 | 17.0 | 7.0 | 12.0 | 17.0 | 21.2 | 30.0 |
| 19 | 11.0 | 7.0 | 11.0 | 14.5 | 20.0 | 27.0 |
| 20 | 10.0 | 5.0 | 9.0 | 13.0 | 17.0 | 26.0 |
| 21 | 9.0 | 4.0 | 8.0 | 11.5 | 15.0 | 22.0 |
| 22 | 5.0 | 3.0 | 6.0 | 9.0 | 13.0 | 19.0 |
| 23 | 9.0 | 2.0 | 4.0 | 7.0 | 10.0 | 16.0 |
| 24 | 8.0 | 1.0 | 4.0 | 6.0 | 9.0 | 14.0 |
| 25 | 8.0 | 1.0 | 3.0 | 4.0 | 7.0 | 12.0 |
| 26 | 4.0 | 0.0 | 2.0 | 4.0 | 6.0 | 10.0 |
| 27 | 4.0 | 0.0 | 1.0 | 3.0 | 5.0 | 8.0 |
| 28 | 2.0 | 0.0 | 1.0 | 2.0 | 4.0 | 7.0 |
| 29 | 2.0 | 0.0 | 0.0 | 1.0 | 3.0 | 6.0 |
| 30 | 2.0 | 0.0 | 0.0 | 1.0 | 3.0 | 6.0 |
| 31 | 2.0 | 0.0 | 0.0 | 1.0 | 2.0 | 5.0 |
| 32 | 5.0 | 0.0 | 0.0 | 1.0 | 2.0 | 4.0 |
| 33 | 0.0 | 0.0 | 0.0 | 0.0 | 1.0 | 4.0 |
| 34 | 0.0 | 0.0 | 0.0 | 0.0 | 1.0 | 3.0 |
| 35 | 2.0 | 0.0 | 0.0 | 0.0 | 1.0 | 2.1 |
| 36 | 5.0 | 0.0 | 0.0 | 0.0 | 1.0 | 2.0 |
| 37 | 3.0 | 0.0 | 0.0 | 0.0 | 0.0 | 2.0 |
| 38 | 1.0 | 0.0 | 0.0 | 0.0 | 0.0 | 2.0 |

Table S22: **Weekly calibration – Netherlands.** Observed weekly hospitalisation counts and the 5th, 25th, 50th (median), 75th, and 95th percentiles of the simulated distributions across the 300 retained simulations.

| Week | Observed | Sim Q05 | Sim Q25 | Sim Median | Sim Q75 | Sim Q95 |
|---|---|---|---|---|---|---|
| 0 | 27.0 | 9.0 | 19.0 | 28.0 | 40.0 | 62.0 |
| 1 | 16.0 | 15.0 | 35.0 | 48.0 | 70.2 | 103.0 |
| 2 | 28.0 | 24.9 | 50.0 | 69.0 | 96.2 | 148.1 |
| 3 | 29.0 | 38.0 | 73.0 | 91.5 | 122.0 | 166.2 |
| 4 | 57.0 | 56.9 | 100.0 | 124.0 | 159.0 | 205.1 |
| 5 | 109.0 | 79.9 | 138.0 | 167.5 | 205.0 | 263.2 |
| 6 | 148.0 | 109.0 | 179.0 | 220.5 | 262.0 | 322.2 |
| 7 | 195.0 | 155.9 | 228.0 | 282.0 | 325.0 | 399.1 |
| 8 | 284.0 | 206.9 | 286.0 | 342.0 | 398.2 | 473.0 |
| 9 | 386.0 | 258.7 | 336.0 | 400.0 | 469.2 | 557.1 |
| 10 | 466.0 | 324.5 | 403.8 | 460.0 | 536.2 | 653.2 |
| 11 | 557.0 | 371.8 | 449.0 | 516.0 | 587.8 | 712.7 |
| 12 | 742.0 | 420.9 | 493.0 | 560.5 | 632.0 | 749.1 |
| 13 | 707.0 | 445.9 | 521.0 | 585.0 | 658.5 | 768.6 |
| 14 | 724.0 | 453.9 | 524.0 | 585.0 | 670.2 | 785.0 |
| 15 | 639.0 | 459.9 | 526.8 | 583.0 | 643.0 | 749.0 |
| 16 | 623.0 | 432.6 | 498.0 | 552.5 | 612.2 | 709.0 |
| 17 | 495.0 | 399.0 | 459.5 | 508.5 | 565.0 | 655.2 |
| 18 | 395.0 | 354.0 | 409.8 | 463.5 | 514.0 | 590.2 |
| 19 | 338.0 | 295.9 | 352.0 | 398.5 | 458.2 | 533.0 |
| 20 | 317.0 | 239.8 | 298.8 | 339.5 | 403.2 | 464.0 |
| 21 | 254.0 | 182.0 | 242.8 | 288.0 | 341.0 | 405.1 |
| 22 | 223.0 | 133.9 | 190.8 | 232.5 | 287.0 | 350.1 |
| 23 | 188.0 | 103.7 | 148.0 | 190.0 | 237.0 | 297.1 |
| 24 | 190.0 | 70.0 | 111.8 | 151.0 | 195.0 | 251.2 |
| 25 | 153.0 | 49.0 | 83.8 | 115.0 | 152.2 | 216.0 |
| 26 | 136.0 | 29.0 | 60.0 | 87.5 | 125.0 | 174.1 |
| 27 | 126.0 | 22.0 | 41.0 | 65.0 | 101.2 | 141.0 |
| 28 | 92.0 | 14.9 | 31.0 | 49.0 | 82.2 | 124.0 |
| 29 | 91.0 | 7.0 | 21.0 | 38.0 | 62.0 | 105.3 |
| 30 | 78.0 | 5.0 | 14.8 | 28.0 | 50.2 | 90.0 |
| 31 | 79.0 | 4.0 | 11.0 | 20.0 | 41.2 | 75.1 |
| 32 | 54.0 | 2.0 | 7.0 | 16.0 | 34.0 | 66.1 |
| 33 | 52.0 | 1.0 | 5.0 | 13.0 | 27.2 | 56.0 |
| 34 | 53.0 | 0.0 | 3.0 | 10.0 | 24.0 | 52.1 |
| 35 | 41.0 | 0.0 | 3.0 | 8.0 | 20.0 | 44.2 |
| 36 | 34.0 | 0.0 | 2.0 | 7.5 | 18.0 | 41.2 |
| 37 | 31.0 | 0.0 | 2.0 | 6.0 | 16.0 | 40.2 |
| 38 | 21.0 | 0.0 | 1.0 | 5.0 | 14.2 | 38.2 |

Table S23: **Weekly calibration – Norway.** Observed weekly hospitalisation counts and the 5th, 25th, 50th (median), 75th, and 95th percentiles of the simulated distributions across the 300 retained simulations.

| Week | Observed | Sim Q05 | Sim Q25 | Sim Median | Sim Q75 | Sim Q95 |
|---|---|---|---|---|---|---|
| 0 | 3.0 | 4.0 | 10.0 | 15.0 | 22.0 | 32.0 |
| 1 | 13.0 | 9.0 | 16.0 | 25.0 | 35.0 | 50.0 |
| 2 | 15.0 | 13.0 | 23.0 | 34.0 | 47.0 | 68.0 |
| 3 | 22.0 | 20.0 | 30.8 | 43.0 | 61.0 | 81.0 |
| 4 | 21.0 | 28.0 | 41.0 | 59.0 | 79.0 | 104.0 |
| 5 | 42.0 | 42.0 | 55.0 | 78.0 | 100.2 | 130.1 |
| 6 | 60.0 | 55.0 | 77.0 | 97.0 | 122.0 | 155.1 |
| 7 | 100.0 | 71.9 | 99.0 | 122.0 | 154.5 | 194.1 |
| 8 | 148.0 | 91.9 | 125.0 | 153.5 | 182.0 | 231.2 |
| 9 | 185.0 | 113.0 | 155.5 | 181.0 | 217.2 | 270.2 |
| 10 | 245.0 | 131.9 | 178.8 | 209.5 | 243.0 | 300.1 |
| 11 | 277.0 | 158.0 | 204.0 | 235.0 | 270.2 | 322.0 |
| 12 | 322.0 | 172.9 | 222.8 | 254.0 | 290.0 | 345.0 |
| 13 | 350.0 | 194.9 | 236.8 | 262.0 | 300.0 | 360.2 |
| 14 | 340.0 | 192.9 | 236.8 | 271.0 | 305.0 | 361.1 |
| 15 | 337.0 | 199.9 | 234.8 | 265.0 | 301.0 | 358.2 |
| 16 | 290.0 | 183.0 | 227.0 | 253.5 | 291.2 | 356.3 |
| 17 | 230.0 | 172.0 | 208.8 | 237.5 | 266.2 | 328.1 |
| 18 | 189.0 | 151.0 | 186.8 | 220.0 | 249.2 | 301.3 |
| 19 | 141.0 | 131.8 | 163.8 | 196.5 | 220.0 | 279.1 |
| 20 | 137.0 | 109.0 | 142.0 | 167.0 | 193.0 | 233.1 |
| 21 | 122.0 | 88.0 | 119.0 | 139.5 | 165.0 | 203.0 |
| 22 | 107.0 | 68.0 | 92.8 | 117.5 | 137.0 | 179.0 |
| 23 | 83.0 | 51.0 | 75.0 | 93.0 | 114.2 | 147.1 |
| 24 | 92.0 | 39.0 | 56.8 | 73.0 | 93.2 | 123.0 |
| 25 | 86.0 | 29.0 | 41.0 | 56.5 | 76.0 | 104.0 |
| 26 | 76.0 | 18.0 | 31.0 | 43.5 | 59.0 | 85.0 |
| 27 | 44.0 | 13.0 | 22.0 | 31.0 | 45.0 | 72.0 |
| 28 | 39.0 | 9.0 | 15.0 | 24.0 | 35.2 | 60.0 |
| 29 | 36.0 | 6.0 | 11.0 | 17.0 | 27.0 | 49.2 |
| 30 | 38.0 | 3.0 | 7.0 | 12.5 | 22.0 | 39.1 |
| 31 | 22.0 | 2.0 | 4.0 | 9.0 | 18.0 | 37.1 |
| 32 | 32.0 | 1.0 | 3.0 | 7.0 | 14.0 | 29.1 |
| 33 | 24.0 | 0.0 | 2.0 | 5.0 | 11.0 | 27.0 |
| 34 | 22.0 | 0.0 | 1.8 | 4.0 | 8.2 | 21.1 |
| 35 | 14.0 | 0.0 | 1.0 | 3.0 | 7.2 | 20.2 |
| 36 | 15.0 | 0.0 | 1.0 | 2.0 | 6.0 | 18.1 |
| 37 | 8.0 | 0.0 | 0.0 | 2.0 | 6.0 | 19.1 |
| 38 | 10.0 | 0.0 | 0.0 | 1.0 | 6.0 | 17.1 |

Table S24: **Weekly calibration – Poland.** Observed weekly hospitalisation counts and the 5th, 25th, 50th (median), 75th, and 95th percentiles of the simulated distributions across the 300 retained simulations.

| Week | Observed | Sim Q05 | Sim Q25 | Sim Median | Sim Q75 | Sim Q95 |
|---|---|---|---|---|---|---|
| 0 | 51.0 | 92.9 | 176.0 | 252.0 | 348.0 | 483.1 |
| 1 | 55.0 | 79.9 | 149.8 | 231.5 | 325.0 | 470.0 |
| 2 | 52.0 | 73.8 | 149.0 | 242.5 | 344.0 | 504.1 |
| 3 | 108.0 | 89.0 | 184.5 | 287.0 | 395.8 | 568.5 |
| 4 | 157.0 | 132.9 | 234.5 | 363.0 | 499.5 | 681.0 |
| 5 | 232.0 | 183.7 | 298.8 | 464.5 | 611.5 | 836.0 |
| 6 | 392.0 | 255.8 | 393.8 | 576.5 | 731.5 | 979.0 |
| 7 | 545.0 | 355.9 | 507.5 | 673.0 | 885.2 | 1,156.2 |
| 8 | 763.0 | 455.1 | 638.8 | 821.5 | 1,041.2 | 1,298.0 |
| 9 | 1,068.0 | 581.0 | 775.8 | 968.0 | 1,186.0 | 1,454.2 |
| 10 | 1,353.0 | 687.8 | 900.8 | 1,090.0 | 1,322.5 | 1,611.2 |
| 11 | 1,604.0 | 826.6 | 1,033.5 | 1,217.5 | 1,448.0 | 1,720.5 |
| 12 | 1,969.0 | 986.5 | 1,135.8 | 1,312.0 | 1,522.8 | 1,825.0 |
| 13 | 1,951.0 | 1,051.8 | 1,224.8 | 1,388.0 | 1,580.5 | 1,955.5 |
| 14 | 2,019.0 | 1,101.8 | 1,281.5 | 1,439.0 | 1,618.8 | 1,971.0 |
| 15 | 1,785.0 | 1,098.8 | 1,273.0 | 1,426.5 | 1,626.8 | 1,966.3 |
| 16 | 1,653.0 | 1,067.0 | 1,234.0 | 1,388.5 | 1,605.0 | 1,910.2 |
| 17 | 1,427.0 | 996.9 | 1,182.5 | 1,343.0 | 1,547.0 | 1,881.2 |
| 18 | 1,100.0 | 901.0 | 1,056.8 | 1,245.5 | 1,465.0 | 1,739.5 |
| 19 | 910.0 | 777.0 | 964.2 | 1,145.5 | 1,349.0 | 1,654.3 |
| 20 | 781.0 | 664.5 | 843.8 | 1,025.0 | 1,222.8 | 1,529.0 |
| 21 | 671.0 | 546.9 | 723.8 | 901.5 | 1,073.0 | 1,364.4 |
| 22 | 607.0 | 427.9 | 602.0 | 762.0 | 931.5 | 1,195.3 |
| 23 | 545.0 | 341.9 | 498.8 | 637.0 | 800.2 | 1,012.0 |
| 24 | 492.0 | 263.9 | 385.0 | 528.5 | 689.2 | 866.5 |
| 25 | 411.0 | 186.9 | 303.5 | 416.0 | 576.2 | 732.4 |
| 26 | 383.0 | 141.7 | 242.0 | 332.5 | 457.5 | 630.0 |
| 27 | 310.0 | 101.0 | 177.5 | 260.0 | 384.2 | 529.0 |
| 28 | 275.0 | 73.0 | 133.8 | 209.0 | 313.2 | 455.1 |
| 29 | 257.0 | 58.0 | 102.0 | 162.5 | 257.0 | 392.1 |
| 30 | 217.0 | 39.0 | 78.0 | 123.0 | 210.0 | 337.2 |
| 31 | 167.0 | 29.0 | 58.0 | 100.0 | 167.8 | 299.0 |
| 32 | 150.0 | 20.0 | 44.8 | 81.0 | 144.2 | 249.5 |
| 33 | 127.0 | 17.0 | 38.2 | 63.0 | 122.2 | 230.1 |
| 34 | 113.0 | 11.9 | 29.8 | 51.5 | 96.2 | 194.2 |
| 35 | 102.0 | 10.0 | 25.0 | 43.5 | 84.2 | 180.2 |
| 36 | 101.0 | 6.0 | 19.0 | 37.0 | 74.2 | 158.3 |
| 37 | 76.0 | 5.0 | 17.0 | 35.0 | 66.0 | 156.2 |
| 38 | 52.0 | 5.0 | 15.0 | 30.0 | 60.2 | 141.0 |

Table S25: **Weekly calibration – Portugal.** Observed weekly hospitalisation counts and the 5th, 25th, 50th (median), 75th, and 95th percentiles of the simulated distributions across the 300 retained simulations.

| Week | Observed | Sim Q05 | Sim Q25 | Sim Median | Sim Q75 | Sim Q95 |
|---|---|---|---|---|---|---|
| 0 | 8.0 | 4.0 | 14.0 | 25.5 | 37.0 | 56.0 |
| 1 | 22.0 | 9.9 | 25.0 | 47.0 | 64.0 | 94.0 |
| 2 | 18.0 | 14.0 | 36.0 | 68.0 | 89.0 | 124.0 |
| 3 | 28.0 | 21.0 | 50.8 | 88.0 | 114.2 | 159.1 |
| 4 | 34.0 | 33.0 | 67.0 | 116.5 | 145.0 | 193.1 |
| 5 | 77.0 | 49.0 | 96.8 | 150.0 | 188.0 | 249.0 |
| 6 | 108.0 | 66.0 | 122.0 | 191.0 | 230.2 | 292.0 |
| 7 | 182.0 | 102.9 | 167.0 | 229.0 | 278.2 | 353.0 |
| 8 | 239.0 | 132.9 | 222.8 | 274.5 | 335.2 | 410.1 |
| 9 | 331.0 | 179.8 | 269.0 | 324.0 | 379.0 | 462.0 |
| 10 | 398.0 | 227.9 | 311.0 | 372.0 | 437.2 | 514.2 |
| 11 | 492.0 | 279.4 | 352.8 | 415.5 | 473.2 | 548.1 |
| 12 | 585.0 | 313.9 | 389.5 | 453.5 | 509.8 | 584.1 |
| 13 | 536.0 | 349.9 | 409.8 | 474.5 | 526.2 | 604.2 |
| 14 | 606.0 | 355.9 | 417.8 | 476.0 | 529.2 | 626.2 |
| 15 | 529.0 | 364.9 | 417.8 | 467.0 | 523.2 | 608.2 |
| 16 | 480.0 | 342.9 | 396.0 | 440.0 | 497.8 | 598.3 |
| 17 | 437.0 | 314.0 | 362.0 | 414.5 | 473.5 | 574.0 |
| 18 | 335.0 | 277.0 | 326.8 | 380.0 | 440.0 | 523.0 |
| 19 | 271.0 | 228.9 | 280.0 | 338.0 | 393.5 | 482.4 |
| 20 | 232.0 | 184.0 | 239.0 | 285.5 | 345.0 | 427.3 |
| 21 | 194.0 | 140.9 | 199.0 | 247.0 | 293.5 | 364.1 |
| 22 | 176.0 | 110.0 | 158.5 | 198.5 | 247.0 | 309.1 |
| 23 | 155.0 | 77.0 | 123.0 | 163.0 | 204.2 | 257.1 |
| 24 | 163.0 | 53.9 | 95.8 | 127.5 | 166.0 | 227.1 |
| 25 | 157.0 | 39.0 | 70.0 | 97.0 | 133.0 | 187.1 |
| 26 | 120.0 | 27.0 | 50.0 | 75.0 | 106.0 | 151.3 |
| 27 | 94.0 | 17.0 | 37.8 | 57.0 | 79.2 | 130.0 |
| 28 | 69.0 | 11.9 | 25.0 | 43.0 | 62.2 | 110.1 |
| 29 | 74.0 | 7.0 | 19.0 | 30.0 | 51.0 | 97.0 |
| 30 | 58.0 | 5.0 | 12.8 | 22.0 | 39.2 | 77.0 |
| 31 | 61.0 | 2.0 | 8.0 | 18.0 | 32.0 | 67.1 |
| 32 | 48.0 | 2.0 | 6.0 | 13.0 | 27.0 | 56.4 |
| 33 | 34.0 | 1.0 | 4.0 | 9.0 | 20.0 | 47.0 |
| 34 | 36.0 | 0.0 | 3.0 | 8.0 | 16.0 | 39.1 |
| 35 | 35.0 | 0.0 | 2.0 | 6.0 | 14.2 | 38.1 |
| 36 | 23.0 | 0.0 | 1.0 | 4.0 | 11.2 | 32.1 |
| 37 | 21.0 | 0.0 | 1.0 | 3.0 | 10.0 | 29.0 |
| 38 | 14.0 | 0.0 | 1.0 | 3.0 | 9.0 | 31.1 |

Table S26: **Weekly calibration – Romania.** Observed weekly hospitalisation counts and the 5th, 25th, 50th (median), 75th, and 95th percentiles of the simulated distributions across the 300 retained simulations.

| Week | Observed | Sim Q05 | Sim Q25 | Sim Median | Sim Q75 | Sim Q95 |
|---|---|---|---|---|---|---|
| 0 | 33.0 | 5.0 | 17.0 | 39.0 | 56.0 | 85.1 |
| 1 | 36.0 | 12.9 | 33.8 | 65.0 | 101.0 | 150.1 |
| 2 | 33.0 | 22.0 | 52.8 | 93.5 | 134.8 | 203.0 |
| 3 | 75.0 | 36.0 | 71.8 | 129.0 | 182.2 | 246.5 |
| 4 | 94.0 | 59.0 | 105.0 | 174.0 | 239.0 | 319.3 |
| 5 | 149.0 | 88.0 | 146.2 | 225.0 | 306.2 | 398.1 |
| 6 | 187.0 | 140.9 | 209.0 | 295.0 | 390.2 | 491.1 |
| 7 | 343.0 | 213.8 | 287.5 | 381.5 | 484.0 | 596.5 |
| 8 | 439.0 | 299.4 | 390.8 | 473.5 | 587.0 | 740.1 |
| 9 | 611.0 | 389.9 | 498.0 | 597.0 | 703.8 | 853.3 |
| 10 | 822.0 | 485.9 | 620.8 | 701.5 | 808.0 | 960.9 |
| 11 | 931.0 | 588.5 | 724.2 | 817.5 | 913.2 | 1,084.0 |
| 12 | 1,121.0 | 666.9 | 792.8 | 905.5 | 998.2 | 1,168.2 |
| 13 | 1,131.0 | 727.0 | 849.0 | 958.0 | 1,059.8 | 1,233.8 |
| 14 | 1,169.0 | 748.7 | 862.0 | 980.5 | 1,083.0 | 1,254.5 |
| 15 | 1,048.0 | 744.0 | 862.8 | 970.0 | 1,066.5 | 1,252.0 |
| 16 | 940.0 | 713.0 | 820.5 | 923.0 | 1,019.2 | 1,206.2 |
| 17 | 846.0 | 656.8 | 768.0 | 850.0 | 947.2 | 1,119.3 |
| 18 | 648.0 | 572.8 | 682.0 | 773.5 | 859.0 | 1,018.3 |
| 19 | 543.0 | 486.9 | 598.8 | 678.0 | 766.2 | 896.3 |
| 20 | 476.0 | 392.9 | 507.8 | 585.5 | 657.2 | 766.7 |
| 21 | 416.0 | 320.0 | 407.0 | 481.5 | 555.2 | 665.1 |
| 22 | 333.0 | 239.0 | 321.8 | 393.5 | 464.2 | 550.2 |
| 23 | 323.0 | 172.0 | 247.0 | 305.0 | 387.5 | 467.1 |
| 24 | 299.0 | 124.9 | 185.5 | 232.0 | 303.8 | 403.1 |
| 25 | 223.0 | 87.0 | 130.5 | 181.0 | 254.0 | 314.3 |
| 26 | 222.0 | 58.0 | 90.8 | 132.5 | 198.2 | 290.1 |
| 27 | 185.0 | 39.0 | 65.8 | 96.0 | 159.2 | 224.3 |
| 28 | 175.0 | 24.0 | 46.8 | 70.0 | 125.0 | 191.0 |
| 29 | 158.0 | 14.9 | 31.0 | 52.0 | 98.0 | 156.1 |
| 30 | 113.0 | 10.0 | 22.0 | 38.0 | 78.2 | 139.2 |
| 31 | 91.0 | 7.0 | 14.0 | 27.5 | 63.0 | 110.2 |
| 32 | 101.0 | 4.0 | 10.0 | 19.0 | 50.2 | 101.1 |
| 33 | 73.0 | 2.0 | 7.0 | 15.0 | 41.0 | 87.1 |
| 34 | 71.0 | 2.0 | 6.0 | 11.0 | 33.0 | 79.1 |
| 35 | 58.0 | 1.0 | 4.0 | 9.0 | 30.0 | 74.1 |
| 36 | 58.0 | 0.0 | 3.0 | 8.0 | 27.0 | 64.1 |
| 37 | 49.0 | 0.0 | 2.8 | 6.0 | 22.0 | 63.4 |
| 38 | 39.0 | 0.0 | 2.0 | 5.5 | 23.0 | 64.2 |

Table S27: **Weekly calibration – Slovakia.** Observed weekly hospitalisation counts and the 5th, 25th, 50th (median), 75th, and 95th percentiles of the simulated distributions across the 300 retained simulations.

| **Week** | **Observed** | **Sim Q05** | **Sim Q25** | **Sim Median** | **Sim Q75** | **Sim Q95** |
|---|---|---|---|---|---|---|
| 0 | 5.0 | 4.0 | 9.0 | 15.0 | 21.0 | 36.0 |
| 1 | 8.0 | 11.9 | 17.8 | 26.0 | 36.2 | 58.1 |
| 2 | 14.0 | 15.9 | 24.0 | 34.0 | 48.2 | 76.1 |
| 3 | 19.0 | 21.0 | 34.0 | 46.0 | 59.0 | 89.1 |
| 4 | 29.0 | 29.0 | 46.0 | 59.0 | 72.2 | 105.1 |
| 5 | 53.0 | 41.0 | 60.8 | 75.0 | 91.2 | 127.0 |
| 6 | 59.0 | 49.9 | 78.8 | 96.0 | 115.0 | 152.1 |
| 7 | 100.0 | 69.0 | 98.8 | 120.5 | 145.2 | 183.1 |
| 8 | 104.0 | 85.8 | 121.0 | 147.0 | 174.2 | 212.0 |
| 9 | 174.0 | 102.0 | 139.0 | 174.5 | 203.0 | 239.2 |
| 10 | 225.0 | 128.9 | 167.8 | 203.0 | 235.0 | 277.1 |
| 11 | 263.0 | 141.9 | 186.0 | 224.5 | 254.0 | 312.1 |
| 12 | 290.0 | 163.0 | 204.0 | 243.0 | 274.2 | 334.3 |
| 13 | 309.0 | 172.8 | 220.8 | 254.0 | 288.0 | 337.1 |
| 14 | 334.0 | 184.9 | 228.0 | 258.0 | 288.0 | 353.3 |
| 15 | 319.0 | 187.9 | 224.2 | 255.5 | 287.2 | 348.1 |
| 16 | 269.0 | 181.0 | 219.0 | 242.5 | 280.0 | 338.1 |
| 17 | 251.0 | 172.0 | 204.8 | 232.0 | 261.0 | 302.1 |
| 18 | 212.0 | 153.9 | 191.8 | 214.0 | 244.0 | 290.1 |
| 19 | 154.0 | 134.9 | 168.8 | 193.5 | 215.2 | 259.2 |
| 20 | 154.0 | 116.0 | 146.0 | 168.0 | 197.0 | 231.2 |
| 21 | 124.0 | 89.0 | 119.0 | 144.0 | 170.2 | 211.2 |
| 22 | 100.0 | 74.8 | 96.8 | 122.0 | 146.0 | 178.0 |
| 23 | 73.0 | 52.0 | 76.8 | 101.0 | 124.0 | 159.1 |
| 24 | 84.0 | 39.0 | 58.8 | 82.5 | 106.0 | 134.1 |
| 25 | 87.0 | 25.0 | 45.0 | 63.5 | 88.0 | 111.0 |
| 26 | 74.0 | 18.0 | 32.0 | 50.0 | 73.0 | 90.1 |
| 27 | 52.0 | 12.9 | 23.0 | 38.0 | 56.2 | 79.0 |
| 28 | 49.0 | 7.0 | 18.0 | 29.0 | 46.2 | 66.0 |
| 29 | 49.0 | 5.0 | 12.0 | 21.0 | 35.2 | 57.1 |
| 30 | 38.0 | 3.0 | 8.0 | 16.0 | 28.0 | 49.0 |
| 31 | 32.0 | 2.0 | 6.0 | 13.0 | 23.0 | 40.0 |
| 32 | 27.0 | 1.0 | 4.0 | 9.0 | 19.2 | 33.1 |
| 33 | 24.0 | 0.0 | 3.0 | 7.0 | 15.0 | 28.0 |
| 34 | 18.0 | 0.0 | 2.0 | 5.0 | 12.2 | 28.0 |
| 35 | 16.0 | 0.0 | 1.0 | 4.0 | 10.2 | 23.0 |
| 36 | 15.0 | 0.0 | 1.0 | 3.5 | 9.2 | 22.1 |
| 37 | 11.0 | 0.0 | 1.0 | 3.0 | 8.0 | 20.1 |
| 38 | 12.0 | 0.0 | 0.0 | 2.0 | 7.0 | 19.0 |

Table S28: **Weekly calibration – Slovenia.** Observed weekly hospitalisation counts and the 5th, 25th, 50th (median), 75th, and 95th percentiles of the simulated distributions across the 300 retained simulations.

| **Week** | **Observed** | **Sim Q05** | **Sim Q25** | **Sim Median** | **Sim Q75** | **Sim Q95** |
|---|---|---|---|---|---|---|
| 0 | 1.0 | 2.0 | 5.0 | 8.0 | 13.0 | 20.0 |
| 1 | 2.0 | 4.0 | 9.8 | 14.0 | 20.0 | 31.0 |
| 2 | 5.0 | 5.0 | 11.0 | 16.5 | 24.0 | 33.1 |
| 3 | 10.0 | 7.0 | 15.0 | 22.0 | 27.0 | 38.1 |
| 4 | 8.0 | 9.0 | 18.0 | 26.0 | 34.0 | 45.0 |
| 5 | 13.0 | 13.0 | 21.0 | 30.0 | 39.0 | 53.1 |
| 6 | 22.0 | 17.0 | 28.0 | 36.0 | 47.0 | 65.0 |
| 7 | 32.0 | 20.9 | 33.0 | 43.0 | 55.2 | 69.1 |
| 8 | 36.0 | 27.0 | 39.0 | 52.0 | 65.0 | 83.1 |
| 9 | 62.0 | 34.0 | 46.8 | 59.0 | 72.0 | 94.0 |
| 10 | 82.0 | 38.0 | 53.0 | 67.5 | 83.2 | 103.0 |
| 11 | 93.0 | 43.0 | 60.0 | 75.0 | 90.0 | 112.2 |
| 12 | 120.0 | 48.0 | 67.0 | 80.0 | 95.0 | 127.1 |
| 13 | 105.0 | 54.0 | 71.0 | 86.0 | 101.0 | 131.0 |
| 14 | 129.0 | 54.0 | 74.0 | 88.0 | 105.0 | 130.1 |
| 15 | 115.0 | 58.0 | 75.8 | 88.0 | 104.0 | 137.0 |
| 16 | 115.0 | 56.0 | 75.0 | 87.0 | 103.2 | 126.0 |
| 17 | 81.0 | 55.0 | 71.0 | 83.0 | 97.2 | 122.0 |
| 18 | 69.0 | 48.0 | 66.0 | 79.0 | 93.0 | 117.0 |
| 19 | 55.0 | 45.0 | 58.0 | 70.0 | 85.2 | 109.0 |
| 20 | 52.0 | 39.0 | 51.0 | 63.0 | 77.0 | 100.0 |
| 21 | 48.0 | 34.0 | 45.0 | 55.0 | 68.0 | 87.1 |
| 22 | 50.0 | 25.0 | 38.8 | 49.0 | 59.0 | 76.1 |
| 23 | 33.0 | 20.0 | 30.0 | 39.0 | 50.0 | 64.0 |
| 24 | 25.0 | 13.0 | 24.0 | 32.0 | 43.0 | 56.0 |
| 25 | 34.0 | 11.0 | 19.0 | 26.0 | 35.0 | 49.1 |
| 26 | 18.0 | 6.0 | 13.0 | 19.0 | 29.0 | 44.0 |
| 27 | 40.0 | 4.0 | 10.0 | 15.0 | 23.0 | 34.1 |
| 28 | 21.0 | 2.0 | 6.0 | 11.0 | 18.0 | 33.1 |
| 29 | 22.0 | 2.0 | 5.0 | 8.0 | 14.0 | 27.0 |
| 30 | 17.0 | 0.0 | 3.0 | 6.0 | 12.0 | 24.0 |
| 31 | 10.0 | 0.0 | 2.0 | 4.0 | 9.0 | 20.0 |
| 32 | 11.0 | 0.0 | 2.0 | 4.0 | 8.0 | 17.0 |
| 33 | 7.0 | 0.0 | 1.0 | 2.0 | 6.0 | 13.0 |
| 34 | 5.0 | 0.0 | 0.0 | 2.0 | 5.0 | 12.1 |
| 35 | 5.0 | 0.0 | 0.0 | 1.0 | 4.0 | 12.0 |
| 36 | 3.0 | 0.0 | 0.0 | 1.0 | 4.0 | 11.0 |
| 37 | 5.0 | 0.0 | 0.0 | 0.0 | 2.0 | 12.0 |
| 38 | 3.0 | 0.0 | 0.0 | 1.0 | 3.0 | 10.0 |

Table S29: **Weekly calibration – Spain.** Observed weekly hospitalisation counts and the 5th, 25th, 50th (median), 75th, and 95th percentiles of the simulated distributions across the 300 retained simulations.

| Week | Observed | Sim Q05 | Sim Q25 | Sim Median | Sim Q75 | Sim Q95 |
|---|---|---|---|---|---|---|
| 0 | 45.0 | 16.0 | 34.0 | 50.0 | 67.0 | 113.0 |
| 1 | 63.0 | 36.9 | 75.0 | 100.5 | 129.0 | 219.1 |
| 2 | 73.0 | 55.0 | 115.8 | 146.5 | 185.2 | 311.4 |
| 3 | 120.0 | 82.6 | 169.0 | 209.5 | 259.0 | 422.2 |
| 4 | 130.0 | 131.2 | 245.0 | 296.0 | 363.5 | 540.4 |
| 5 | 262.0 | 191.7 | 342.5 | 415.5 | 485.0 | 707.3 |
| 6 | 376.0 | 287.2 | 466.2 | 560.0 | 666.2 | 884.0 |
| 7 | 603.0 | 415.4 | 611.5 | 731.0 | 878.2 | 1,124.1 |
| 8 | 782.0 | 558.8 | 780.8 | 940.0 | 1,104.5 | 1,359.2 |
| 9 | 1,151.0 | 764.0 | 962.0 | 1,138.0 | 1,326.0 | 1,578.0 |
| 10 | 1,474.0 | 960.5 | 1,143.2 | 1,315.0 | 1,540.2 | 1,815.6 |
| 11 | 1,742.0 | 1,144.0 | 1,307.0 | 1,482.0 | 1,678.2 | 1,965.3 |
| 12 | 1,998.0 | 1,269.3 | 1,439.8 | 1,591.0 | 1,815.0 | 2,091.1 |
| 13 | 2,030.0 | 1,344.8 | 1,520.5 | 1,677.5 | 1,877.8 | 2,145.6 |
| 14 | 2,079.0 | 1,378.8 | 1,552.8 | 1,718.5 | 1,934.5 | 2,171.5 |
| 15 | 2,008.0 | 1,349.6 | 1,529.8 | 1,669.5 | 1,887.0 | 2,099.3 |
| 16 | 1,763.0 | 1,288.9 | 1,470.8 | 1,623.5 | 1,805.5 | 2,041.0 |
| 17 | 1,624.0 | 1,195.9 | 1,370.5 | 1,530.5 | 1,696.2 | 1,931.1 |
| 18 | 1,247.0 | 1,028.2 | 1,222.0 | 1,403.5 | 1,549.0 | 1,768.2 |
| 19 | 956.0 | 852.9 | 1,054.8 | 1,228.5 | 1,402.2 | 1,625.6 |
| 20 | 907.0 | 695.4 | 872.2 | 1,053.0 | 1,235.0 | 1,442.0 |
| 21 | 695.0 | 537.0 | 714.8 | 895.0 | 1,062.0 | 1,254.4 |
| 22 | 664.0 | 411.9 | 578.5 | 735.0 | 897.8 | 1,091.3 |
| 23 | 566.0 | 301.9 | 447.8 | 575.0 | 736.2 | 911.1 |
| 24 | 584.0 | 208.9 | 324.2 | 450.5 | 592.2 | 781.2 |
| 25 | 462.0 | 141.9 | 235.8 | 351.5 | 466.2 | 649.1 |
| 26 | 358.0 | 100.0 | 172.2 | 266.5 | 378.2 | 536.0 |
| 27 | 311.0 | 70.0 | 125.8 | 196.0 | 306.2 | 439.5 |
| 28 | 295.0 | 46.0 | 88.0 | 141.5 | 231.8 | 359.1 |
| 29 | 288.0 | 31.0 | 62.8 | 104.5 | 177.2 | 302.5 |
| 30 | 245.0 | 19.0 | 43.8 | 76.0 | 142.0 | 255.3 |
| 31 | 198.0 | 13.9 | 30.8 | 57.5 | 110.2 | 222.2 |
| 32 | 166.0 | 10.0 | 21.0 | 43.0 | 89.2 | 183.1 |
| 33 | 155.0 | 6.0 | 15.0 | 33.0 | 68.0 | 175.1 |
| 34 | 132.0 | 4.0 | 11.0 | 26.0 | 57.2 | 154.0 |
| 35 | 94.0 | 2.0 | 8.8 | 20.0 | 48.0 | 148.2 |
| 36 | 96.0 | 2.0 | 6.0 | 17.0 | 41.2 | 143.1 |
| 37 | 85.0 | 1.0 | 4.0 | 14.0 | 37.2 | 145.1 |
| 38 | 66.0 | 1.0 | 4.0 | 12.0 | 37.0 | 154.3 |

Table S30: **Weekly calibration – Sweden.** Observed weekly hospitalisation counts and the 5th, 25th, 50th (median), 75th, and 95th percentiles of the simulated distributions across the 300 retained simulations.

| Week | Observed | Sim Q05 | Sim Q25 | Sim Median | Sim Q75 | Sim Q95 |
|---|---|---|---|---|---|---|
| 0 | 18.0 | 7.0 | 13.0 | 29.0 | 44.0 | 73.0 |
| 1 | 20.0 | 12.0 | 26.0 | 46.5 | 73.0 | 112.0 |
| 2 | 22.0 | 19.0 | 35.8 | 58.5 | 98.2 | 139.0 |
| 3 | 29.0 | 30.0 | 51.8 | 74.0 | 128.0 | 175.1 |
| 4 | 50.0 | 44.9 | 70.0 | 97.5 | 158.0 | 222.2 |
| 5 | 81.0 | 66.8 | 96.8 | 127.0 | 192.0 | 269.1 |
| 6 | 134.0 | 88.0 | 131.5 | 168.5 | 240.2 | 318.3 |
| 7 | 161.0 | 122.8 | 174.0 | 219.0 | 288.0 | 389.3 |
| 8 | 288.0 | 165.9 | 224.8 | 283.5 | 342.2 | 444.3 |
| 9 | 355.0 | 210.9 | 281.0 | 345.0 | 406.2 | 519.0 |
| 10 | 478.0 | 263.9 | 335.0 | 397.0 | 457.2 | 570.3 |
| 11 | 548.0 | 317.0 | 388.2 | 454.0 | 518.0 | 599.4 |
| 12 | 602.0 | 360.9 | 431.0 | 488.5 | 560.0 | 649.2 |
| 13 | 660.0 | 393.9 | 461.8 | 508.5 | 588.0 | 692.5 |
| 14 | 669.0 | 408.9 | 468.8 | 524.0 | 590.0 | 707.1 |
| 15 | 598.0 | 406.0 | 460.0 | 514.5 | 585.0 | 720.2 |
| 16 | 545.0 | 376.9 | 434.8 | 504.0 | 564.0 | 682.0 |
| 17 | 483.0 | 348.0 | 401.5 | 466.0 | 533.2 | 636.5 |
| 18 | 390.0 | 310.9 | 361.8 | 420.0 | 490.2 | 565.0 |
| 19 | 274.0 | 259.9 | 315.8 | 366.0 | 439.0 | 517.2 |
| 20 | 236.0 | 211.9 | 273.8 | 317.5 | 384.0 | 461.0 |
| 21 | 231.0 | 168.0 | 220.8 | 268.0 | 327.0 | 404.0 |
| 22 | 206.0 | 128.0 | 177.8 | 216.5 | 272.2 | 334.1 |
| 23 | 170.0 | 104.0 | 143.0 | 177.0 | 221.2 | 275.1 |
| 24 | 161.0 | 71.0 | 107.0 | 140.0 | 180.2 | 236.1 |
| 25 | 140.0 | 46.0 | 81.0 | 108.0 | 145.5 | 193.0 |
| 26 | 135.0 | 36.0 | 56.8 | 84.5 | 109.8 | 157.1 |
| 27 | 112.0 | 23.0 | 42.8 | 62.0 | 85.0 | 133.2 |
| 28 | 75.0 | 17.0 | 30.0 | 46.0 | 67.0 | 107.0 |
| 29 | 90.0 | 10.0 | 22.0 | 36.0 | 52.0 | 90.0 |
| 30 | 80.0 | 7.0 | 15.8 | 24.0 | 42.0 | 76.1 |
| 31 | 59.0 | 4.0 | 10.0 | 18.0 | 33.2 | 65.1 |
| 32 | 43.0 | 3.0 | 7.0 | 14.0 | 26.0 | 55.0 |
| 33 | 44.0 | 2.0 | 5.0 | 11.0 | 22.0 | 49.1 |
| 34 | 44.0 | 1.0 | 4.0 | 9.0 | 19.2 | 46.0 |
| 35 | 31.0 | 0.0 | 3.0 | 6.5 | 15.0 | 38.2 |
| 36 | 21.0 | 0.0 | 2.0 | 5.0 | 13.2 | 37.0 |
| 37 | 18.0 | 0.0 | 1.0 | 5.0 | 12.0 | 36.0 |
| 38 | 31.0 | 0.0 | 1.0 | 4.0 | 12.0 | 36.0 |

##### S4.2.2 Age-stratified hospitalisation calibration tables

Table S31: **Age-stratified calibration – Austria.** Observed cumulative hospitalisations by age group over the season, and the 5th, 25th, 50th (median), 75th, and 95th percentiles of the simulated distributions across the 300 retained simulations.

| Age group | Observed | Sim Q05 | Sim Q25 | Sim Median | Sim Q75 | Sim Q95 |
|---|---|---|---|---|---|---|
| 0–2m | 1,330 | 923 | 1,162 | 1,330 | 1,497 | 1,774 |
| 3–5m | 650 | 427 | 546 | 637 | 722 | 828 |
| 6–11m | 546 | 331 | 445 | 530 | 597 | 697 |
| 1–4y | 881 | 393 | 652 | 773 | 940 | 1,244 |
| 5–64y | 295 | 132 | 225 | 281 | 338 | 403 |
| 65+ | 2,678 | 1,544 | 2,078 | 2,528 | 2,851 | 3,415 |

Table S32: **Age-stratified calibration – Belgium.** Observed cumulative hospitalisations by age group over the season, and the 5th, 25th, 50th (median), 75th, and 95th percentiles of the simulated distributions across the 300 retained simulations.

| Age group | Observed | Sim Q05 | Sim Q25 | Sim Median | Sim Q75 | Sim Q95 |
|---|---|---|---|---|---|---|
| 0–2m | 1,942 | 1,494 | 1,796 | 2,040 | 2,276 | 2,761 |
| 3–5m | 1,048 | 658 | 816 | 972 | 1,093 | 1,314 |
| 6–11m | 888 | 494 | 693 | 836 | 946 | 1,094 |
| 1–4y | 1,392 | 762 | 1,064 | 1,299 | 1,521 | 1,849 |
| 5–64y | 395 | 134 | 304 | 398 | 468 | 616 |
| 65+ | 3,705 | 2,100 | 2,982 | 3,686 | 4,322 | 5,390 |

Table S33: **Age-stratified calibration – Bulgaria.** Observed cumulative hospitalisations by age group over the season, and the 5th, 25th, 50th (median), 75th, and 95th percentiles of the simulated distributions across the 300 retained simulations.

| Age group | Observed | Sim Q05 | Sim Q25 | Sim Median | Sim Q75 | Sim Q95 |
|---|---|---|---|---|---|---|
| 0–2m | 1,154 | 845 | 1,006 | 1,134 | 1,262 | 1,463 |
| 3–5m | 649 | 458 | 550 | 634 | 706 | 824 |
| 6–11m | 567 | 326 | 436 | 528 | 616 | 747 |
| 1–4y | 748 | 370 | 551 | 691 | 800 | 1,075 |
| 5–64y | 231 | 112 | 177 | 218 | 280 | 360 |
| 65+ | 2,268 | 1,264 | 1,757 | 2,152 | 2,450 | 3,035 |

Table S34: **Age-stratified calibration – Croatia.** Observed cumulative hospitalisations by age group over the season, and the 5th, 25th, 50th (median), 75th, and 95th percentiles of the simulated distributions across the 300 retained simulations.

| **Age group** | **Observed** | **Sim Q05** | **Sim Q25** | **Sim Median** | **Sim Q75** | **Sim Q95** |
|---|---|---|---|---|---|---|
| 0–2m | 715 | 421 | 554 | 644 | 751 | 902 |
| 3–5m | 356 | 241 | 301 | 358 | 411 | 513 |
| 6–11m | 320 | 162 | 214 | 280 | 338 | 430 |
| 1–4y | 393 | 135 | 244 | 333 | 430 | 590 |
| 5–64y | 100 | 37 | 69 | 92 | 129 | 183 |
| 65+ | 1,154 | 622 | 960 | 1,198 | 1,468 | 1,968 |

Table S35: **Age-stratified calibration – Cyprus.** Observed cumulative hospitalisations by age group over the season, and the 5th, 25th, 50th (median), 75th, and 95th percentiles of the simulated distributions across the 300 retained simulations.

| **Age group** | **Observed** | **Sim Q05** | **Sim Q25** | **Sim Median** | **Sim Q75** | **Sim Q95** |
|---|---|---|---|---|---|---|
| 0–2m | 194 | 104 | 145 | 173 | 205 | 273 |
| 3–5m | 86 | 55 | 73 | 88 | 102 | 133 |
| 6–11m | 84 | 37 | 57 | 71 | 88 | 112 |
| 1–4y | 101 | 34 | 63 | 88 | 114 | 164 |
| 5–64y | 30 | 9 | 19 | 28 | 38 | 59 |
| 65+ | 252 | 110 | 184 | 248 | 322 | 427 |

Table S36: **Age-stratified calibration – Czechia.** Observed cumulative hospitalisations by age group over the season, and the 5th, 25th, 50th (median), 75th, and 95th percentiles of the simulated distributions across the 300 retained simulations.

| **Age group** | **Observed** | **Sim Q05** | **Sim Q25** | **Sim Median** | **Sim Q75** | **Sim Q95** |
|---|---|---|---|---|---|---|
| 0–2m | 1,839 | 1,329 | 1,645 | 1,882 | 2,085 | 2,544 |
| 3–5m | 948 | 713 | 858 | 960 | 1,085 | 1,268 |
| 6–11m | 928 | 617 | 755 | 883 | 1,005 | 1,189 |
| 1–4y | 1,510 | 679 | 1,034 | 1,326 | 1,578 | 1,958 |
| 5–64y | 315 | 145 | 248 | 306 | 375 | 475 |
| 65+ | 2,995 | 1,791 | 2,420 | 2,868 | 3,304 | 3,986 |

Table S37: **Age-stratified calibration – Denmark.** Observed cumulative hospitalisations by age group over the season, and the 5th, 25th, 50th (median), 75th, and 95th percentiles of the simulated distributions across the 300 retained simulations.

| Age group | Observed | Sim Q05 | Sim Q25 | Sim Median | Sim Q75 | Sim Q95 |
|---|---|---|---|---|---|---|
| 0–2m | 896 | 725 | 842 | 952 | 1,086 | 1,257 |
| 3–5m | 575 | 402 | 499 | 568 | 640 | 761 |
| 6–11m | 545 | 324 | 442 | 514 | 579 | 675 |
| 1–4y | 1,111 | 418 | 675 | 882 | 1,069 | 1,383 |
| 5–64y | 165 | 72 | 132 | 164 | 204 | 272 |
| 65+ | 1,546 | 933 | 1,293 | 1,543 | 1,824 | 2,301 |

Table S38: **Age-stratified calibration – Estonia.** Observed cumulative hospitalisations by age group over the season, and the 5th, 25th, 50th (median), 75th, and 95th percentiles of the simulated distributions across the 300 retained simulations.

| Age group | Observed | Sim Q05 | Sim Q25 | Sim Median | Sim Q75 | Sim Q95 |
|---|---|---|---|---|---|---|
| 0–2m | 192 | 129 | 172 | 193 | 226 | 293 |
| 3–5m | 127 | 66 | 85 | 108 | 131 | 169 |
| 6–11m | 98 | 45 | 69 | 87 | 104 | 137 |
| 1–4y | 170 | 64 | 108 | 141 | 188 | 251 |
| 5–64y | 42 | 16 | 29 | 41 | 55 | 77 |
| 65+ | 376 | 158 | 267 | 366 | 454 | 605 |

Table S39: **Age-stratified calibration – Finland.** Observed cumulative hospitalisations by age group over the season, and the 5th, 25th, 50th (median), 75th, and 95th percentiles of the simulated distributions across the 300 retained simulations.

| Age group | Observed | Sim Q05 | Sim Q25 | Sim Median | Sim Q75 | Sim Q95 |
|---|---|---|---|---|---|---|
| 0–2m | 914 | 597 | 774 | 872 | 1,000 | 1,242 |
| 3–5m | 455 | 304 | 377 | 453 | 521 | 642 |
| 6–11m | 361 | 212 | 289 | 341 | 396 | 495 |
| 1–4y | 571 | 107 | 310 | 453 | 568 | 742 |
| 5–64y | 51 | 25 | 37 | 52 | 64 | 88 |
| 65+ | 1,242 | 735 | 1,022 | 1,236 | 1,538 | 1,926 |

Table S40: **Age-stratified calibration – France.** Observed cumulative hospitalisations by age group over the season, and the 5th, 25th, 50th (median), 75th, and 95th percentiles of the simulated distributions across the 300 retained simulations.

| Age group | Observed | Sim Q05 | Sim Q25 | Sim Median | Sim Q75 | Sim Q95 |
|---|---|---|---|---|---|---|
| 0–2m | 16,393 | 11,635 | 13,966 | 15,659 | 17,093 | 19,770 |
| 3–5m | 7,975 | 6,301 | 7,381 | 8,193 | 9,302 | 11,404 |
| 6–11m | 8,634 | 4,333 | 6,468 | 7,476 | 8,574 | 10,061 |
| 1–4y | 8,332 | 4,567 | 6,646 | 7,860 | 9,117 | 11,217 |
| 5–64y | 1,955 | 1,159 | 1,658 | 1,989 | 2,228 | 2,741 |
| 65+ | 20,189 | 13,381 | 16,704 | 19,544 | 22,496 | 26,446 |

Table S41: **Age-stratified calibration – Germany.** Observed cumulative hospitalisations by age group over the season, and the 5th, 25th, 50th (median), 75th, and 95th percentiles of the simulated distributions across the 300 retained simulations.

| Age group | Observed | Sim Q05 | Sim Q25 | Sim Median | Sim Q75 | Sim Q95 |
|---|---|---|---|---|---|---|
| 0–2m | 13,281 | 9,938 | 12,174 | 13,535 | 15,160 | 17,871 |
| 3–5m | 6,585 | 4,939 | 6,028 | 6,768 | 7,565 | 8,759 |
| 6–11m | 6,115 | 3,814 | 5,073 | 5,934 | 6,665 | 7,769 |
| 1–4y | 9,572 | 4,651 | 6,870 | 8,388 | 9,664 | 12,396 |
| 5–64y | 1,812 | 961 | 1,470 | 1,797 | 2,062 | 2,624 |
| 65+ | 24,876 | 14,469 | 19,900 | 23,117 | 26,178 | 30,190 |

Table S42: **Age-stratified calibration – Greece.** Observed cumulative hospitalisations by age group over the season, and the 5th, 25th, 50th (median), 75th, and 95th percentiles of the simulated distributions across the 300 retained simulations.

| Age group | Observed | Sim Q05 | Sim Q25 | Sim Median | Sim Q75 | Sim Q95 |
|---|---|---|---|---|---|---|
| 0–2m | 1,570 | 1,156 | 1,407 | 1,576 | 1,770 | 2,034 |
| 3–5m | 803 | 542 | 684 | 800 | 891 | 1,060 |
| 6–11m | 733 | 445 | 586 | 692 | 792 | 966 |
| 1–4y | 848 | 411 | 634 | 788 | 933 | 1,139 |
| 5–64y | 323 | 149 | 243 | 311 | 374 | 498 |
| 65+ | 3,679 | 2,287 | 3,019 | 3,523 | 4,085 | 4,953 |

Table S43: **Age-stratified calibration – Hungary.** Observed cumulative hospitalisations by age group over the season, and the 5th, 25th, 50th (median), 75th, and 95th percentiles of the simulated distributions across the 300 retained simulations.

| Age group | Observed | Sim Q05 | Sim Q25 | Sim Median | Sim Q75 | Sim Q95 |
|---|---|---|---|---|---|---|
| 0–2m | 1,812 | 1,334 | 1,532 | 1,741 | 1,954 | 2,311 |
| 3–5m | 932 | 679 | 826 | 933 | 1,063 | 1,261 |
| 6–11m | 850 | 547 | 688 | 807 | 907 | 1,050 |
| 1–4y | 1,190 | 599 | 844 | 1,046 | 1,237 | 1,520 |
| 5–64y | 267 | 142 | 215 | 263 | 308 | 410 |
| 65+ | 2,593 | 1,791 | 2,169 | 2,576 | 2,934 | 3,650 |

Table S44: **Age-stratified calibration – Ireland.** Observed cumulative hospitalisations by age group over the season, and the 5th, 25th, 50th (median), 75th, and 95th percentiles of the simulated distributions across the 300 retained simulations.

| Age group | Observed | Sim Q05 | Sim Q25 | Sim Median | Sim Q75 | Sim Q95 |
|---|---|---|---|---|---|---|
| 0–2m | 958 | 750 | 862 | 986 | 1,118 | 1,293 |
| 3–5m | 633 | 458 | 558 | 630 | 708 | 829 |
| 6–11m | 628 | 356 | 476 | 556 | 631 | 767 |
| 1–4y | 991 | 414 | 644 | 826 | 996 | 1,274 |
| 5–64y | 302 | 130 | 226 | 288 | 355 | 469 |
| 65+ | 1,298 | 796 | 1,101 | 1,312 | 1,544 | 1,919 |

Table S45: **Age-stratified calibration – Italy.** Observed cumulative hospitalisations by age group over the season, and the 5th, 25th, 50th (median), 75th, and 95th percentiles of the simulated distributions across the 300 retained simulations.

| Age group | Observed | Sim Q05 | Sim Q25 | Sim Median | Sim Q75 | Sim Q95 |
|---|---|---|---|---|---|---|
| 0–2m | 7,888 | 6,229 | 7,468 | 8,402 | 9,333 | 10,810 |
| 3–5m | 3,524 | 2,834 | 3,252 | 3,542 | 4,025 | 4,804 |
| 6–11m | 3,666 | 1,966 | 2,836 | 3,377 | 3,778 | 4,292 |
| 1–4y | 4,142 | 2,563 | 3,502 | 4,104 | 4,535 | 5,535 |
| 5–64y | 1,664 | 769 | 1,318 | 1,580 | 1,845 | 2,231 |
| 65+ | 19,732 | 12,076 | 15,686 | 17,918 | 20,301 | 24,111 |

Table S46: **Age-stratified calibration – Latvia.** Observed cumulative hospitalisations by age group over the season, and the 5th, 25th, 50th (median), 75th, and 95th percentiles of the simulated distributions across the 300 retained simulations.

| Age group | Observed | Sim Q05 | Sim Q25 | Sim Median | Sim Q75 | Sim Q95 |
|---|---|---|---|---|---|---|
| 0–2m | 283 | 165 | 234 | 278 | 327 | 380 |
| 3–5m | 166 | 89 | 121 | 154 | 180 | 229 |
| 6–11m | 123 | 56 | 94 | 116 | 140 | 177 |
| 1–4y | 222 | 52 | 116 | 164 | 222 | 294 |
| 5–64y | 33 | 10 | 20 | 30 | 40 | 61 |
| 65+ | 578 | 217 | 398 | 563 | 684 | 899 |

Table S47: **Age-stratified calibration – Lithuania.** Observed cumulative hospitalisations by age group over the season, and the 5th, 25th, 50th (median), 75th, and 95th percentiles of the simulated distributions across the 300 retained simulations.

| Age group | Observed | Sim Q05 | Sim Q25 | Sim Median | Sim Q75 | Sim Q95 |
|---|---|---|---|---|---|---|
| 0–2m | 422 | 283 | 361 | 418 | 469 | 580 |
| 3–5m | 223 | 139 | 184 | 212 | 251 | 306 |
| 6–11m | 202 | 97 | 140 | 178 | 207 | 270 |
| 1–4y | 215 | 95 | 151 | 196 | 241 | 312 |
| 5–64y | 69 | 28 | 45 | 65 | 84 | 112 |
| 65+ | 870 | 441 | 648 | 840 | 1,032 | 1,249 |

Table S48: **Age-stratified calibration – Luxembourg.** Observed cumulative hospitalisations by age group over the season, and the 5th, 25th, 50th (median), 75th, and 95th percentiles of the simulated distributions across the 300 retained simulations.

| Age group | Observed | Sim Q05 | Sim Q25 | Sim Median | Sim Q75 | Sim Q95 |
|---|---|---|---|---|---|---|
| 0–2m | 103 | 57 | 78 | 95 | 114 | 144 |
| 3–5m | 57 | 26 | 39 | 49 | 61 | 82 |
| 6–11m | 45 | 16 | 27 | 37 | 48 | 61 |
| 1–4y | 64 | 22 | 41 | 55 | 70 | 96 |
| 5–64y | 26 | 8 | 17 | 24 | 34 | 51 |
| 65+ | 140 | 86 | 129 | 158 | 198 | 256 |

Table S49: **Age-stratified calibration – Malta.** Observed cumulative hospitalisations by age group over the season, and the 5th, 25th, 50th (median), 75th, and 95th percentiles of the simulated distributions across the 300 retained simulations.

| Age group | Observed | Sim Q05 | Sim Q25 | Sim Median | Sim Q75 | Sim Q95 |
|---|---|---|---|---|---|---|
| 0–2m | 67 | 34 | 49 | 62 | 77 | 104 |
| 3–5m | 29 | 11 | 18 | 25 | 32 | 43 |
| 6–11m | 41 | 7 | 15 | 19 | 26 | 38 |
| 1–4y | 64 | 17 | 31 | 46 | 62 | 90 |
| 5–64y | 19 | 5 | 12 | 18 | 26 | 38 |
| 65+ | 156 | 99 | 146 | 196 | 249 | 326 |

Table S50: **Age-stratified calibration – Netherlands.** Observed cumulative hospitalisations by age group over the season, and the 5th, 25th, 50th (median), 75th, and 95th percentiles of the simulated distributions across the 300 retained simulations.

| Age group | Observed | Sim Q05 | Sim Q25 | Sim Median | Sim Q75 | Sim Q95 |
|---|---|---|---|---|---|---|
| 0–2m | 2,033 | 1,420 | 1,826 | 2,092 | 2,369 | 2,894 |
| 3–5m | 785 | 532 | 687 | 785 | 897 | 1,035 |
| 6–11m | 666 | 408 | 555 | 642 | 727 | 854 |
| 1–4y | 1,038 | 536 | 814 | 978 | 1,124 | 1,384 |
| 5–64y | 473 | 267 | 388 | 484 | 565 | 681 |
| 65+ | 4,183 | 2,676 | 3,360 | 3,906 | 4,394 | 5,356 |

Table S51: **Age-stratified calibration – Norway.** Observed cumulative hospitalisations by age group over the season, and the 5th, 25th, 50th (median), 75th, and 95th percentiles of the simulated distributions across the 300 retained simulations.

| Age group | Observed | Sim Q05 | Sim Q25 | Sim Median | Sim Q75 | Sim Q95 |
|---|---|---|---|---|---|---|
| 0–2m | 716 | 574 | 688 | 773 | 872 | 1,071 |
| 3–5m | 465 | 306 | 380 | 424 | 488 | 588 |
| 6–11m | 384 | 235 | 304 | 357 | 417 | 494 |
| 1–4y | 733 | 318 | 524 | 642 | 750 | 922 |
| 5–64y | 208 | 99 | 158 | 199 | 247 | 314 |
| 65+ | 1,831 | 1,033 | 1,380 | 1,732 | 1,959 | 2,409 |

Table S52: **Age-stratified calibration – Poland.** Observed cumulative hospitalisations by age group over the season, and the 5th, 25th, 50th (median), 75th, and 95th percentiles of the simulated distributions across the 300 retained simulations.

| Age group | Observed | Sim Q05 | Sim Q25 | Sim Median | Sim Q75 | Sim Q95 |
|---|---|---|---|---|---|---|
| 0–2m | 5,272 | 2,866 | 3,988 | 4,750 | 5,559 | 6,528 |
| 3–5m | 2,562 | 1,341 | 1,978 | 2,474 | 2,836 | 3,362 |
| 6–11m | 2,374 | 1,202 | 1,668 | 2,110 | 2,474 | 3,015 |
| 1–4y | 3,686 | 1,085 | 1,970 | 2,926 | 3,573 | 4,882 |
| 5–64y | 1,129 | 396 | 818 | 1,084 | 1,380 | 1,918 |
| 65+ | 10,008 | 6,507 | 9,158 | 11,250 | 13,427 | 16,972 |

Table S53: **Age-stratified calibration – Portugal.** Observed cumulative hospitalisations by age group over the season, and the 5th, 25th, 50th (median), 75th, and 95th percentiles of the simulated distributions across the 300 retained simulations.

| Age group | Observed | Sim Q05 | Sim Q25 | Sim Median | Sim Q75 | Sim Q95 |
|---|---|---|---|---|---|---|
| 0–2m | 1,449 | 976 | 1,247 | 1,412 | 1,620 | 2,008 |
| 3–5m | 619 | 445 | 540 | 603 | 694 | 834 |
| 6–11m | 602 | 359 | 477 | 561 | 626 | 728 |
| 1–4y | 963 | 450 | 722 | 892 | 1,078 | 1,373 |
| 5–64y | 285 | 131 | 209 | 270 | 332 | 420 |
| 65+ | 3,562 | 2,144 | 2,929 | 3,510 | 3,972 | 4,836 |

Table S54: **Age-stratified calibration – Romania.** Observed cumulative hospitalisations by age group over the season, and the 5th, 25th, 50th (median), 75th, and 95th percentiles of the simulated distributions across the 300 retained simulations.

| Age group | Observed | Sim Q05 | Sim Q25 | Sim Median | Sim Q75 | Sim Q95 |
|---|---|---|---|---|---|---|
| 0–2m | 2,959 | 2,163 | 2,630 | 2,994 | 3,301 | 3,893 |
| 3–5m | 1,567 | 1,210 | 1,444 | 1,588 | 1,761 | 2,134 |
| 6–11m | 1,642 | 934 | 1,250 | 1,484 | 1,657 | 1,905 |
| 1–4y | 1,879 | 1,097 | 1,527 | 1,798 | 2,067 | 2,446 |
| 5–64y | 830 | 430 | 665 | 800 | 905 | 1,127 |
| 65+ | 5,785 | 3,577 | 4,643 | 5,396 | 6,166 | 7,344 |

Table S55: **Age-stratified calibration – Slovakia.** Observed cumulative hospitalisations by age group over the season, and the 5th, 25th, 50th (median), 75th, and 95th percentiles of the simulated distributions across the 300 retained simulations.

| Age group | Observed | Sim Q05 | Sim Q25 | Sim Median | Sim Q75 | Sim Q95 |
|---|---|---|---|---|---|---|
| 0–2m | 986 | 728 | 857 | 968 | 1,102 | 1,332 |
| 3–5m | 529 | 373 | 466 | 526 | 600 | 729 |
| 6–11m | 505 | 260 | 381 | 451 | 524 | 629 |
| 1–4y | 614 | 296 | 467 | 566 | 678 | 850 |
| 5–64y | 140 | 65 | 107 | 137 | 159 | 218 |
| 65+ | 1,456 | 880 | 1,146 | 1,362 | 1,617 | 1,949 |

Table S56: **Age-stratified calibration – Slovenia.** Observed cumulative hospitalisations by age group over the season, and the 5th, 25th, 50th (median), 75th, and 95th percentiles of the simulated distributions across the 300 retained simulations.

| Age group | Observed | Sim Q05 | Sim Q25 | Sim Median | Sim Q75 | Sim Q95 |
|---|---|---|---|---|---|---|
| 0–2m | 361 | 228 | 287 | 340 | 388 | 468 |
| 3–5m | 192 | 119 | 155 | 182 | 214 | 254 |
| 6–11m | 157 | 85 | 116 | 145 | 168 | 216 |
| 1–4y | 240 | 79 | 150 | 201 | 244 | 323 |
| 5–64y | 43 | 15 | 30 | 42 | 55 | 78 |
| 65+ | 571 | 319 | 469 | 578 | 722 | 931 |

Table S57: **Age-stratified calibration – Spain.** Observed cumulative hospitalisations by age group over the season, and the 5th, 25th, 50th (median), 75th, and 95th percentiles of the simulated distributions across the 300 retained simulations.

| Age group | Observed | Sim Q05 | Sim Q25 | Sim Median | Sim Q75 | Sim Q95 |
|---|---|---|---|---|---|---|
| 0–2m | 5,818 | 4,231 | 5,345 | 5,881 | 6,666 | 7,990 |
| 3–5m | 2,453 | 1,850 | 2,245 | 2,534 | 2,826 | 3,352 |
| 6–11m | 2,731 | 1,232 | 1,936 | 2,362 | 2,716 | 3,144 |
| 1–4y | 2,581 | 1,448 | 2,048 | 2,559 | 2,952 | 3,707 |
| 5–64y | 1,053 | 524 | 836 | 977 | 1,190 | 1,470 |
| 65+ | 12,257 | 6,845 | 9,334 | 11,100 | 12,920 | 15,422 |

Table S58: **Age-stratified calibration – Sweden.** Observed cumulative hospitalisations by age group over the season, and the 5th, 25th, 50th (median), 75th, and 95th percentiles of the simulated distributions across the 300 retained simulations.

| Age group | Observed | Sim Q05 | Sim Q25 | Sim Median | Sim Q75 | Sim Q95 |
|---|---|---|---|---|---|---|
| 0–2m | 1,645 | 1,227 | 1,463 | 1,628 | 1,818 | 2,098 |
| 3–5m | 1,005 | 703 | 870 | 978 | 1,112 | 1,320 |
| 6–11m | 797 | 547 | 684 | 794 | 892 | 1,094 |
| 1–4y | 1,484 | 730 | 1,061 | 1,292 | 1,511 | 1,896 |
| 5–64y | 386 | 212 | 309 | 372 | 444 | 549 |
| 65+ | 3,015 | 1,759 | 2,454 | 2,855 | 3,276 | 3,960 |

### S5 Country-level averted hospitalisations

This section reports the country-level number of averted hospitalisations in infants under one year of age, expressed in both absolute and relative terms, for each intervention scenario and coverage level. For every combination of strategy (long-acting monoclonal antibodies, maternal vaccination, and the two with seasonal catch-up) and coverage level (25%, 50%, 75%, and 95%), we provide a table reporting the 5th, 25th, 50th (median), 75th, and 95th percentiles of the simulation-level distribution across the 300 retained simulations, for both metrics.

Absolute values can take negative values at the lower percentiles: this reflects the stochastic variability of paired baseline–intervention simulations and indicates that, for a fraction of simulations, the intervention did not result in a net reduction; the median across simulations remains positive in all country–scenario combinations. The percentage of hospitalisations averted is computed at the simulation level and is therefore unbounded below in principle; consistently with our outcome framing, we report aggregated percentiles so that the displayed values refer to non-negative reductions.

Tables are organised by strategy and, within each strategy, by decreasing coverage level. Countries are listed alphabetically.

#### S5.1 Long-acting monoclonal antibodies

Table S59: **Country-level averted hospitalisations under 1 year of age – la-mAbs,** 95% **coverage.** For each country, we report the 5th, 25th, 50th (median), 75th, and 95th percentiles of both absolute averted hospitalisations and the corresponding percentage, computed across the 300 retained simulations.

| | **Absolute averted** | | | | | **Percentage averted** | | | | |
|---|---|---|---|---|---|---|---|---|---|---|
| **Country** | **Q05** | **Q25** | **Median** | **Q75** | **Q95** | **Q05** | **Q25** | **Median** | **Q75** | **Q95** |
| Austria | 505 | 650 | 770 | 905 | 1 202 | 22.2% | 27.5% | 30.9% | 34.7% | 42.2% |
| Belgium | 754 | 972 | 1 149 | 1 356 | 1 702 | 23.0% | 27.1% | 30.1% | 33.3% | 38.2% |
| Bulgaria | 384 | 554 | 673 | 794 | 1 015 | 19.0% | 25.8% | 29.2% | 32.8% | 39.2% |
| Croatia | 149 | 310 | 408 | 522 | 720 | 14.5% | 26.2% | 31.7% | 37.1% | 46.5% |
| Cyprus | 20.9 | 71.8 | 97.5 | 137 | 209 | 6.2% | 21.8% | 30.8% | 38.4% | 52.1% |
| Czechia | 637 | 870 | 1 059 | 1 224 | 1 528 | 19.1% | 25.0% | 28.0% | 31.2% | 36.5% |
| Denmark | 302 | 465 | 565 | 686 | 852 | 17.1% | 24.4% | 28.2% | 31.6% | 38.1% |
| Estonia | 28.9 | 76.0 | 104 | 149 | 237 | 8.0% | 20.9% | 27.7% | 34.4% | 47.5% |
| Finland | 278 | 404 | 507 | 602 | 784 | 20.0% | 26.4% | 30.0% | 33.6% | 39.9% |
| France | 7 125 | 8 550 | 9 706 | 10 930 | 12 232 | 25.0% | 28.5% | 30.8% | 33.3% | 37.5% |
| Germany | 5 824 | 6 825 | 8 004 | 9 104 | 10 810 | 24.4% | 27.8% | 30.3% | 32.9% | 36.6% |
| Greece | 597 | 754 | 891 | 1 054 | 1 300 | 22.2% | 26.2% | 29.2% | 32.4% | 38.2% |
| Hungary | 674 | 908 | 1 058 | 1 226 | 1 519 | 22.0% | 26.8% | 30.3% | 33.7% | 39.1% |
| Ireland | 317 | 522 | 633 | 777 | 1 012 | 18.2% | 25.8% | 29.2% | 34.3% | 42.3% |
| Italy | 3 361 | 4 178 | 4 831 | 5 412 | 6 428 | 24.6% | 29.1% | 31.2% | 33.9% | 37.6% |
| Latvia | 18.2 | 106 | 155 | 196 | 305 | 4.8% | 21.5% | 28.4% | 33.9% | 52.0% |
| Lithuania | 92.0 | 174 | 223 | 277 | 411 | 12.9% | 23.2% | 27.7% | 33.1% | 44.3% |
| Luxembourg | 3.9 | 32.0 | 56.0 | 75.0 | 108 | 2.5% | 19.9% | 29.1% | 38.6% | 50.4% |
| Malta | 8.0 | 23.0 | 36.5 | 49.2 | 78.0 | 10.0% | 25.2% | 33.9% | 41.8% | 53.9% |
| Netherlands | 762 | 990 | 1 164 | 1 319 | 1 648 | 26.1% | 29.9% | 32.8% | 35.8% | 40.2% |
| Norway | 281 | 360 | 432 | 505 | 652 | 19.7% | 24.9% | 27.6% | 30.5% | 36.7% |
| Poland | 1 538 | 2 286 | 2 776 | 3 400 | 4 474 | 22.3% | 27.2% | 30.4% | 33.5% | 39.2% |
| Portugal | 504 | 672 | 794 | 936 | 1 239 | 23.2% | 27.5% | 30.6% | 34.8% | 40.2% |
| Romania | 1 131 | 1 405 | 1 726 | 2 003 | 2 452 | 21.0% | 25.1% | 28.2% | 31.2% | 37.3% |
| Slovakia | 303 | 452 | 570 | 676 | 874 | 18.3% | 24.1% | 29.0% | 32.9% | 39.8% |
| Slovenia | 77.8 | 156 | 200 | 250 | 337 | 13.3% | 24.2% | 29.9% | 35.2% | 45.2% |
| Spain | 2 337 | 2 910 | 3 337 | 3 896 | 4 623 | 24.7% | 28.2% | 30.9% | 33.9% | 37.3% |
| Sweden | 581 | 815 | 959 | 1 162 | 1 500 | 18.7% | 24.8% | 28.4% | 32.6% | 39.7% |

Table S60: **Country-level averted hospitalisations under 1 year of age – la-mAbs,** 75% **coverage.** For each country, we report the 5th, 25th, 50th (median), 75th, and 95th percentiles of both absolute averted hospitalisations and the corresponding percentage, computed across the 300 retained simulations.

| | Absolute averted | | | | | Percentage averted | | | | |
|---|---|---|---|---|---|---|---|---|---|---|
| **Country** | **Q05** | **Q25** | **Median** | **Q75** | **Q95** | **Q05** | **Q25** | **Median** | **Q75** | **Q95** |
| Austria | 345 | 502 | 600 | 722 | 962 | 15.3% | 21.4% | 24.1% | 27.3% | 34.8% |
| Belgium | 570 | 749 | 904 | 1 051 | 1 378 | 17.8% | 20.7% | 23.7% | 26.3% | 30.7% |
| Bulgaria | 229 | 416 | 537 | 650 | 864 | 10.8% | 19.2% | 22.9% | 26.9% | 35.0% |
| Croatia | 69.7 | 228 | 313 | 414 | 638 | 6.3% | 18.9% | 24.8% | 30.7% | 41.4% |
| Cyprus | -12.1 | 44.8 | 78.5 | 112 | 187 | 0.0% | 14.7% | 23.7% | 31.6% | 46.1% |
| Czechia | 444 | 672 | 826 | 987 | 1 308 | 13.2% | 19.2% | 22.2% | 25.6% | 30.8% |
| Denmark | 226 | 357 | 456 | 532 | 710 | 11.9% | 18.5% | 21.7% | 25.4% | 31.2% |
| Estonia | 12.0 | 52.8 | 78.5 | 123 | 191 | 3.5% | 14.5% | 20.6% | 27.8% | 39.1% |
| Finland | 184 | 300 | 396 | 501 | 696 | 12.4% | 19.5% | 23.5% | 28.2% | 35.6% |
| France | 5 557 | 6 670 | 7 522 | 8 552 | 9 717 | 19.4% | 22.1% | 24.2% | 26.2% | 29.5% |
| Germany | 4 401 | 5 396 | 6 188 | 7 228 | 8 397 | 18.6% | 21.6% | 23.9% | 26.1% | 28.7% |
| Greece | 434 | 601 | 705 | 848 | 1 131 | 15.8% | 20.4% | 23.1% | 26.5% | 32.0% |
| Hungary | 494 | 690 | 795 | 978 | 1 241 | 15.6% | 20.3% | 23.3% | 26.4% | 32.6% |
| Ireland | 260 | 404 | 495 | 628 | 882 | 12.8% | 19.5% | 23.5% | 27.5% | 36.0% |
| Italy | 2 585 | 3 300 | 3 816 | 4 254 | 5 099 | 19.6% | 22.7% | 24.8% | 26.8% | 29.7% |
| Latvia | -5.2 | 71.8 | 114 | 163 | 269 | 0.0% | 14.9% | 21.9% | 27.9% | 41.3% |
| Lithuania | 47.9 | 125 | 169 | 229 | 321 | 6.3% | 16.1% | 21.9% | 26.6% | 33.7% |
| Luxembourg | -0.1 | 28.0 | 45.5 | 65.0 | 94.1 | 0.0% | 16.5% | 24.3% | 32.3% | 43.3% |
| Malta | 4.0 | 15.0 | 28.0 | 44.0 | 63.0 | 4.5% | 17.9% | 26.7% | 35.0% | 47.5% |
| Netherlands | 552 | 770 | 898 | 1 062 | 1 418 | 18.7% | 23.0% | 25.8% | 28.8% | 34.4% |
| Norway | 196 | 279 | 342 | 403 | 531 | 13.8% | 18.7% | 21.7% | 24.6% | 29.9% |
| Poland | 1 147 | 1 771 | 2 198 | 2 717 | 3 490 | 16.9% | 20.7% | 23.9% | 26.8% | 31.0% |
| Portugal | 381 | 526 | 628 | 745 | 995 | 17.6% | 21.4% | 24.3% | 27.2% | 31.8% |
| Romania | 778 | 1 072 | 1 338 | 1 598 | 1 984 | 15.2% | 19.0% | 21.6% | 25.2% | 29.5% |
| Slovakia | 204 | 342 | 449 | 570 | 773 | 11.8% | 18.9% | 22.7% | 27.4% | 34.8% |
| Slovenia | 40.5 | 113 | 154 | 204 | 286 | 7.5% | 17.6% | 23.0% | 28.5% | 38.5% |
| Spain | 1 801 | 2 244 | 2 588 | 3 062 | 3 698 | 18.9% | 21.6% | 24.3% | 26.8% | 30.2% |
| Sweden | 416 | 625 | 738 | 904 | 1 157 | 12.7% | 18.7% | 22.0% | 25.2% | 31.6% |

Table S61: **Country-level averted hospitalisations under 1 year of age – la-mAbs,** 50% **coverage.** For each country, we report the 5th, 25th, 50th (median), 75th, and 95th percentiles of both absolute averted hospitalisations and the corresponding percentage, computed across the 300 retained simulations.

| | **Absolute averted** | | | | | **Percentage averted** | | | | |
|---|---|---|---|---|---|---|---|---|---|---|
| **Country** | **Q05** | **Q25** | **Median** | **Q75** | **Q95** | **Q05** | **Q25** | **Median** | **Q75** | **Q95** |
| Austria | 148 | 297 | 378 | 507 | 751 | 6.3% | 12.4% | 15.7% | 19.4% | 28.0% |
| Belgium | 355 | 480 | 594 | 713 | 948 | 10.3% | 13.5% | 15.8% | 18.1% | 21.2% |
| Bulgaria | 74.0 | 248 | 344 | 460 | 665 | 4.0% | 11.0% | 14.7% | 19.5% | 26.8% |
| Croatia | -33.4 | 128 | 218 | 308 | 529 | 0.0% | 11.1% | 17.0% | 23.7% | 34.8% |
| Cyprus | -53.0 | 23.0 | 52.0 | 86.2 | 153 | 0.0% | 7.1% | 16.3% | 23.8% | 44.2% |
| Czechia | 180 | 397 | 549 | 688 | 1 011 | 5.5% | 11.3% | 14.6% | 17.8% | 23.4% |
| Denmark | 94.0 | 216 | 296 | 387 | 557 | 5.1% | 11.4% | 14.6% | 17.7% | 24.7% |
| Estonia | -34.1 | 28.0 | 54.0 | 84.2 | 156 | 0.0% | 7.3% | 14.0% | 20.8% | 35.8% |
| Finland | 74.8 | 182 | 258 | 363 | 530 | 4.7% | 11.5% | 15.6% | 19.5% | 27.5% |
| France | 3 500 | 4 277 | 5 025 | 5 865 | 6 885 | 12.1% | 14.6% | 16.0% | 17.9% | 21.6% |
| Germany | 2 889 | 3 618 | 4 186 | 4 837 | 5 961 | 12.0% | 14.4% | 15.9% | 17.8% | 19.9% |
| Greece | 219 | 365 | 462 | 590 | 828 | 7.9% | 12.5% | 15.6% | 18.1% | 24.5% |
| Hungary | 240 | 431 | 545 | 687 | 1 016 | 7.4% | 12.6% | 16.1% | 19.3% | 24.8% |
| Ireland | 72.8 | 244 | 334 | 455 | 699 | 4.0% | 11.8% | 15.9% | 19.8% | 28.4% |
| Italy | 1 678 | 2 176 | 2 512 | 2 890 | 3 584 | 12.3% | 14.7% | 16.3% | 18.4% | 21.0% |
| Latvia | -80.0 | 43.0 | 82.0 | 126 | 226 | 0.0% | 8.6% | 14.4% | 22.3% | 37.5% |
| Lithuania | -14.1 | 67.0 | 110 | 158 | 263 | 0.0% | 8.7% | 14.0% | 19.6% | 28.2% |
| Luxembourg | -20.1 | 14.0 | 29.0 | 45.0 | 79.1 | 0.0% | 8.2% | 16.6% | 24.3% | 37.4% |
| Malta | -8.1 | 8.0 | 17.0 | 31.0 | 54.1 | 0.0% | 8.8% | 17.0% | 26.6% | 40.0% |
| Netherlands | 344 | 484 | 590 | 715 | 971 | 11.3% | 14.5% | 16.9% | 19.6% | 23.4% |
| Norway | 84.0 | 167 | 216 | 282 | 414 | 5.9% | 11.3% | 14.2% | 17.2% | 22.4% |
| Poland | 732 | 1 130 | 1 440 | 1 788 | 2 554 | 10.1% | 13.4% | 15.8% | 18.3% | 23.5% |
| Portugal | 213 | 324 | 422 | 520 | 715 | 9.5% | 13.1% | 16.3% | 19.1% | 24.0% |
| Romania | 435 | 658 | 884 | 1 131 | 1 560 | 7.6% | 11.5% | 14.4% | 18.0% | 23.5% |
| Slovakia | 72.6 | 214 | 298 | 387 | 555 | 3.7% | 11.5% | 14.9% | 19.0% | 26.4% |
| Slovenia | -22.0 | 58.2 | 99.0 | 145 | 213 | 0.0% | 9.0% | 14.7% | 20.6% | 29.9% |
| Spain | 1 134 | 1 481 | 1 711 | 2 031 | 2 592 | 11.7% | 14.2% | 16.1% | 18.1% | 20.9% |
| Sweden | 141 | 374 | 516 | 659 | 1 034 | 3.7% | 11.5% | 15.2% | 18.8% | 27.6% |

Table S62: **Country-level averted hospitalisations under 1 year of age – la-mAbs,** 25% **coverage.** For each country, we report the 5th, 25th, 50th (median), 75th, and 95th percentiles of both absolute averted hospitalisations and the corresponding percentage, computed across the 300 retained simulations.

| | **Absolute averted** | | | | | **Percentage averted** | | | | |
|---|---|---|---|---|---|---|---|---|---|---|
| **Country** | **Q05** | **Q25** | **Median** | **Q75** | **Q95** | **Q05** | **Q25** | **Median** | **Q75** | **Q95** |
| Austria | -47.9 | 112 | 198 | 278 | 534 | 0.0% | 4.8% | 8.0% | 11.1% | 20.1% |
| Belgium | 71.5 | 212 | 285 | 390 | 587 | 1.8% | 5.7% | 7.8% | 9.6% | 13.4% |
| Bulgaria | -182 | 75.8 | 158 | 258 | 438 | 0.0% | 3.4% | 7.1% | 11.1% | 18.8% |
| Croatia | -206 | 17.8 | 113 | 200 | 396 | 0.0% | 1.4% | 8.8% | 15.0% | 26.6% |
| Cyprus | -59.0 | -1.0 | 22.0 | 51.0 | 137 | 0.0% | 0.0% | 7.3% | 15.2% | 38.4% |
| Czechia | -88.2 | 158 | 270 | 392 | 644 | 0.0% | 4.4% | 7.0% | 10.3% | 16.0% |
| Denmark | -46.8 | 79.0 | 148 | 228 | 363 | 0.0% | 4.2% | 7.4% | 10.8% | 16.8% |
| Estonia | -69.2 | 3.8 | 30.0 | 56.2 | 142 | 0.0% | 0.9% | 7.6% | 13.5% | 26.9% |
| Finland | -67.4 | 59.0 | 128 | 208 | 430 | 0.0% | 3.9% | 8.1% | 11.5% | 23.1% |
| France | 1 301 | 1 959 | 2 430 | 3 048 | 4 391 | 4.2% | 6.5% | 7.9% | 9.5% | 13.4% |
| Germany | 1 118 | 1 696 | 2 110 | 2 586 | 3 475 | 4.6% | 6.7% | 8.1% | 9.4% | 12.1% |
| Greece | -45.5 | 155 | 239 | 328 | 540 | 0.0% | 5.3% | 8.1% | 10.2% | 17.8% |
| Hungary | -44.5 | 164 | 288 | 432 | 677 | 0.0% | 4.7% | 8.4% | 12.1% | 17.9% |
| Ireland | -112 | 85.8 | 168 | 280 | 537 | 0.0% | 4.0% | 7.8% | 12.4% | 22.7% |
| Italy | 584 | 962 | 1 272 | 1 566 | 2 022 | 4.0% | 6.8% | 8.3% | 9.6% | 12.3% |
| Latvia | -95.0 | 2.8 | 41.0 | 81.2 | 197 | 0.0% | 0.5% | 7.9% | 15.3% | 31.8% |
| Lithuania | -85.2 | 8.8 | 55.5 | 106 | 203 | 0.0% | 1.0% | 7.0% | 12.5% | 22.0% |
| Luxembourg | -51.1 | -5.0 | 12.0 | 31.0 | 65.2 | 0.0% | 0.0% | 6.4% | 15.5% | 29.8% |
| Malta | -22.1 | 0.0 | 8.0 | 20.0 | 40.0 | 0.0% | 0.0% | 8.1% | 18.6% | 30.1% |
| Netherlands | 68.8 | 200 | 294 | 390 | 592 | 2.1% | 6.0% | 8.6% | 11.0% | 15.2% |
| Norway | -48.0 | 53.2 | 106 | 160 | 267 | 0.0% | 3.4% | 7.2% | 9.9% | 14.8% |
| Poland | 135 | 496 | 690 | 948 | 1 799 | 1.5% | 5.9% | 7.8% | 10.0% | 17.7% |
| Portugal | 12.9 | 130 | 192 | 280 | 466 | 0.5% | 5.4% | 7.6% | 10.5% | 15.1% |
| Romania | -97.4 | 273 | 447 | 633 | 1 035 | 0.0% | 4.6% | 7.5% | 10.4% | 15.9% |
| Slovakia | -105 | 62.2 | 148 | 246 | 400 | 0.0% | 3.5% | 7.6% | 12.3% | 18.1% |
| Slovenia | -73.0 | 10.0 | 53.0 | 92.0 | 188 | 0.0% | 1.7% | 8.1% | 13.3% | 26.2% |
| Spain | 377 | 684 | 858 | 1 121 | 1 598 | 3.6% | 6.3% | 8.1% | 10.2% | 13.3% |
| Sweden | -134 | 121 | 241 | 398 | 639 | 0.0% | 3.8% | 7.2% | 10.9% | 18.4% |

#### S5.2 Long-acting monoclonal antibodies with seasonal catch-up

Table S63: **Country-level averted hospitalisations under 1 year of age – la-mAbs with catch-up, 95% coverage.** For each country, we report the 5th, 25th, 50th (median), 75th, and 95th percentiles of both absolute averted hospitalisations and the corresponding percentage, computed across the 300 retained simulations.

| | Absolute averted | | | | | Percentage averted | | | | |
|---|---|---|---|---|---|---|---|---|---|---|
| **Country** | **Q05** | **Q25** | **Median** | **Q75** | **Q95** | **Q05** | **Q25** | **Median** | **Q75** | **Q95** |
| Austria | 930 | 1 147 | 1 306 | 1 512 | 1 865 | 43.8% | 49.0% | 53.4% | 57.6% | 64.3% |
| Belgium | 1 303 | 1 664 | 1 905 | 2 203 | 2 690 | 41.0% | 45.8% | 50.3% | 54.4% | 59.3% |
| Bulgaria | 855 | 1 038 | 1 195 | 1 390 | 1 642 | 41.9% | 48.0% | 52.5% | 57.1% | 63.8% |
| Croatia | 419 | 572 | 691 | 839 | 1 087 | 40.4% | 48.8% | 53.9% | 59.9% | 68.0% |
| Cyprus | 86.0 | 139 | 169 | 210 | 293 | 31.5% | 44.2% | 51.5% | 59.1% | 72.3% |
| Czechia | 1 344 | 1 668 | 1 915 | 2 143 | 2 612 | 42.0% | 47.8% | 51.3% | 55.3% | 60.9% |
| Denmark | 696 | 890 | 1 043 | 1 176 | 1 442 | 40.6% | 46.9% | 50.9% | 54.8% | 62.1% |
| Estonia | 113 | 152 | 190 | 238 | 339 | 35.1% | 43.3% | 48.7% | 54.7% | 67.1% |
| Finland | 583 | 758 | 886 | 1 032 | 1 324 | 41.4% | 48.0% | 52.3% | 57.5% | 64.3% |
| France | 12 744 | 15 049 | 16 874 | 18 520 | 20 750 | 44.7% | 50.1% | 53.4% | 56.8% | 61.0% |
| Germany | 10 043 | 11 895 | 13 970 | 15 472 | 18 070 | 43.4% | 48.5% | 52.7% | 56.6% | 59.9% |
| Greece | 1 092 | 1 328 | 1 559 | 1 772 | 2 127 | 41.2% | 46.6% | 50.6% | 55.0% | 60.8% |
| Hungary | 1 356 | 1 622 | 1 871 | 2 102 | 2 440 | 42.9% | 49.2% | 53.4% | 57.5% | 62.2% |
| Ireland | 862 | 1 015 | 1 156 | 1 332 | 1 630 | 43.9% | 49.6% | 53.7% | 57.8% | 64.6% |
| Italy | 5 922 | 7 103 | 8 020 | 8 946 | 10 326 | 43.9% | 48.8% | 52.2% | 56.0% | 60.4% |
| Latvia | 154 | 224 | 274 | 337 | 446 | 35.5% | 45.1% | 50.5% | 57.8% | 70.0% |
| Lithuania | 257 | 339 | 409 | 485 | 628 | 36.9% | 45.7% | 50.6% | 56.5% | 63.9% |
| Luxembourg | 41.0 | 68.8 | 94.0 | 120 | 166 | 30.7% | 42.3% | 51.2% | 59.3% | 72.0% |
| Malta | 19.0 | 36.0 | 50.0 | 65.2 | 97.1 | 24.5% | 38.0% | 47.3% | 54.4% | 65.8% |
| Netherlands | 1 281 | 1 699 | 1 960 | 2 230 | 2 673 | 44.4% | 52.3% | 55.5% | 59.6% | 64.7% |
| Norway | 527 | 688 | 799 | 898 | 1 126 | 40.8% | 46.1% | 50.0% | 54.4% | 61.1% |
| Poland | 2 613 | 3 889 | 4 772 | 5 730 | 6 980 | 41.0% | 46.0% | 51.5% | 56.3% | 62.9% |
| Portugal | 903 | 1 119 | 1 318 | 1 524 | 1 882 | 40.5% | 46.1% | 50.7% | 55.7% | 61.3% |
| Romania | 2 325 | 2 702 | 3 202 | 3 540 | 4 083 | 42.3% | 48.1% | 52.5% | 56.3% | 60.3% |
| Slovakia | 706 | 885 | 1 026 | 1 189 | 1 424 | 41.7% | 47.8% | 52.6% | 57.1% | 63.3% |
| Slovenia | 217 | 287 | 341 | 402 | 501 | 39.1% | 46.4% | 51.3% | 56.4% | 65.8% |
| Spain | 3 996 | 4 882 | 5 539 | 6 422 | 7 467 | 42.4% | 47.9% | 51.5% | 55.4% | 60.5% |
| Sweden | 1 337 | 1 628 | 1 836 | 2 052 | 2 509 | 43.0% | 49.6% | 54.2% | 58.5% | 64.4% |

Table S64: **Country-level averted hospitalisations under 1 year of age – la-mAbs with catch-up, 75% coverage.** For each country, we report the 5th, 25th, 50th (median), 75th, and 95th percentiles of both absolute averted hospitalisations and the corresponding percentage, computed across the 300 retained simulations.

| | **Absolute averted** | | | | | **Percentage averted** | | | | |
|---|---|---|---|---|---|---|---|---|---|---|
| **Country** | **Q05** | **Q25** | **Median** | **Q75** | **Q95** | **Q05** | **Q25** | **Median** | **Q75** | **Q95** |
| Austria | 698 | 913 | 1 028 | 1 172 | 1 507 | 32.7% | 37.9% | 41.5% | 45.7% | 53.4% |
| Belgium | 1 020 | 1 303 | 1 527 | 1 753 | 2 211 | 32.1% | 36.5% | 39.9% | 43.2% | 49.0% |
| Bulgaria | 625 | 800 | 935 | 1 080 | 1 302 | 31.5% | 37.4% | 40.8% | 45.1% | 52.5% |
| Croatia | 273 | 449 | 536 | 674 | 902 | 26.4% | 36.8% | 42.7% | 48.3% | 58.4% |
| Cyprus | 60.0 | 103 | 136 | 173 | 256 | 22.8% | 33.5% | 40.5% | 49.1% | 62.6% |
| Czechia | 1 030 | 1 285 | 1 492 | 1 734 | 2 058 | 32.2% | 36.8% | 40.3% | 44.2% | 49.5% |
| Denmark | 541 | 696 | 825 | 946 | 1 143 | 31.4% | 36.7% | 39.8% | 43.8% | 50.6% |
| Estonia | 68.0 | 118 | 149 | 200 | 278 | 21.6% | 32.4% | 38.5% | 46.5% | 59.0% |
| Finland | 458 | 578 | 692 | 822 | 1 042 | 31.5% | 37.2% | 40.6% | 45.7% | 53.7% |
| France | 9 834 | 11 754 | 13 438 | 14 581 | 16 674 | 35.1% | 39.3% | 42.2% | 45.1% | 49.1% |
| Germany | 7 876 | 9 507 | 11 042 | 12 345 | 14 401 | 34.4% | 38.4% | 41.8% | 44.7% | 48.0% |
| Greece | 824 | 1 041 | 1 220 | 1 401 | 1 704 | 32.0% | 35.9% | 39.7% | 43.3% | 49.2% |
| Hungary | 1 051 | 1 259 | 1 448 | 1 676 | 2 022 | 31.5% | 38.5% | 42.4% | 45.8% | 51.2% |
| Ireland | 643 | 794 | 930 | 1 077 | 1 361 | 32.9% | 38.8% | 42.7% | 47.2% | 54.0% |
| Italy | 4 539 | 5 556 | 6 361 | 7 111 | 8 193 | 34.3% | 38.4% | 41.2% | 44.6% | 47.8% |
| Latvia | 89.6 | 177 | 219 | 276 | 371 | 20.6% | 34.0% | 40.6% | 46.9% | 58.9% |
| Lithuania | 192 | 259 | 318 | 387 | 530 | 26.5% | 35.5% | 40.1% | 44.6% | 54.4% |
| Luxembourg | 22.0 | 50.0 | 70.0 | 94.2 | 146 | 16.2% | 29.8% | 38.6% | 46.1% | 62.1% |
| Malta | 12.9 | 28.0 | 40.0 | 54.0 | 81.0 | 15.3% | 29.4% | 38.2% | 46.0% | 56.8% |
| Netherlands | 997 | 1 339 | 1 556 | 1 772 | 2 150 | 34.6% | 40.8% | 44.0% | 47.1% | 52.7% |
| Norway | 422 | 538 | 632 | 726 | 940 | 32.0% | 36.3% | 39.9% | 43.8% | 50.6% |
| Poland | 2 084 | 3 015 | 3 795 | 4 572 | 5 546 | 32.2% | 36.3% | 40.2% | 44.6% | 51.6% |
| Portugal | 711 | 871 | 1 031 | 1 210 | 1 539 | 31.3% | 36.5% | 39.6% | 43.7% | 49.5% |
| Romania | 1 753 | 2 156 | 2 500 | 2 807 | 3 301 | 33.2% | 37.4% | 41.1% | 44.8% | 50.0% |
| Slovakia | 529 | 678 | 816 | 958 | 1 183 | 30.1% | 36.7% | 41.2% | 46.1% | 52.4% |
| Slovenia | 151 | 212 | 268 | 322 | 411 | 26.4% | 34.6% | 39.8% | 45.0% | 55.1% |
| Spain | 3 134 | 3 856 | 4 362 | 5 025 | 5 912 | 32.9% | 37.8% | 40.6% | 43.5% | 48.5% |
| Sweden | 1 021 | 1 259 | 1 446 | 1 661 | 1 972 | 33.5% | 38.8% | 42.4% | 46.7% | 51.4% |

Table S65: **Country-level averted hospitalisations under 1 year of age – la-mAbs with catch-up, 50% coverage.** For each country, we report the 5th, 25th, 50th (median), 75th, and 95th percentiles of both absolute averted hospitalisations and the corresponding percentage, computed across the 300 retained simulations.

| | Absolute averted | | | | | Percentage averted | | | | |
|---|---|---|---|---|---|---|---|---|---|---|
| **Country** | **Q05** | **Q25** | **Median** | **Q75** | **Q95** | **Q05** | **Q25** | **Median** | **Q75** | **Q95** |
| Austria | 410 | 593 | 694 | 797 | 1 055 | 19.2% | 25.2% | 27.8% | 31.4% | 36.6% |
| Belgium | 669 | 858 | 992 | 1 190 | 1 511 | 20.3% | 23.6% | 26.3% | 29.3% | 33.8% |
| Bulgaria | 336 | 506 | 628 | 762 | 987 | 16.2% | 23.1% | 27.0% | 31.8% | 38.2% |
| Croatia | 78.0 | 268 | 359 | 466 | 700 | 7.4% | 22.1% | 28.4% | 34.1% | 44.3% |
| Cyprus | -6.1 | 60.8 | 93.0 | 124 | 210 | 0.0% | 19.3% | 27.4% | 35.4% | 52.8% |
| Czechia | 623 | 831 | 982 | 1 156 | 1 617 | 19.6% | 23.4% | 26.4% | 29.6% | 36.0% |
| Denmark | 327 | 440 | 543 | 641 | 836 | 18.5% | 23.3% | 26.6% | 30.2% | 36.9% |
| Estonia | 24.9 | 68.0 | 97.5 | 138 | 235 | 7.4% | 18.6% | 26.2% | 32.4% | 47.0% |
| Finland | 253 | 369 | 456 | 566 | 763 | 17.8% | 24.0% | 27.1% | 30.8% | 38.9% |
| France | 6 449 | 7 723 | 8 770 | 9 736 | 11 290 | 22.5% | 25.8% | 28.1% | 30.1% | 33.6% |
| Germany | 5 178 | 6 287 | 7 375 | 8 303 | 9 679 | 22.6% | 25.5% | 27.9% | 30.1% | 33.0% |
| Greece | 562 | 700 | 800 | 964 | 1 286 | 20.1% | 24.1% | 26.5% | 30.1% | 36.5% |
| Hungary | 638 | 806 | 959 | 1 128 | 1 441 | 19.4% | 24.7% | 27.6% | 31.1% | 38.1% |
| Ireland | 327 | 502 | 626 | 752 | 1 022 | 17.9% | 24.5% | 28.8% | 33.1% | 40.8% |
| Italy | 3 086 | 3 712 | 4 202 | 4 740 | 5 577 | 22.4% | 25.4% | 27.1% | 29.6% | 32.9% |
| Latvia | 16.8 | 103 | 144 | 187 | 312 | 3.9% | 19.7% | 26.3% | 31.7% | 49.4% |
| Lithuania | 83.8 | 158 | 213 | 283 | 383 | 12.2% | 21.1% | 26.2% | 32.8% | 41.8% |
| Luxembourg | 0.0 | 29.0 | 46.5 | 68.0 | 121 | 0.0% | 17.6% | 26.1% | 34.2% | 51.5% |
| Malta | -1.0 | 16.0 | 28.0 | 42.0 | 61.0 | 0.0% | 16.7% | 26.3% | 34.2% | 46.6% |
| Netherlands | 648 | 870 | 1 010 | 1 198 | 1 569 | 21.7% | 25.7% | 29.2% | 32.0% | 37.3% |
| Norway | 259 | 335 | 404 | 474 | 624 | 18.3% | 22.8% | 25.8% | 28.8% | 35.6% |
| Poland | 1 342 | 1 985 | 2 516 | 3 121 | 3 844 | 20.6% | 23.9% | 26.7% | 30.5% | 36.6% |
| Portugal | 448 | 568 | 688 | 829 | 1 101 | 19.5% | 23.7% | 26.9% | 29.9% | 35.1% |
| Romania | 1 051 | 1 371 | 1 660 | 1 897 | 2 496 | 19.7% | 24.1% | 27.0% | 30.7% | 36.2% |
| Slovakia | 289 | 423 | 536 | 660 | 883 | 17.7% | 23.0% | 26.9% | 32.0% | 41.4% |
| Slovenia | 75.9 | 131 | 180 | 227 | 317 | 14.1% | 21.4% | 26.7% | 32.0% | 44.1% |
| Spain | 2 063 | 2 547 | 2 904 | 3 358 | 4 151 | 21.3% | 24.9% | 26.9% | 29.4% | 34.7% |
| Sweden | 599 | 812 | 960 | 1 138 | 1 428 | 19.7% | 24.3% | 28.3% | 32.9% | 38.7% |

Table S66: **Country-level averted hospitalisations under 1 year of age – la-mAbs with catch-up, 25% coverage.** For each country, we report the 5th, 25th, 50th (median), 75th, and 95th percentiles of both absolute averted hospitalisations and the corresponding percentage, computed across the 300 retained simulations.

| | **Absolute averted** | | | | | **Percentage averted** | | | | |
|---|---|---|---|---|---|---|---|---|---|---|
| **Country** | **Q05** | **Q25** | **Median** | **Q75** | **Q95** | **Q05** | **Q25** | **Median** | **Q75** | **Q95** |
| Austria | 50.8 | 260 | 343 | 430 | 697 | 2.8% | 10.5% | 13.7% | 16.6% | 25.1% |
| Belgium | 294 | 407 | 500 | 610 | 815 | 8.0% | 11.5% | 13.2% | 15.4% | 18.6% |
| Bulgaria | 43.0 | 222 | 308 | 417 | 621 | 2.3% | 10.1% | 13.8% | 17.4% | 26.0% |
| Croatia | -92.0 | 80.8 | 178 | 272 | 495 | 0.0% | 6.8% | 14.4% | 21.1% | 32.4% |
| Cyprus | -40.0 | 16.0 | 43.5 | 68.2 | 160 | 0.0% | 4.9% | 12.8% | 20.8% | 38.7% |
| Czechia | 187 | 386 | 484 | 621 | 961 | 5.0% | 10.7% | 13.2% | 16.3% | 23.1% |
| Denmark | 106 | 193 | 254 | 342 | 513 | 5.2% | 10.1% | 13.0% | 16.2% | 22.8% |
| Estonia | -23.1 | 23.0 | 50.0 | 86.0 | 159 | 0.0% | 7.1% | 12.6% | 18.9% | 32.7% |
| Finland | 39.8 | 148 | 220 | 313 | 470 | 2.6% | 9.3% | 13.4% | 17.7% | 23.9% |
| France | 2 912 | 3 638 | 4 250 | 5 023 | 6 410 | 10.0% | 12.1% | 13.6% | 15.5% | 19.3% |
| Germany | 2 400 | 3 076 | 3 588 | 4 279 | 5 111 | 10.2% | 12.3% | 13.6% | 15.4% | 18.4% |
| Greece | 141 | 294 | 394 | 511 | 872 | 4.7% | 10.4% | 13.1% | 15.9% | 23.9% |
| Hungary | 181 | 365 | 478 | 620 | 895 | 4.9% | 10.7% | 13.8% | 17.2% | 23.2% |
| Ireland | 40.0 | 205 | 304 | 429 | 662 | 2.1% | 9.7% | 14.2% | 19.1% | 28.6% |
| Italy | 1 322 | 1 783 | 2 077 | 2 519 | 3 064 | 9.4% | 12.1% | 13.7% | 15.7% | 18.6% |
| Latvia | -65.3 | 36.8 | 76.0 | 119 | 219 | 0.0% | 7.1% | 13.9% | 20.9% | 37.8% |
| Lithuania | -0.1 | 59.5 | 103 | 156 | 244 | 0.0% | 7.6% | 13.1% | 17.5% | 28.6% |
| Luxembourg | -23.0 | 6.0 | 24.5 | 44.0 | 86.0 | 0.0% | 3.9% | 13.8% | 23.2% | 38.1% |
| Malta | -12.1 | 4.0 | 13.5 | 25.0 | 47.1 | 0.0% | 4.0% | 12.4% | 20.9% | 38.6% |
| Netherlands | 262 | 405 | 498 | 633 | 892 | 9.0% | 12.2% | 14.5% | 17.1% | 22.3% |
| Norway | 69.8 | 146 | 194 | 252 | 384 | 4.6% | 9.8% | 12.6% | 15.6% | 21.2% |
| Poland | 548 | 932 | 1 207 | 1 515 | 2 400 | 7.8% | 11.1% | 12.9% | 15.7% | 22.1% |
| Portugal | 151 | 251 | 332 | 427 | 623 | 6.4% | 10.2% | 12.9% | 16.3% | 22.2% |
| Romania | 312 | 610 | 802 | 993 | 1 469 | 5.3% | 10.4% | 13.2% | 16.1% | 22.3% |
| Slovakia | 60.5 | 170 | 257 | 364 | 586 | 3.4% | 9.0% | 13.1% | 17.1% | 24.7% |
| Slovenia | -37.2 | 52.8 | 86.0 | 127 | 238 | 0.0% | 8.0% | 13.1% | 18.6% | 35.2% |
| Spain | 846 | 1 210 | 1 406 | 1 688 | 2 225 | 8.8% | 11.6% | 13.2% | 15.0% | 18.2% |
| Sweden | 133 | 340 | 454 | 594 | 997 | 4.1% | 9.8% | 13.5% | 17.7% | 26.1% |

#### S5.3 Maternal vaccination

Table S67: **Country-level averted hospitalisations under 1 year of age – Maternal vaccination, 95% coverage.** For each country, we report the 5th, 25th, 50th (median), 75th, and 95th percentiles of both absolute averted hospitalisations and the corresponding percentage, computed across the 300 retained simulations.

| | Absolute averted | | | | | Percentage averted | | | | |
|---|---|---|---|---|---|---|---|---|---|---|
| **Country** | **Q05** | **Q25** | **Median** | **Q75** | **Q95** | **Q05** | **Q25** | **Median** | **Q75** | **Q95** |
| Austria | 269 | 464 | 578 | 721 | 1 017 | 13.2% | 18.9% | 23.5% | 27.8% | 35.5% |
| Belgium | 453 | 667 | 856 | 1 023 | 1 379 | 13.6% | 18.1% | 21.8% | 25.7% | 31.5% |
| Bulgaria | 177 | 378 | 518 | 673 | 873 | 8.6% | 16.8% | 22.6% | 27.8% | 35.0% |
| Croatia | 57.8 | 219 | 317 | 440 | 666 | 6.0% | 18.1% | 24.6% | 32.3% | 44.0% |
| Cyprus | -40.1 | 43.8 | 77.5 | 120 | 220 | 0.0% | 14.2% | 23.1% | 33.3% | 47.8% |
| Czechia | 416 | 610 | 784 | 966 | 1 340 | 12.4% | 17.5% | 21.1% | 25.5% | 31.2% |
| Denmark | 208 | 327 | 426 | 537 | 704 | 10.8% | 16.7% | 21.3% | 24.8% | 31.7% |
| Estonia | -4.2 | 48.8 | 80.0 | 125 | 194 | 0.0% | 13.5% | 20.6% | 28.2% | 38.9% |
| Finland | 160 | 297 | 382 | 492 | 751 | 10.6% | 18.1% | 23.0% | 27.5% | 37.6% |
| France | 4 473 | 6 183 | 7 288 | 8 487 | 10 535 | 15.7% | 20.1% | 23.2% | 26.7% | 32.8% |
| Germany | 3 496 | 4 862 | 5 930 | 7 068 | 8 971 | 14.1% | 19.2% | 22.6% | 25.6% | 31.2% |
| Greece | 340 | 526 | 660 | 849 | 1 180 | 12.2% | 17.7% | 21.6% | 26.2% | 35.4% |
| Hungary | 441 | 636 | 788 | 985 | 1 341 | 13.6% | 18.7% | 22.4% | 27.3% | 34.9% |
| Ireland | 200 | 376 | 504 | 618 | 871 | 10.6% | 18.2% | 23.0% | 27.4% | 35.7% |
| Italy | 2 178 | 2 934 | 3 550 | 4 222 | 5 214 | 15.4% | 20.0% | 23.1% | 26.9% | 31.1% |
| Latvia | -27.7 | 72.0 | 114 | 164 | 275 | 0.0% | 13.9% | 20.9% | 28.6% | 46.5% |
| Lithuania | 45.9 | 116 | 167 | 236 | 362 | 6.7% | 15.1% | 20.8% | 26.9% | 39.8% |
| Luxembourg | -12.1 | 21.0 | 40.5 | 59.0 | 99.1 | 0.0% | 12.5% | 22.1% | 29.5% | 46.2% |
| Malta | -1.1 | 13.0 | 25.0 | 39.0 | 69.0 | 0.0% | 13.4% | 24.0% | 33.7% | 49.5% |
| Netherlands | 452 | 698 | 882 | 1 064 | 1 354 | 15.6% | 20.6% | 25.6% | 28.8% | 34.7% |
| Norway | 163 | 242 | 319 | 399 | 557 | 11.4% | 16.3% | 20.0% | 24.8% | 31.0% |
| Poland | 986 | 1 528 | 2 073 | 2 709 | 3 727 | 13.2% | 18.3% | 22.7% | 27.4% | 33.3% |
| Portugal | 312 | 466 | 575 | 712 | 981 | 13.6% | 18.6% | 22.3% | 26.5% | 33.1% |
| Romania | 668 | 1 029 | 1 318 | 1 589 | 2 027 | 11.6% | 17.9% | 21.7% | 25.7% | 31.2% |
| Slovakia | 168 | 311 | 427 | 563 | 779 | 10.9% | 16.5% | 21.5% | 27.1% | 35.1% |
| Slovenia | 20.9 | 99.8 | 146 | 205 | 297 | 4.4% | 15.4% | 22.3% | 28.7% | 41.3% |
| Spain | 1 481 | 2 011 | 2 400 | 2 951 | 3 794 | 14.4% | 19.6% | 22.4% | 25.9% | 31.4% |
| Sweden | 358 | 550 | 730 | 928 | 1 286 | 11.1% | 16.9% | 21.7% | 26.9% | 33.6% |

Table S68: **Country-level averted hospitalisations under 1 year of age – Maternal vaccination, 75% coverage.** For each country, we report the 5th, 25th, 50th (median), 75th, and 95th percentiles of both absolute averted hospitalisations and the corresponding percentage, computed across the 300 retained simulations.

| | **Absolute averted** | | | | | **Percentage averted** | | | | |
|---|---|---|---|---|---|---|---|---|---|---|
| **Country** | **Q05** | **Q25** | **Median** | **Q75** | **Q95** | **Q05** | **Q25** | **Median** | **Q75** | **Q95** |
| Austria | 200 | 351 | 454 | 578 | 855 | 9.0% | 14.8% | 18.4% | 22.6% | 31.2% |
| Belgium | 307 | 515 | 653 | 802 | 1 121 | 9.4% | 14.0% | 17.1% | 20.7% | 26.0% |
| Bulgaria | 121 | 286 | 412 | 549 | 746 | 5.4% | 13.2% | 18.0% | 22.1% | 30.8% |
| Croatia | -9.7 | 140 | 243 | 343 | 553 | 0.0% | 11.5% | 18.6% | 26.3% | 38.5% |
| Cyprus | -30.0 | 30.8 | 63.0 | 100 | 178 | 0.0% | 10.4% | 19.0% | 27.2% | 48.9% |
| Czechia | 225 | 438 | 602 | 772 | 1 129 | 6.2% | 12.8% | 16.2% | 19.7% | 27.0% |
| Denmark | 137 | 241 | 330 | 445 | 651 | 7.6% | 12.1% | 16.6% | 20.5% | 29.0% |
| Estonia | -15.1 | 33.0 | 64.5 | 94.0 | 188 | 0.0% | 9.1% | 16.6% | 23.1% | 35.5% |
| Finland | 84.7 | 207 | 304 | 417 | 603 | 5.8% | 12.9% | 17.6% | 23.1% | 31.0% |
| France | 3 479 | 4 667 | 5 764 | 6 825 | 8 602 | 11.4% | 15.9% | 18.5% | 21.2% | 26.0% |
| Germany | 2 756 | 3 796 | 4 648 | 5 582 | 6 924 | 11.6% | 14.7% | 18.0% | 20.3% | 24.2% |
| Greece | 256 | 396 | 510 | 659 | 865 | 9.2% | 13.5% | 16.6% | 20.4% | 26.7% |
| Hungary | 302 | 489 | 608 | 767 | 1 086 | 8.5% | 14.7% | 17.7% | 21.8% | 28.1% |
| Ireland | 150 | 273 | 378 | 500 | 733 | 7.3% | 13.3% | 18.0% | 22.7% | 30.0% |
| Italy | 1 602 | 2 273 | 2 782 | 3 373 | 4 138 | 11.4% | 15.6% | 18.4% | 20.9% | 24.7% |
| Latvia | -58.2 | 50.0 | 91.0 | 139 | 264 | 0.0% | 10.0% | 16.8% | 23.5% | 39.2% |
| Lithuania | 16.9 | 82.0 | 128 | 183 | 280 | 2.4% | 10.7% | 16.7% | 21.6% | 31.2% |
| Luxembourg | -24.0 | 12.0 | 29.0 | 49.5 | 92.0 | 0.0% | 7.2% | 16.4% | 26.2% | 40.7% |
| Malta | -7.0 | 9.0 | 19.0 | 33.2 | 57.0 | 0.0% | 9.2% | 19.4% | 27.9% | 40.3% |
| Netherlands | 361 | 557 | 704 | 828 | 1 115 | 11.8% | 16.4% | 19.8% | 23.3% | 28.5% |
| Norway | 94.8 | 188 | 248 | 306 | 480 | 6.0% | 12.2% | 15.7% | 19.3% | 26.7% |
| Poland | 720 | 1 161 | 1 608 | 2 117 | 2 901 | 9.8% | 13.9% | 17.6% | 21.3% | 27.7% |
| Portugal | 233 | 354 | 463 | 583 | 843 | 10.2% | 14.3% | 17.7% | 22.1% | 27.9% |
| Romania | 452 | 799 | 1 038 | 1 281 | 1 756 | 8.6% | 13.8% | 16.8% | 20.8% | 27.5% |
| Slovakia | 97.0 | 238 | 349 | 477 | 689 | 5.0% | 13.0% | 17.8% | 22.8% | 30.2% |
| Slovenia | 8.9 | 73.0 | 120 | 171 | 256 | 1.4% | 11.6% | 18.2% | 24.3% | 37.2% |
| Spain | 1 151 | 1 533 | 1 868 | 2 302 | 3 031 | 12.0% | 14.9% | 17.6% | 20.6% | 25.2% |
| Sweden | 225 | 422 | 583 | 756 | 1 064 | 6.6% | 12.9% | 17.3% | 21.6% | 29.6% |

Table S69: **Country-level averted hospitalisations under 1 year of age – Maternal vaccination, 50% coverage.** For each country, we report the 5th, 25th, 50th (median), 75th, and 95th percentiles of both absolute averted hospitalisations and the corresponding percentage, computed across the 300 retained simulations.

| | **Absolute averted** | | | | | **Percentage averted** | | | | |
|---|---|---|---|---|---|---|---|---|---|---|
| **Country** | **Q05** | **Q25** | **Median** | **Q75** | **Q95** | **Q05** | **Q25** | **Median** | **Q75** | **Q95** |
| Austria | 66.3 | 213 | 298 | 414 | 623 | 2.9% | 8.7% | 12.2% | 15.7% | 23.0% |
| Belgium | 174 | 324 | 427 | 546 | 766 | 4.8% | 8.9% | 11.6% | 13.8% | 18.0% |
| Bulgaria | 3.9 | 155 | 280 | 392 | 621 | 0.2% | 7.5% | 11.7% | 16.1% | 25.9% |
| Croatia | -97.3 | 72.8 | 164 | 270 | 434 | 0.0% | 5.9% | 12.7% | 19.3% | 30.7% |
| Cyprus | -64.0 | 4.8 | 40.5 | 77.2 | 166 | 0.0% | 1.5% | 12.3% | 22.1% | 42.9% |
| Czechia | 116 | 299 | 420 | 550 | 887 | 3.3% | 8.4% | 11.2% | 14.6% | 22.6% |
| Denmark | 37.8 | 143 | 220 | 289 | 429 | 2.0% | 7.5% | 10.7% | 13.9% | 20.5% |
| Estonia | -35.1 | 13.8 | 44.0 | 75.0 | 160 | 0.0% | 4.3% | 10.7% | 18.1% | 34.6% |
| Finland | -4.2 | 112 | 196 | 273 | 479 | 0.0% | 7.2% | 11.6% | 16.0% | 25.8% |
| France | 2 105 | 3 113 | 3 847 | 4 563 | 5 701 | 7.1% | 10.2% | 12.3% | 14.5% | 18.2% |
| Germany | 1 647 | 2 480 | 3 151 | 3 901 | 5 040 | 7.0% | 9.9% | 12.1% | 14.1% | 17.1% |
| Greece | 96.0 | 242 | 334 | 460 | 757 | 3.2% | 8.1% | 11.2% | 14.6% | 21.3% |
| Hungary | 118 | 303 | 432 | 589 | 887 | 3.6% | 9.0% | 12.3% | 16.1% | 23.5% |
| Ireland | -24.1 | 170 | 270 | 395 | 630 | 0.0% | 8.1% | 12.9% | 17.2% | 26.3% |
| Italy | 1 002 | 1 549 | 1 884 | 2 324 | 3 103 | 7.0% | 10.4% | 12.3% | 14.7% | 18.5% |
| Latvia | -51.2 | 28.0 | 70.5 | 102 | 191 | 0.0% | 5.0% | 12.2% | 18.4% | 33.9% |
| Lithuania | -40.1 | 37.8 | 86.5 | 140 | 252 | 0.0% | 4.9% | 10.5% | 16.5% | 27.8% |
| Luxembourg | -36.3 | 2.0 | 21.0 | 38.0 | 78.0 | 0.0% | 1.2% | 12.0% | 19.1% | 34.9% |
| Malta | -12.0 | 4.0 | 13.5 | 25.0 | 47.0 | 0.0% | 3.3% | 13.4% | 20.9% | 35.0% |
| Netherlands | 171 | 346 | 452 | 581 | 842 | 5.3% | 10.3% | 13.3% | 15.9% | 21.7% |
| Norway | 40.9 | 111 | 156 | 220 | 348 | 2.8% | 7.6% | 10.3% | 13.7% | 20.6% |
| Poland | 428 | 758 | 1 069 | 1 381 | 2 407 | 5.0% | 9.2% | 11.9% | 14.8% | 22.2% |
| Portugal | 112 | 217 | 294 | 414 | 593 | 4.7% | 8.6% | 11.6% | 15.2% | 20.9% |
| Romania | 166 | 494 | 677 | 934 | 1 271 | 2.9% | 8.5% | 11.6% | 14.6% | 19.6% |
| Slovakia | -1.1 | 125 | 224 | 325 | 553 | 0.0% | 7.0% | 11.3% | 15.8% | 23.2% |
| Slovenia | -26.1 | 35.8 | 82.5 | 122 | 212 | 0.0% | 5.9% | 12.4% | 18.5% | 27.9% |
| Spain | 588 | 989 | 1 244 | 1 540 | 2 176 | 6.1% | 9.4% | 11.6% | 13.7% | 18.0% |
| Sweden | -22.1 | 240 | 370 | 556 | 803 | 0.0% | 7.1% | 11.1% | 15.8% | 22.9% |

Table S70: **Country-level averted hospitalisations under 1 year of age – Maternal vaccination, 25% coverage.** For each country, we report the 5th, 25th, 50th (median), 75th, and 95th percentiles of both absolute averted hospitalisations and the corresponding percentage, computed across the 300 retained simulations.

| | Absolute averted | | | | | Percentage averted | | | | |
|---|---|---|---|---|---|---|---|---|---|---|
| **Country** | **Q05** | **Q25** | **Median** | **Q75** | **Q95** | **Q05** | **Q25** | **Median** | **Q75** | **Q95** |
| Austria | -131 | 76.0 | 160 | 248 | 469 | 0.0% | 3.4% | 6.6% | 9.6% | 17.1% |
| Belgium | -34.0 | 128 | 213 | 307 | 483 | 0.0% | 3.7% | 5.8% | 7.9% | 11.4% |
| Bulgaria | -174 | 17.2 | 124 | 232 | 402 | 0.0% | 0.7% | 5.3% | 9.4% | 16.5% |
| Croatia | -241 | -32.5 | 74.5 | 187 | 357 | 0.0% | 0.0% | 5.9% | 13.9% | 24.9% |
| Cyprus | -69.3 | -11.2 | 21.5 | 62.2 | 134 | 0.0% | 0.0% | 6.9% | 17.2% | 38.6% |
| Czechia | -110 | 93.0 | 204 | 312 | 674 | 0.0% | 2.8% | 5.6% | 8.2% | 16.8% |
| Denmark | -113 | 44.5 | 103 | 186 | 367 | 0.0% | 2.1% | 5.3% | 8.9% | 15.6% |
| Estonia | -77.5 | -5.0 | 19.0 | 46.0 | 143 | 0.0% | 0.0% | 4.8% | 12.1% | 32.4% |
| Finland | -113 | 9.0 | 102 | 174 | 372 | 0.0% | 0.6% | 6.0% | 10.0% | 18.8% |
| France | 543 | 1 255 | 1 898 | 2 505 | 3 840 | 1.8% | 4.2% | 5.9% | 7.9% | 11.7% |
| Germany | 533 | 1 118 | 1 582 | 2 128 | 2 860 | 2.2% | 4.6% | 5.9% | 7.7% | 10.5% |
| Greece | -114 | 82.0 | 165 | 262 | 519 | 0.0% | 2.5% | 5.4% | 8.3% | 15.9% |
| Hungary | -110 | 87.8 | 212 | 349 | 608 | 0.0% | 2.4% | 6.4% | 9.9% | 15.9% |
| Ireland | -199 | 31.0 | 142 | 258 | 470 | 0.0% | 1.4% | 6.4% | 10.9% | 20.7% |
| Italy | 306 | 654 | 896 | 1 211 | 1 717 | 2.2% | 4.2% | 6.0% | 7.6% | 10.2% |
| Latvia | -130 | -2.2 | 34.0 | 68.0 | 170 | 0.0% | 0.0% | 6.1% | 13.0% | 28.2% |
| Lithuania | -81.1 | 3.0 | 44.0 | 90.2 | 211 | 0.0% | 0.4% | 6.0% | 10.2% | 22.0% |
| Luxembourg | -39.0 | -8.0 | 10.5 | 25.0 | 75.0 | 0.0% | 0.0% | 6.1% | 14.0% | 31.3% |
| Malta | -23.1 | -4.0 | 7.0 | 17.0 | 37.0 | 0.0% | 0.0% | 6.2% | 15.5% | 29.4% |
| Netherlands | -21.1 | 130 | 222 | 316 | 573 | 0.0% | 3.7% | 6.5% | 9.1% | 13.8% |
| Norway | -56.2 | 26.0 | 77.5 | 125 | 259 | 0.0% | 1.9% | 5.0% | 8.1% | 14.5% |
| Poland | 76.0 | 316 | 526 | 766 | 1 526 | 0.9% | 3.8% | 6.0% | 8.0% | 14.3% |
| Portugal | -25.0 | 85.8 | 145 | 226 | 396 | 0.0% | 3.3% | 6.0% | 8.9% | 14.2% |
| Romania | -129 | 166 | 358 | 530 | 1 000 | 0.0% | 2.8% | 5.8% | 8.5% | 14.4% |
| Slovakia | -84.2 | 25.8 | 110 | 195 | 387 | 0.0% | 1.2% | 5.6% | 9.6% | 18.4% |
| Slovenia | -79.1 | -6.2 | 35.0 | 73.0 | 161 | 0.0% | 0.0% | 5.1% | 10.3% | 21.9% |
| Spain | 111 | 428 | 620 | 846 | 1 213 | 0.9% | 4.2% | 6.1% | 7.6% | 10.8% |
| Sweden | -172 | 64.2 | 197 | 344 | 633 | 0.0% | 2.0% | 6.1% | 9.9% | 17.1% |

#### S5.4 Maternal vaccination with seasonal catch-up

Table S71: **Country-level averted hospitalisations under 1 year of age – Maternal vaccination with catch-up,** 95% **coverage.** For each country, we report the 5th, 25th, 50th (median), 75th, and 95th percentiles of both absolute averted hospitalisations and the corresponding percentage, computed across the 300 retained simulations.

| | Absolute averted | | | | | Percentage averted | | | | |
|---|---|---|---|---|---|---|---|---|---|---|
| **Country** | **Q05** | **Q25** | **Median** | **Q75** | **Q95** | **Q05** | **Q25** | **Median** | **Q75** | **Q95** |
| Austria | 751 | 949 | 1 103 | 1 321 | 1 666 | 34.0% | 40.7% | 44.9% | 51.1% | 58.0% |
| Belgium | 1 075 | 1 360 | 1 593 | 1 882 | 2 397 | 32.1% | 37.7% | 42.0% | 46.0% | 53.7% |
| Bulgaria | 673 | 877 | 1 026 | 1 199 | 1 510 | 32.2% | 39.3% | 44.8% | 50.3% | 59.0% |
| Croatia | 310 | 474 | 599 | 716 | 929 | 30.8% | 39.5% | 46.5% | 52.2% | 61.8% |
| Cyprus | 66.0 | 109 | 143 | 181 | 271 | 25.4% | 35.2% | 43.0% | 51.7% | 65.2% |
| Czechia | 1 124 | 1 426 | 1 646 | 1 917 | 2 300 | 34.4% | 40.2% | 44.9% | 48.7% | 55.6% |
| Denmark | 598 | 757 | 901 | 1 057 | 1 303 | 32.0% | 39.6% | 44.0% | 48.9% | 57.6% |
| Estonia | 74.8 | 123 | 158 | 211 | 317 | 24.6% | 33.6% | 40.0% | 48.7% | 62.6% |
| Finland | 473 | 627 | 748 | 893 | 1 152 | 31.9% | 40.0% | 44.1% | 50.4% | 59.1% |
| France | 10 368 | 12 782 | 14 485 | 16 091 | 18 456 | 36.6% | 42.3% | 46.0% | 50.5% | 55.1% |
| Germany | 8 187 | 10 109 | 11 896 | 13 517 | 15 826 | 35.6% | 40.5% | 45.4% | 49.2% | 54.4% |
| Greece | 874 | 1 126 | 1 291 | 1 521 | 1 885 | 32.4% | 38.1% | 42.2% | 47.2% | 54.0% |
| Hungary | 1 128 | 1 396 | 1 608 | 1 854 | 2 264 | 35.2% | 41.4% | 46.0% | 51.1% | 57.5% |
| Ireland | 695 | 862 | 1 010 | 1 182 | 1 450 | 34.4% | 42.1% | 46.7% | 51.5% | 59.1% |
| Italy | 4 669 | 5 911 | 6 755 | 7 711 | 9 127 | 35.5% | 40.5% | 43.9% | 48.4% | 53.0% |
| Latvia | 106 | 187 | 234 | 298 | 424 | 25.3% | 36.8% | 43.1% | 51.5% | 65.2% |
| Lithuania | 203 | 281 | 343 | 423 | 592 | 28.9% | 37.2% | 43.5% | 49.6% | 60.5% |
| Luxembourg | 27.8 | 59.0 | 78.0 | 106 | 157 | 19.0% | 34.8% | 42.6% | 52.5% | 69.1% |
| Malta | 10.9 | 29.8 | 40.0 | 57.0 | 84.0 | 14.0% | 30.8% | 38.8% | 48.5% | 60.5% |
| Netherlands | 1 081 | 1 425 | 1 675 | 1 934 | 2 428 | 35.8% | 43.3% | 47.8% | 52.3% | 59.2% |
| Norway | 436 | 572 | 672 | 791 | 1 019 | 32.5% | 38.0% | 42.8% | 47.8% | 56.1% |
| Poland | 2 211 | 3 277 | 4 069 | 4 950 | 6 237 | 33.4% | 38.4% | 43.3% | 50.0% | 57.4% |
| Portugal | 706 | 907 | 1 090 | 1 324 | 1 710 | 31.7% | 37.0% | 42.7% | 47.6% | 55.8% |
| Romania | 1 810 | 2 344 | 2 734 | 3 131 | 3 678 | 34.4% | 40.2% | 45.2% | 50.2% | 56.8% |
| Slovakia | 541 | 737 | 884 | 1 055 | 1 282 | 32.1% | 39.4% | 45.1% | 51.4% | 58.7% |
| Slovenia | 171 | 238 | 287 | 342 | 458 | 30.5% | 37.6% | 43.4% | 48.8% | 59.0% |
| Spain | 3 239 | 4 024 | 4 634 | 5 469 | 6 602 | 33.0% | 38.9% | 43.4% | 47.6% | 54.3% |
| Sweden | 1 140 | 1 384 | 1 620 | 1 841 | 2 265 | 36.3% | 42.6% | 47.6% | 52.5% | 57.8% |

Table S72: **Country-level averted hospitalisations under 1 year of age – Maternal vaccination with catch-up, 75% coverage.** For each country, we report the 5th, 25th, 50th (median), 75th, and 95th percentiles of both absolute averted hospitalisations and the corresponding percentage, computed across the 300 retained simulations.

| | **Absolute averted** | | | | | **Percentage averted** | | | | |
|---|---|---|---|---|---|---|---|---|---|---|
| **Country** | **Q05** | **Q25** | **Median** | **Q75** | **Q95** | **Q05** | **Q25** | **Median** | **Q75** | **Q95** |
| Austria | 546 | 738 | 869 | 1 042 | 1 405 | 25.1% | 31.3% | 35.7% | 40.4% | 49.3% |
| Belgium | 818 | 1 040 | 1 246 | 1 489 | 1 910 | 24.6% | 29.3% | 33.0% | 37.1% | 42.2% |
| Bulgaria | 519 | 669 | 816 | 990 | 1 268 | 24.5% | 31.3% | 35.9% | 41.3% | 49.3% |
| Croatia | 224 | 367 | 457 | 580 | 813 | 18.3% | 29.8% | 36.3% | 42.2% | 55.5% |
| Cyprus | 47.0 | 84.0 | 116 | 154 | 235 | 16.9% | 26.9% | 35.6% | 43.0% | 58.2% |
| Czechia | 816 | 1 103 | 1 299 | 1 518 | 1 870 | 25.7% | 30.6% | 34.9% | 39.4% | 45.5% |
| Denmark | 441 | 584 | 718 | 835 | 1 027 | 25.3% | 30.8% | 34.8% | 38.8% | 45.1% |
| Estonia | 42.0 | 92.0 | 128 | 177 | 259 | 13.8% | 25.8% | 33.0% | 39.7% | 54.9% |
| Finland | 347 | 480 | 580 | 716 | 952 | 23.4% | 30.8% | 35.0% | 39.9% | 48.6% |
| France | 8 134 | 10 081 | 11 432 | 12 838 | 14 860 | 28.5% | 33.0% | 36.2% | 39.9% | 44.0% |
| Germany | 6 373 | 7 925 | 9 468 | 10 739 | 12 639 | 27.7% | 32.2% | 35.8% | 39.3% | 43.2% |
| Greece | 666 | 875 | 1 020 | 1 216 | 1 547 | 24.6% | 29.9% | 33.5% | 38.0% | 44.5% |
| Hungary | 872 | 1 078 | 1 272 | 1 498 | 1 828 | 26.5% | 33.0% | 36.3% | 40.7% | 47.6% |
| Ireland | 512 | 674 | 801 | 950 | 1 245 | 25.5% | 32.9% | 37.2% | 41.8% | 49.1% |
| Italy | 3 739 | 4 653 | 5 318 | 6 075 | 7 119 | 27.7% | 31.8% | 34.5% | 38.2% | 42.0% |
| Latvia | 56.8 | 142 | 186 | 245 | 370 | 13.6% | 28.2% | 34.1% | 41.9% | 57.5% |
| Lithuania | 141 | 221 | 274 | 345 | 465 | 21.0% | 29.0% | 34.0% | 39.4% | 49.9% |
| Luxembourg | 19.9 | 43.0 | 62.0 | 84.0 | 138 | 11.2% | 25.5% | 34.0% | 42.8% | 61.0% |
| Malta | 3.0 | 19.8 | 31.0 | 46.0 | 72.1 | 4.8% | 20.4% | 30.0% | 38.7% | 52.5% |
| Netherlands | 840 | 1 118 | 1 316 | 1 550 | 1 926 | 27.7% | 33.5% | 37.8% | 41.8% | 47.3% |
| Norway | 337 | 449 | 522 | 625 | 791 | 24.6% | 30.0% | 33.6% | 37.6% | 46.5% |
| Poland | 1 675 | 2 548 | 3 201 | 3 996 | 5 096 | 25.2% | 29.9% | 34.3% | 39.5% | 47.8% |
| Portugal | 561 | 710 | 864 | 1 040 | 1 372 | 24.8% | 29.2% | 33.3% | 37.9% | 44.9% |
| Romania | 1 448 | 1 834 | 2 174 | 2 523 | 3 024 | 26.6% | 31.6% | 35.9% | 39.9% | 45.9% |
| Slovakia | 394 | 574 | 704 | 848 | 1 080 | 24.4% | 30.8% | 36.2% | 41.0% | 48.0% |
| Slovenia | 126 | 185 | 240 | 286 | 387 | 21.8% | 29.7% | 34.8% | 41.1% | 51.4% |
| Spain | 2 469 | 3 169 | 3 635 | 4 288 | 5 234 | 25.7% | 30.6% | 33.8% | 37.3% | 43.0% |
| Sweden | 848 | 1 082 | 1 276 | 1 478 | 1 852 | 27.9% | 32.7% | 37.7% | 42.3% | 49.5% |

Table S73: **Country-level averted hospitalisations under 1 year of age – Maternal vaccination with catch-up,** 50% **coverage.** For each country, we report the 5th, 25th, 50th (median), 75th, and 95th percentiles of both absolute averted hospitalisations and the corresponding percentage, computed across the 300 retained simulations.

| | **Absolute averted** | | | | | **Percentage averted** | | | | |
|---|---|---|---|---|---|---|---|---|---|---|
| **Country** | **Q05** | **Q25** | **Median** | **Q75** | **Q95** | **Q05** | **Q25** | **Median** | **Q75** | **Q95** |
| Austria | 348 | 491 | 587 | 696 | 993 | 14.9% | 20.7% | 23.7% | 27.8% | 34.8% |
| Belgium | 509 | 698 | 818 | 996 | 1 304 | 15.7% | 19.4% | 21.9% | 24.5% | 29.1% |
| Bulgaria | 279 | 420 | 525 | 680 | 920 | 13.4% | 19.4% | 22.9% | 28.0% | 36.7% |
| Croatia | 67.5 | 209 | 308 | 436 | 613 | 5.4% | 17.9% | 24.3% | 31.1% | 43.1% |
| Cyprus | -8.5 | 45.0 | 74.5 | 107 | 198 | 0.0% | 14.3% | 22.3% | 29.9% | 51.6% |
| Czechia | 525 | 728 | 850 | 1 019 | 1 403 | 15.8% | 20.1% | 22.8% | 26.6% | 32.9% |
| Denmark | 255 | 367 | 479 | 590 | 776 | 14.6% | 19.0% | 23.3% | 27.8% | 34.5% |
| Estonia | 1.0 | 52.8 | 87.0 | 123 | 208 | 0.3% | 14.5% | 21.9% | 29.7% | 41.9% |
| Finland | 198 | 314 | 398 | 498 | 713 | 14.0% | 19.9% | 23.5% | 27.5% | 39.0% |
| France | 5 241 | 6 442 | 7 608 | 8 586 | 10 289 | 18.1% | 21.6% | 24.1% | 26.7% | 30.6% |
| Germany | 4 159 | 5 226 | 6 196 | 7 314 | 8 849 | 18.2% | 21.1% | 23.5% | 26.2% | 30.1% |
| Greece | 397 | 551 | 676 | 823 | 1 142 | 14.0% | 19.2% | 22.2% | 25.9% | 33.3% |
| Hungary | 498 | 691 | 846 | 1 006 | 1 305 | 15.7% | 20.8% | 24.3% | 27.4% | 34.5% |
| Ireland | 295 | 426 | 550 | 654 | 941 | 14.3% | 20.8% | 25.8% | 29.2% | 38.4% |
| Italy | 2 487 | 3 094 | 3 544 | 4 086 | 4 912 | 17.9% | 21.0% | 23.1% | 25.7% | 29.1% |
| Latvia | -9.2 | 84.8 | 126 | 183 | 303 | 0.0% | 16.7% | 22.8% | 30.6% | 46.1% |
| Lithuania | 47.0 | 121 | 174 | 229 | 360 | 6.7% | 16.2% | 22.2% | 27.0% | 39.3% |
| Luxembourg | -11.2 | 21.8 | 43.0 | 62.0 | 110 | 0.0% | 13.9% | 23.3% | 32.1% | 51.0% |
| Malta | -8.1 | 10.0 | 21.0 | 33.0 | 55.1 | 0.0% | 10.2% | 19.8% | 27.7% | 41.2% |
| Netherlands | 543 | 722 | 889 | 1 047 | 1 400 | 17.8% | 22.0% | 25.2% | 28.3% | 34.3% |
| Norway | 193 | 288 | 350 | 423 | 598 | 13.5% | 19.0% | 22.3% | 25.6% | 33.1% |
| Poland | 1 123 | 1 652 | 2 102 | 2 686 | 3 834 | 16.1% | 19.5% | 22.6% | 26.5% | 34.5% |
| Portugal | 339 | 458 | 566 | 708 | 950 | 14.7% | 18.8% | 21.9% | 26.0% | 31.8% |
| Romania | 865 | 1 170 | 1 439 | 1 712 | 2 244 | 14.9% | 20.5% | 23.6% | 27.0% | 32.7% |
| Slovakia | 216 | 357 | 456 | 577 | 812 | 12.6% | 19.4% | 23.4% | 28.0% | 36.7% |
| Slovenia | 30.8 | 106 | 158 | 217 | 303 | 5.4% | 16.8% | 23.7% | 29.4% | 43.2% |
| Spain | 1 670 | 2 118 | 2 446 | 2 888 | 3 621 | 17.1% | 20.5% | 22.5% | 25.1% | 30.9% |
| Sweden | 485 | 682 | 852 | 1 018 | 1 352 | 15.9% | 20.8% | 24.4% | 28.9% | 37.2% |

Table S74: **Country-level averted hospitalisations under 1 year of age – Maternal vaccination with catch-up,** 25% **coverage.** For each country, we report the 5th, 25th, 50th (median), 75th, and 95th percentiles of both absolute averted hospitalisations and the corresponding percentage, computed across the 300 retained simulations.

| | **Absolute averted** | | | | | **Percentage averted** | | | | |
|---|---|---|---|---|---|---|---|---|---|---|
| **Country** | **Q05** | **Q25** | **Median** | **Q75** | **Q95** | **Q05** | **Q25** | **Median** | **Q75** | **Q95** |
| Austria | 38.0 | 196 | 288 | 395 | 670 | 2.3% | 8.0% | 12.0% | 15.6% | 24.0% |
| Belgium | 195 | 324 | 418 | 526 | 786 | 5.9% | 9.0% | 11.2% | 13.5% | 18.5% |
| Bulgaria | 44.9 | 171 | 262 | 372 | 609 | 2.0% | 7.7% | 11.4% | 16.0% | 24.0% |
| Croatia | -145 | 57.0 | 142 | 258 | 445 | 0.0% | 4.3% | 11.5% | 18.4% | 30.8% |
| Cyprus | -47.4 | 8.0 | 37.0 | 74.2 | 156 | 0.0% | 2.5% | 10.9% | 21.2% | 36.8% |
| Czechia | 104 | 297 | 428 | 550 | 834 | 3.4% | 8.5% | 11.3% | 14.4% | 20.4% |
| Denmark | 43.8 | 154 | 240 | 324 | 499 | 2.1% | 8.0% | 11.7% | 15.4% | 22.7% |
| Estonia | -40.1 | 15.0 | 39.0 | 77.0 | 153 | 0.0% | 4.1% | 10.2% | 17.4% | 35.8% |
| Finland | -8.9 | 112 | 180 | 269 | 455 | 0.0% | 7.2% | 11.0% | 15.5% | 23.8% |
| France | 2 224 | 3 069 | 3 658 | 4 396 | 5 669 | 7.3% | 9.9% | 11.6% | 14.0% | 17.1% |
| Germany | 1 941 | 2 518 | 3 158 | 3 691 | 4 614 | 8.3% | 10.2% | 11.8% | 13.5% | 16.7% |
| Greece | 94.8 | 240 | 322 | 430 | 733 | 3.6% | 8.2% | 10.7% | 13.7% | 21.5% |
| Hungary | 143 | 306 | 412 | 573 | 870 | 4.3% | 9.0% | 11.8% | 15.6% | 22.5% |
| Ireland | -21.6 | 178 | 300 | 418 | 660 | 0.0% | 8.7% | 13.5% | 18.1% | 29.9% |
| Italy | 1 086 | 1 497 | 1 803 | 2 117 | 2 697 | 7.5% | 10.2% | 11.8% | 13.5% | 16.4% |
| Latvia | -95.2 | 31.0 | 64.0 | 113 | 249 | 0.0% | 6.3% | 11.6% | 19.3% | 39.8% |
| Lithuania | -40.5 | 39.0 | 83.5 | 136 | 267 | 0.0% | 5.4% | 10.7% | 15.7% | 29.2% |
| Luxembourg | -30.0 | 3.0 | 20.0 | 44.0 | 83.4 | 0.0% | 1.9% | 11.2% | 21.2% | 40.6% |
| Malta | -14.1 | 1.0 | 11.0 | 23.0 | 42.0 | 0.0% | 1.0% | 10.8% | 20.1% | 32.2% |
| Netherlands | 169 | 348 | 440 | 543 | 811 | 4.9% | 10.2% | 12.4% | 15.4% | 19.8% |
| Norway | 39.9 | 113 | 168 | 235 | 385 | 2.8% | 7.8% | 10.8% | 14.9% | 21.9% |
| Poland | 399 | 754 | 1 040 | 1 363 | 2 188 | 5.3% | 9.3% | 11.1% | 14.0% | 20.9% |
| Portugal | 104 | 196 | 268 | 374 | 573 | 4.0% | 7.8% | 10.9% | 13.7% | 19.1% |
| Romania | 258 | 521 | 704 | 950 | 1 387 | 4.4% | 8.8% | 11.9% | 15.1% | 21.5% |
| Slovakia | 7.6 | 145 | 246 | 360 | 501 | 0.4% | 7.9% | 12.4% | 16.9% | 23.3% |
| Slovenia | -60.1 | 30.8 | 80.5 | 126 | 233 | 0.0% | 4.6% | 11.8% | 19.6% | 32.3% |
| Spain | 717 | 972 | 1 214 | 1 442 | 1 968 | 6.8% | 9.4% | 11.2% | 13.0% | 16.8% |
| Sweden | 28.9 | 265 | 412 | 554 | 896 | 1.0% | 8.3% | 12.4% | 16.8% | 23.8% |

### S6 Age-specific distribution of averted hospitalisations

This section reports the age-specific share of averted hospitalisations under each intervention scenario and coverage level. For each combination of immunisation strategy and coverage level, the share is computed at the simulation level as the fraction of averted hospitalisations attributable to each age group across the 28 European countries.

Tables are organised by coverage level (decreasing from 95% to 25%) and, within each table, by intervention strategy. For each combination, we report the 5th, 25th, 50th (median), 75th, and 95th percentiles of the simulation-level distribution. Age groups follow the resolution of the calibration targets ($0-2$ m, $3-5$ m, $6-11$ m, $1-4$ y, $5-64$ y, and 65+ y).

Table S75: **Age-specific share of averted hospitalisations** – 95% **coverage.** For each intervention strategy and age group, we report the 5th, 25th, 50th (median), 75th, and 95th percentiles of the simulation-level distribution across the 300 retained simulations, pooled across the 28 European countries.

| Intervention | Age group | Q05 | Q25 | Median | Q75 | Q95 |
|---|---|---|---|---|---|---|
| la-mAbs | 0–2 m | 67.3% | 73.1% | 78.2% | 80.6% | 82.9% |
| | 3–5 m | 14.0% | 15.2% | 16.1% | 17.1% | 18.2% |
| | 6–11 m | 1.4% | 2.2% | 2.8% | 3.4% | 4.3% |
| | 1–4 y | 0.0% | 0.0% | 0.7% | 1.7% | 3.0% |
| | 5–64 y | 0.0% | 0.0% | 0.2% | 0.6% | 1.3% |
| | 65+ y | 0.0% | 0.0% | 1.3% | 5.7% | 10.8% |
| la-mAbs + CU | 0–2 m | 55.5% | 59.4% | 61.7% | 63.8% | 66.3% |
| | 3–5 m | 23.0% | 24.5% | 25.6% | 26.6% | 28.1% |
| | 6–11 m | 6.8% | 7.6% | 8.2% | 8.7% | 9.4% |
| | 1–4 y | 0.0% | 0.7% | 1.3% | 2.0% | 3.0% |
| | 5–64 y | 0.0% | 0.0% | 0.3% | 0.5% | 0.9% |
| | 65+ y | 0.0% | 0.3% | 2.9% | 5.3% | 8.1% |
| MV | 0–2 m | 61.4% | 67.8% | 72.6% | 78.5% | 82.6% |
| | 3–5 m | 12.5% | 13.6% | 14.7% | 15.9% | 17.5% |
| | 6–11 m | 1.6% | 2.5% | 3.1% | 3.7% | 4.5% |
| | 1–4 y | 0.0% | 0.4% | 1.9% | 2.9% | 4.6% |
| | 5–64 y | 0.0% | 0.1% | 0.7% | 1.1% | 1.8% |
| | 65+ y | 0.0% | 1.4% | 7.0% | 10.8% | 16.4% |
| MV + CU | 0–2 m | 51.1% | 54.7% | 57.1% | 59.1% | 62.2% |
| | 3–5 m | 23.0% | 24.5% | 25.9% | 27.4% | 29.3% |
| | 6–11 m | 7.5% | 8.4% | 9.0% | 9.6% | 10.2% |
| | 1–4 y | 0.3% | 1.4% | 2.0% | 2.6% | 3.5% |
| | 5–64 y | 0.0% | 0.2% | 0.5% | 0.8% | 1.2% |
| | 65+ y | 0.0% | 3.4% | 5.5% | 7.8% | 11.1% |

Table S76: **Age-specific share of averted hospitalisations – 75% coverage.** For each intervention strategy and age group, we report the 5th, 25th, 50th (median), 75th, and 95th percentiles of the simulation-level distribution across the 300 retained simulations, pooled across the 28 European countries.

| **Intervention** | **Age group** | **Q05** | **Q25** | **Median** | **Q75** | **Q95** |
|---|---|---|---|---|---|---|
| la-mAbs | 0–2 m | 66.7% | 73.2% | 78.2% | 81.1% | 83.0% |
| | 3–5 m | 13.9% | 15.0% | 16.0% | 17.0% | 18.3% |
| | 6–11 m | 0.9% | 2.1% | 2.8% | 3.3% | 4.3% |
| | 1–4 y | 0.0% | 0.0% | 0.5% | 1.6% | 3.3% |
| | 5–64 y | 0.0% | 0.0% | 0.1% | 0.7% | 1.3% |
| | 65+ y | 0.0% | 0.0% | 1.6% | 5.7% | 10.9% |
| la-mAbs + CU | 0–2 m | 54.6% | 58.7% | 61.8% | 64.0% | 66.1% |
| | 3–5 m | 22.3% | 24.2% | 25.7% | 26.8% | 28.4% |
| | 6–11 m | 6.8% | 7.6% | 8.2% | 8.7% | 9.4% |
| | 1–4 y | 0.0% | 0.5% | 1.4% | 2.1% | 3.3% |
| | 5–64 y | 0.0% | 0.0% | 0.3% | 0.6% | 1.1% |
| | 65+ y | 0.0% | 0.0% | 2.5% | 5.6% | 9.2% |
| MV | 0–2 m | 59.5% | 68.5% | 73.6% | 79.7% | 83.4% |
| | 3–5 m | 12.0% | 13.7% | 14.9% | 16.1% | 17.7% |
| | 6–11 m | 0.9% | 2.3% | 3.0% | 3.8% | 4.7% |
| | 1–4 y | 0.0% | 0.0% | 1.6% | 3.0% | 5.1% |
| | 5–64 y | 0.0% | 0.0% | 0.5% | 1.2% | 2.2% |
| | 65+ y | 0.0% | 0.0% | 5.7% | 10.8% | 17.6% |
| MV + CU | 0–2 m | 50.1% | 54.1% | 56.9% | 59.5% | 62.8% |
| | 3–5 m | 22.6% | 24.2% | 25.8% | 27.5% | 29.4% |
| | 6–11 m | 7.3% | 8.3% | 8.9% | 9.6% | 10.4% |
| | 1–4 y | 0.0% | 1.2% | 2.1% | 2.9% | 4.2% |
| | 5–64 y | 0.0% | 0.2% | 0.5% | 0.9% | 1.4% |
| | 65+ y | 0.0% | 2.3% | 6.0% | 8.7% | 12.6% |

Table S77: **Age-specific share of averted hospitalisations – 50% coverage.** For each intervention strategy and age group, we report the 5th, 25th, 50th (median), 75th, and 95th percentiles of the simulation-level distribution across the 300 retained simulations, pooled across the 28 European countries.

| **Intervention** | **Age group** | **Q05** | **Q25** | **Median** | **Q75** | **Q95** |
|---|---|---|---|---|---|---|
| la-mAbs | 0–2 m | 59.7% | 70.7% | 77.8% | 81.1% | 83.8% |
| | 3–5 m | 12.9% | 14.7% | 15.7% | 16.9% | 18.7% |
| | 6–11 m | 0.1% | 1.8% | 2.8% | 3.8% | 5.0% |
| | 1–4 y | 0.0% | 0.0% | 0.4% | 2.5% | 4.6% |
| | 5–64 y | 0.0% | 0.0% | 0.2% | 0.9% | 2.1% |
| | 65+ y | 0.0% | 0.0% | 1.3% | 7.5% | 16.4% |
| la-mAbs + CU | 0–2 m | 52.2% | 57.7% | 61.3% | 64.1% | 66.4% |
| | 3–5 m | 22.0% | 23.7% | 25.3% | 26.9% | 28.7% |
| | 6–11 m | 6.5% | 7.4% | 8.1% | 8.7% | 9.6% |
| | 1–4 y | 0.0% | 0.3% | 1.5% | 2.4% | 4.0% |
| | 5–64 y | 0.0% | 0.0% | 0.3% | 0.7% | 1.4% |
| | 65+ y | 0.0% | 0.0% | 3.1% | 7.0% | 12.4% |
| MV | 0–2 m | 54.3% | 62.7% | 72.8% | 79.8% | 84.6% |
| | 3–5 m | 11.3% | 13.0% | 14.6% | 16.0% | 18.0% |
| | 6–11 m | 0.0% | 1.9% | 3.2% | 4.2% | 5.4% |
| | 1–4 y | 0.0% | 0.0% | 1.8% | 4.2% | 6.8% |
| | 5–64 y | 0.0% | 0.0% | 0.7% | 1.5% | 2.9% |
| | 65+ y | 0.0% | 0.0% | 6.5% | 14.5% | 21.6% |
| MV + CU | 0–2 m | 48.1% | 53.2% | 56.6% | 60.2% | 64.2% |
| | 3–5 m | 21.6% | 23.8% | 25.8% | 27.6% | 30.2% |
| | 6–11 m | 7.1% | 8.1% | 9.0% | 9.8% | 10.8% |
| | 1–4 y | 0.0% | 0.9% | 2.2% | 3.3% | 4.9% |
| | 5–64 y | 0.0% | 0.0% | 0.5% | 1.1% | 1.7% |
| | 65+ y | 0.0% | 1.1% | 5.1% | 9.3% | 15.0% |

Table S78: **Age-specific share of averted hospitalisations – 25% coverage.** For each intervention strategy and age group, we report the 5th, 25th, 50th (median), 75th, and 95th percentiles of the simulation-level distribution across the 300 retained simulations, pooled across the 28 European countries.

| **Intervention** | **Age group** | **Q05** | **Q25** | **Median** | **Q75** | **Q95** |
|---|---|---|---|---|---|---|
| la-mAbs | 0–2 m | 49.7% | 62.3% | 75.2% | 81.5% | 86.6% |
| | 3–5 m | 10.5% | 13.2% | 15.0% | 17.0% | 19.3% |
| | 6–11 m | 0.0% | 0.8% | 2.9% | 4.5% | 6.2% |
| | 1–4 y | 0.0% | 0.0% | 0.2% | 4.2% | 7.6% |
| | 5–64 y | 0.0% | 0.0% | 0.1% | 1.8% | 3.3% |
| | 65+ y | 0.0% | 0.0% | 0.7% | 15.0% | 25.5% |
| la-mAbs + CU | 0–2 m | 47.9% | 55.9% | 62.0% | 64.7% | 67.7% |
| | 3–5 m | 19.4% | 22.9% | 25.6% | 27.4% | 29.4% |
| | 6–11 m | 5.4% | 6.8% | 7.8% | 8.8% | 10.3% |
| | 1–4 y | 0.0% | 0.0% | 0.9% | 3.2% | 5.6% |
| | 5–64 y | 0.0% | 0.0% | 0.1% | 0.9% | 2.3% |
| | 65+ y | 0.0% | 0.0% | 1.1% | 8.9% | 18.0% |
| MV | 0–2 m | 46.6% | 58.0% | 71.4% | 80.2% | 87.7% |
| | 3–5 m | 9.2% | 11.7% | 14.2% | 16.6% | 20.1% |
| | 6–11 m | 0.0% | 0.4% | 3.1% | 5.1% | 7.1% |
| | 1–4 y | 0.0% | 0.0% | 1.7% | 5.0% | 8.8% |
| | 5–64 y | 0.0% | 0.0% | 0.6% | 1.9% | 3.7% |
| | 65+ y | 0.0% | 0.0% | 5.1% | 18.8% | 29.6% |
| MV + CU | 0–2 m | 41.8% | 49.2% | 56.1% | 60.8% | 65.1% |
| | 3–5 m | 18.7% | 22.3% | 25.5% | 27.9% | 31.2% |
| | 6–11 m | 5.9% | 7.5% | 8.6% | 9.7% | 11.4% |
| | 1–4 y | 0.0% | 0.0% | 2.2% | 4.1% | 6.9% |
| | 5–64 y | 0.0% | 0.0% | 0.6% | 1.5% | 2.4% |
| | 65+ y | 0.0% | 0.0% | 5.9% | 13.7% | 23.7% |

### S7 Country-level doses per averted hospitalisation

This section reports the country-level number of doses required to avert one hospitalisation in infants under one year of age (doses per averted hospitalisation, DPA), complementing the aggregate results presented in the main text (Figure 7). For each combination of immunisation strategy (long-acting monoclonal antibodies, maternal vaccination, and the two with seasonal catch-up) and coverage level (25%, 50%, 75%, and 95%), we report the 5th, 25th, 50th (median), 75th, and 95th percentiles of the simulation-level distribution across the 300 retained simulations.

DPA is computed at the simulation level as the ratio between the total doses administered for country $c$ under scenario $S$ and the corresponding number of averted hospitalisations in the first year of life; simulations in which the intervention did not avert at least one hospitalisation are discarded from the calculation, as DPA is undefined in those cases. The fraction of discarded simulations is small in most country–scenario combinations and grows for low-coverage scenarios in countries with smaller infant populations, where stochastic variability is more pronounced.

Tables are organised by strategy and, within each strategy, by decreasing coverage level. Countries are listed alphabetically.

#### S7.1 Long-acting monoclonal antibodies

Table S79: **Country-level doses per averted hospitalisation in the first year of life – la-mAbs, 95% coverage.** For each country, we report the 5th, 25th, 50th (median), 75th, and 95th percentiles of the simulation-level distribution across the 300 retained simulations.

| Country | Q05 | Q25 | Median | Q75 | Q95 |
|---|---|---|---|---|---|
| Austria | 35 | 47 | 55 | 65 | 84 |
| Belgium | 35 | 44 | 52 | 62 | 79 |
| Bulgaria | 30 | 39 | 45 | 55 | 79 |
| Croatia | 25 | 34 | 44 | 57 | 106 |
| Cyprus | 26 | 39 | 55 | 73 | 173 |
| Czechia | 31 | 38 | 44 | 54 | 74 |
| Denmark | 36 | 45 | 55 | 67 | 102 |
| Estonia | 24 | 38 | 53 | 72 | 129 |
| Finland | 30 | 40 | 47 | 59 | 86 |
| France | 30 | 34 | 38 | 43 | 52 |
| Germany | 35 | 41 | 47 | 55 | 64 |
| Greece | 30 | 38 | 44 | 52 | 66 |
| Hungary | 30 | 38 | 44 | 51 | 68 |
| Ireland | 30 | 39 | 48 | 58 | 96 |
| Italy | 33 | 39 | 44 | 51 | 63 |
| Latvia | 24 | 37 | 46 | 63 | 113 |
| Lithuania | 25 | 38 | 47 | 59 | 96 |
| Luxembourg | 30 | 45 | 60 | 94 | 213 |
| Malta | 32 | 49 | 68 | 100 | 228 |
| Netherlands | 54 | 68 | 77 | 91 | 118 |
| Norway | 43 | 55 | 65 | 78 | 99 |
| Poland | 32 | 42 | 51 | 62 | 92 |
| Portugal | 38 | 51 | 60 | 71 | 94 |
| Romania | 33 | 41 | 47 | 58 | 71 |
| Slovakia | 30 | 38 | 45 | 57 | 85 |
| Slovenia | 27 | 36 | 45 | 57 | 109 |
| Spain | 39 | 46 | 54 | 62 | 77 |
| Sweden | 35 | 45 | 55 | 65 | 91 |

Table S80: **Country-level doses per averted hospitalisation in the first year of life – la-mAbs, 75% coverage.** For each country, we report the 5th, 25th, 50th (median), 75th, and 95th percentiles of the simulation-level distribution across the 300 retained simulations.

| Country | Q05 | Q25 | Median | Q75 | Q95 |
|---|---|---|---|---|---|
| Austria | 35 | 46 | 55 | 66 | 95 |
| Belgium | 34 | 45 | 52 | 63 | 83 |
| Bulgaria | 28 | 37 | 45 | 57 | 92 |
| Croatia | 22 | 33 | 44 | 59 | 150 |
| Cyprus | 22 | 38 | 53 | 79 | 196 |
| Czechia | 28 | 38 | 45 | 55 | 83 |
| Denmark | 34 | 46 | 54 | 68 | 105 |
| Estonia | 23 | 36 | 56 | 81 | 154 |
| Finland | 27 | 38 | 47 | 63 | 100 |
| France | 30 | 34 | 39 | 44 | 53 |
| Germany | 35 | 41 | 48 | 55 | 67 |
| Greece | 28 | 37 | 44 | 52 | 72 |
| Hungary | 29 | 37 | 46 | 53 | 74 |
| Ireland | 27 | 38 | 48 | 60 | 91 |
| Italy | 33 | 39 | 44 | 51 | 65 |
| Latvia | 21 | 35 | 48 | 69 | 191 |
| Lithuania | 26 | 36 | 48 | 63 | 126 |
| Luxembourg | 28 | 41 | 55 | 90 | 207 |
| Malta | 31 | 45 | 66 | 117 | 331 |
| Netherlands | 50 | 67 | 79 | 92 | 128 |
| Norway | 41 | 55 | 65 | 79 | 112 |
| Poland | 32 | 41 | 51 | 63 | 98 |
| Portugal | 38 | 50 | 60 | 71 | 96 |
| Romania | 32 | 40 | 48 | 60 | 83 |
| Slovakia | 26 | 36 | 45 | 59 | 97 |
| Slovenia | 25 | 35 | 46 | 61 | 118 |
| Spain | 38 | 46 | 55 | 63 | 79 |
| Sweden | 36 | 46 | 56 | 67 | 100 |

Table S81: **Country-level doses per averted hospitalisation in the first year of life – la-mAbs, 50% coverage.** For each country, we report the 5th, 25th, 50th (median), 75th, and 95th percentiles of the simulation-level distribution across the 300 retained simulations.

| Country | Q05 | Q25 | Median | Q75 | Q95 |
|---|---|---|---|---|---|
| Austria | 30 | 44 | 58 | 73 | 121 |
| Belgium | 33 | 44 | 53 | 66 | 89 |
| Bulgaria | 24 | 35 | 46 | 64 | 127 |
| Croatia | 17 | 29 | 40 | 64 | 200 |
| Cyprus | 18 | 31 | 47 | 80 | 221 |
| Czechia | 24 | 36 | 45 | 61 | 104 |
| Denmark | 29 | 42 | 55 | 74 | 142 |
| Estonia | 18 | 34 | 47 | 77 | 248 |
| Finland | 24 | 34 | 48 | 67 | 135 |
| France | 28 | 33 | 39 | 46 | 56 |
| Germany | 33 | 41 | 47 | 54 | 68 |
| Greece | 25 | 35 | 45 | 56 | 84 |
| Hungary | 24 | 35 | 44 | 56 | 91 |
| Ireland | 23 | 35 | 47 | 64 | 148 |
| Italy | 31 | 39 | 44 | 51 | 67 |
| Latvia | 16 | 29 | 42 | 64 | 175 |
| Lithuania | 20 | 33 | 47 | 71 | 157 |
| Luxembourg | 22 | 37 | 55 | 91 | 260 |
| Malta | 23 | 40 | 69 | 120 | 330 |
| Netherlands | 49 | 66 | 80 | 98 | 137 |
| Norway | 35 | 52 | 68 | 87 | 165 |
| Poland | 29 | 42 | 52 | 66 | 99 |
| Portugal | 35 | 48 | 59 | 76 | 109 |
| Romania | 27 | 38 | 48 | 65 | 96 |
| Slovakia | 24 | 35 | 45 | 61 | 131 |
| Slovenia | 22 | 32 | 45 | 66 | 207 |
| Spain | 36 | 47 | 55 | 64 | 83 |
| Sweden | 27 | 42 | 52 | 72 | 179 |

Table S82: **Country-level doses per averted hospitalisation in the first year of life – la-mAbs, 25% coverage.** For each country, we report the 5th, 25th, 50th (median), 75th, and 95th percentiles of the simulation-level distribution across the 300 retained simulations.

| **Country** | **Q05** | **Q25** | **Median** | **Q75** | **Q95** |
|---|---|---|---|---|---|
| Austria | 20 | 40 | 54 | 85 | 253 |
| Belgium | 27 | 40 | 55 | 71 | 163 |
| Bulgaria | 17 | 29 | 42 | 68 | 194 |
| Croatia | 11 | 21 | 32 | 56 | 223 |
| Cyprus | 10 | 24 | 41 | 72 | 240 |
| Czechia | 19 | 30 | 42 | 64 | 162 |
| Denmark | 22 | 35 | 51 | 82 | 242 |
| Estonia | 10 | 23 | 39 | 71 | 374 |
| Finland | 14 | 27 | 42 | 71 | 202 |
| France | 22 | 32 | 40 | 49 | 74 |
| Germany | 28 | 38 | 47 | 58 | 88 |
| Greece | 18 | 31 | 42 | 60 | 167 |
| Hungary | 18 | 28 | 40 | 67 | 206 |
| Ireland | 14 | 27 | 41 | 71 | 243 |
| Italy | 28 | 36 | 44 | 58 | 95 |
| Latvia | 9 | 21 | 32 | 55 | 231 |
| Lithuania | 13 | 23 | 36 | 70 | 305 |
| Luxembourg | 12 | 23 | 40 | 83 | 456 |
| Malta | 14 | 27 | 46 | 93 | 602 |
| Netherlands | 40 | 61 | 79 | 116 | 310 |
| Norway | 27 | 44 | 60 | 110 | 387 |
| Poland | 20 | 39 | 53 | 72 | 144 |
| Portugal | 27 | 44 | 63 | 90 | 272 |
| Romania | 20 | 33 | 46 | 66 | 157 |
| Slovakia | 17 | 26 | 40 | 68 | 201 |
| Slovenia | 12 | 23 | 35 | 61 | 263 |
| Spain | 30 | 42 | 55 | 69 | 118 |
| Sweden | 21 | 34 | 51 | 79 | 240 |

#### S7.2 Long-acting monoclonal antibodies with seasonal catch-up

Table S83: **Country-level doses per averted hospitalisation in the first year of life – la-mAbs with catch-up,** 95% **coverage.** For each country, we report the 5th, 25th, 50th (median), 75th, and 95th percentiles of the simulation-level distribution across the 300 retained simulations.

| Country | Q05 | Q25 | Median | Q75 | Q95 |
|---|---|---|---|---|---|
| Austria | 43 | 53 | 62 | 70 | 87 |
| Belgium | 43 | 52 | 61 | 69 | 89 |
| Bulgaria | 35 | 42 | 49 | 56 | 68 |
| Croatia | 31 | 40 | 49 | 59 | 81 |
| Cyprus | 35 | 49 | 61 | 73 | 115 |
| Czechia | 37 | 45 | 50 | 57 | 71 |
| Denmark | 41 | 51 | 57 | 67 | 86 |
| Estonia | 34 | 48 | 60 | 76 | 102 |
| Finland | 34 | 44 | 51 | 60 | 78 |
| France | 33 | 37 | 41 | 46 | 54 |
| Germany | 40 | 46 | 51 | 60 | 71 |
| Greece | 35 | 42 | 48 | 56 | 68 |
| Hungary | 36 | 42 | 47 | 54 | 65 |
| Ireland | 35 | 43 | 49 | 56 | 66 |
| Italy | 38 | 44 | 49 | 55 | 66 |
| Latvia | 34 | 45 | 55 | 68 | 98 |
| Lithuania | 34 | 44 | 52 | 63 | 83 |
| Luxembourg | 39 | 54 | 68 | 93 | 157 |
| Malta | 46 | 68 | 89 | 124 | 234 |
| Netherlands | 64 | 77 | 87 | 101 | 133 |
| Norway | 48 | 61 | 68 | 79 | 103 |
| Poland | 40 | 49 | 59 | 73 | 108 |
| Portugal | 46 | 57 | 66 | 78 | 96 |
| Romania | 40 | 46 | 51 | 61 | 71 |
| Slovakia | 36 | 43 | 50 | 58 | 73 |
| Slovenia | 35 | 43 | 51 | 61 | 80 |
| Spain | 45 | 52 | 60 | 68 | 84 |
| Sweden | 42 | 51 | 57 | 65 | 79 |

Table S84: **Country-level doses per averted hospitalisation in the first year of life – la-mAbs with catch-up, 75% coverage.** For each country, we report the 5th, 25th, 50th (median), 75th, and 95th percentiles of the simulation-level distribution across the 300 retained simulations.

| **Country** | **Q05** | **Q25** | **Median** | **Q75** | **Q95** |
|---|---|---|---|---|---|
| Austria | 42 | 54 | 62 | 70 | 91 |
| Belgium | 41 | 52 | 60 | 70 | 89 |
| Bulgaria | 35 | 43 | 49 | 57 | 74 |
| Croatia | 29 | 39 | 50 | 59 | 95 |
| Cyprus | 31 | 47 | 59 | 78 | 122 |
| Czechia | 37 | 44 | 51 | 59 | 73 |
| Denmark | 41 | 50 | 57 | 68 | 87 |
| Estonia | 33 | 45 | 61 | 76 | 126 |
| Finland | 35 | 44 | 52 | 62 | 79 |
| France | 33 | 37 | 40 | 46 | 55 |
| Germany | 39 | 46 | 51 | 60 | 72 |
| Greece | 35 | 42 | 48 | 57 | 72 |
| Hungary | 35 | 42 | 48 | 55 | 66 |
| Ireland | 33 | 42 | 49 | 57 | 70 |
| Italy | 38 | 44 | 49 | 56 | 68 |
| Latvia | 32 | 43 | 54 | 66 | 110 |
| Lithuania | 32 | 43 | 53 | 65 | 87 |
| Luxembourg | 35 | 53 | 72 | 100 | 197 |
| Malta | 44 | 65 | 88 | 122 | 252 |
| Netherlands | 63 | 76 | 87 | 101 | 135 |
| Norway | 46 | 59 | 68 | 80 | 102 |
| Poland | 40 | 49 | 59 | 74 | 107 |
| Portugal | 45 | 57 | 67 | 79 | 97 |
| Romania | 39 | 46 | 52 | 60 | 74 |
| Slovakia | 34 | 42 | 50 | 60 | 77 |
| Slovenia | 33 | 43 | 51 | 65 | 91 |
| Spain | 45 | 53 | 61 | 68 | 84 |
| Sweden | 42 | 50 | 57 | 66 | 81 |

Table S85: **Country-level doses per averted hospitalisation in the first year of life – la-mAbs with catch-up, 50% coverage.** For each country, we report the 5th, 25th, 50th (median), 75th, and 95th percentiles of the simulation-level distribution across the 300 retained simulations.

| **Country** | **Q05** | **Q25** | **Median** | **Q75** | **Q95** |
|---|---|---|---|---|---|
| Austria | 40 | 53 | 61 | 71 | 98 |
| Belgium | 40 | 51 | 61 | 71 | 91 |
| Bulgaria | 31 | 40 | 49 | 60 | 90 |
| Croatia | 25 | 38 | 49 | 65 | 150 |
| Cyprus | 25 | 42 | 56 | 83 | 199 |
| Czechia | 31 | 44 | 51 | 61 | 81 |
| Denmark | 37 | 49 | 58 | 71 | 96 |
| Estonia | 25 | 44 | 61 | 87 | 170 |
| Finland | 31 | 42 | 53 | 65 | 95 |
| France | 32 | 37 | 41 | 47 | 56 |
| Germany | 39 | 45 | 51 | 60 | 73 |
| Greece | 31 | 41 | 49 | 56 | 70 |
| Hungary | 32 | 41 | 49 | 58 | 73 |
| Ireland | 29 | 40 | 48 | 60 | 92 |
| Italy | 37 | 44 | 49 | 56 | 67 |
| Latvia | 25 | 42 | 54 | 73 | 145 |
| Lithuania | 29 | 39 | 52 | 70 | 120 |
| Luxembourg | 28 | 49 | 71 | 100 | 263 |
| Malta | 38 | 56 | 81 | 130 | 395 |
| Netherlands | 57 | 75 | 89 | 103 | 139 |
| Norway | 46 | 60 | 71 | 85 | 111 |
| Poland | 39 | 48 | 59 | 75 | 111 |
| Portugal | 42 | 55 | 66 | 80 | 102 |
| Romania | 35 | 46 | 52 | 63 | 82 |
| Slovakia | 31 | 41 | 50 | 64 | 94 |
| Slovenia | 29 | 40 | 50 | 69 | 109 |
| Spain | 42 | 52 | 61 | 69 | 85 |
| Sweden | 39 | 49 | 58 | 68 | 93 |

Table S86: **Country-level doses per averted hospitalisation in the first year of life – la-mAbs with catch-up, 25% coverage.** For each country, we report the 5th, 25th, 50th (median), 75th, and 95th percentiles of the simulation-level distribution across the 300 retained simulations.

| Country | Q05 | Q25 | Median | Q75 | Q95 |
|---|---|---|---|---|---|
| Austria | 30 | 49 | 61 | 79 | 142 |
| Belgium | 37 | 50 | 61 | 74 | 102 |
| Bulgaria | 24 | 36 | 48 | 65 | 157 |
| Croatia | 18 | 30 | 43 | 67 | 235 |
| Cyprus | 15 | 36 | 55 | 93 | 269 |
| Czechia | 26 | 40 | 52 | 64 | 118 |
| Denmark | 30 | 46 | 60 | 77 | 128 |
| Estonia | 18 | 34 | 56 | 92 | 433 |
| Finland | 26 | 38 | 53 | 75 | 155 |
| France | 28 | 36 | 43 | 50 | 62 |
| Germany | 37 | 44 | 53 | 61 | 79 |
| Greece | 22 | 38 | 50 | 65 | 121 |
| Hungary | 26 | 37 | 49 | 62 | 116 |
| Ireland | 23 | 35 | 49 | 70 | 195 |
| Italy | 34 | 41 | 50 | 58 | 78 |
| Latvia | 17 | 30 | 49 | 76 | 209 |
| Lithuania | 23 | 35 | 53 | 84 | 265 |
| Luxembourg | 19 | 36 | 55 | 104 | 427 |
| Malta | 22 | 40 | 68 | 129 | 291 |
| Netherlands | 50 | 71 | 90 | 111 | 170 |
| Norway | 37 | 56 | 73 | 95 | 157 |
| Poland | 31 | 49 | 62 | 79 | 131 |
| Portugal | 37 | 54 | 68 | 91 | 139 |
| Romania | 29 | 43 | 53 | 70 | 127 |
| Slovakia | 23 | 37 | 52 | 76 | 163 |
| Slovenia | 19 | 35 | 49 | 70 | 196 |
| Spain | 40 | 52 | 63 | 73 | 104 |
| Sweden | 27 | 46 | 61 | 79 | 179 |

#### S7.3 Maternal vaccination

Table S87: **Country-level doses per averted hospitalisation in the first year of life – Maternal vaccination, 95% coverage.** For each country, we report the 5th, 25th, 50th (median), 75th, and 95th percentiles of the simulation-level distribution across the 300 retained simulations.

| Country | Q05 | Q25 | Median | Q75 | Q95 |
|---|---|---|---|---|---|
| Austria | 33 | 47 | 58 | 73 | 125 |
| Belgium | 35 | 47 | 56 | 72 | 105 |
| Bulgaria | 28 | 36 | 47 | 64 | 117 |
| Croatia | 21 | 32 | 44 | 61 | 128 |
| Cyprus | 19 | 34 | 50 | 78 | 182 |
| Czechia | 28 | 39 | 47 | 61 | 90 |
| Denmark | 35 | 45 | 57 | 75 | 117 |
| Estonia | 23 | 35 | 53 | 82 | 219 |
| Finland | 25 | 38 | 49 | 63 | 115 |
| France | 28 | 35 | 41 | 48 | 66 |
| Germany | 33 | 42 | 50 | 61 | 84 |
| Greece | 26 | 37 | 47 | 59 | 92 |
| Hungary | 27 | 37 | 46 | 57 | 82 |
| Ireland | 28 | 39 | 48 | 65 | 108 |
| Italy | 32 | 40 | 47 | 57 | 77 |
| Latvia | 20 | 33 | 46 | 71 | 229 |
| Lithuania | 23 | 35 | 49 | 69 | 146 |
| Luxembourg | 26 | 44 | 61 | 102 | 178 |
| Malta | 29 | 50 | 77 | 134 | 488 |
| Netherlands | 53 | 67 | 81 | 102 | 158 |
| Norway | 40 | 55 | 69 | 91 | 135 |
| Poland | 30 | 41 | 54 | 73 | 113 |
| Portugal | 38 | 52 | 65 | 80 | 119 |
| Romania | 31 | 40 | 48 | 61 | 94 |
| Slovakia | 26 | 37 | 48 | 66 | 123 |
| Slovenia | 24 | 34 | 48 | 69 | 140 |
| Spain | 38 | 48 | 60 | 71 | 97 |
| Sweden | 33 | 45 | 58 | 76 | 114 |

Table S88: **Country-level doses per averted hospitalisation in the first year of life – Maternal vaccination, 75% coverage.** For each country, we report the 5th, 25th, 50th (median), 75th, and 95th percentiles of the simulation-level distribution across the 300 retained simulations.

| **Country** | **Q05** | **Q25** | **Median** | **Q75** | **Q95** |
|---|---|---|---|---|---|
| Austria | 31 | 46 | 58 | 73 | 122 |
| Belgium | 34 | 47 | 58 | 73 | 123 |
| Bulgaria | 26 | 35 | 47 | 65 | 152 |
| Croatia | 20 | 31 | 44 | 71 | 239 |
| Cyprus | 18 | 32 | 46 | 78 | 258 |
| Czechia | 26 | 38 | 49 | 67 | 126 |
| Denmark | 30 | 43 | 58 | 80 | 127 |
| Estonia | 19 | 36 | 52 | 84 | 299 |
| Finland | 25 | 36 | 49 | 72 | 154 |
| France | 27 | 34 | 41 | 50 | 67 |
| Germany | 34 | 42 | 50 | 61 | 85 |
| Greece | 28 | 37 | 48 | 62 | 93 |
| Hungary | 26 | 37 | 47 | 59 | 94 |
| Ireland | 26 | 38 | 51 | 70 | 112 |
| Italy | 32 | 39 | 48 | 58 | 83 |
| Latvia | 17 | 32 | 46 | 68 | 179 |
| Lithuania | 23 | 35 | 49 | 74 | 155 |
| Luxembourg | 22 | 39 | 61 | 106 | 236 |
| Malta | 27 | 44 | 66 | 114 | 318 |
| Netherlands | 51 | 68 | 80 | 101 | 156 |
| Norway | 36 | 57 | 70 | 92 | 175 |
| Poland | 30 | 42 | 55 | 76 | 121 |
| Portugal | 35 | 50 | 63 | 83 | 125 |
| Romania | 28 | 39 | 48 | 62 | 98 |
| Slovakia | 24 | 34 | 46 | 68 | 150 |
| Slovenia | 22 | 33 | 45 | 72 | 170 |
| Spain | 37 | 49 | 61 | 74 | 98 |
| Sweden | 31 | 44 | 57 | 78 | 143 |

Table S89: **Country-level doses per averted hospitalisation in the first year of life – Maternal vaccination,** 50% **coverage.** For each country, we report the 5th, 25th, 50th (median), 75th, and 95th percentiles of the simulation-level distribution across the 300 retained simulations.

| **Country** | **Q05** | **Q25** | **Median** | **Q75** | **Q95** |
|---|---|---|---|---|---|
| Austria | 28 | 42 | 57 | 80 | 188 |
| Belgium | 33 | 46 | 59 | 78 | 145 |
| Bulgaria | 21 | 32 | 45 | 72 | 238 |
| Croatia | 16 | 26 | 40 | 72 | 243 |
| Cyprus | 13 | 25 | 43 | 90 | 561 |
| Czechia | 22 | 35 | 46 | 64 | 123 |
| Denmark | 30 | 44 | 58 | 86 | 193 |
| Estonia | 13 | 29 | 47 | 78 | 318 |
| Finland | 21 | 35 | 49 | 77 | 310 |
| France | 27 | 34 | 40 | 50 | 74 |
| Germany | 31 | 40 | 49 | 63 | 94 |
| Greece | 22 | 35 | 49 | 66 | 153 |
| Hungary | 21 | 32 | 44 | 61 | 130 |
| Ireland | 20 | 32 | 45 | 67 | 178 |
| Italy | 28 | 38 | 47 | 57 | 88 |
| Latvia | 15 | 25 | 38 | 59 | 201 |
| Lithuania | 16 | 30 | 43 | 80 | 328 |
| Luxembourg | 17 | 31 | 49 | 86 | 413 |
| Malta | 22 | 38 | 59 | 117 | 351 |
| Netherlands | 45 | 65 | 83 | 108 | 220 |
| Norway | 33 | 52 | 73 | 100 | 196 |
| Poland | 24 | 43 | 55 | 76 | 133 |
| Portugal | 33 | 47 | 66 | 90 | 154 |
| Romania | 26 | 36 | 49 | 65 | 156 |
| Slovakia | 19 | 33 | 47 | 73 | 230 |
| Slovenia | 17 | 29 | 42 | 71 | 228 |
| Spain | 35 | 49 | 61 | 76 | 120 |
| Sweden | 27 | 39 | 56 | 83 | 251 |

Table S90: **Country-level doses per averted hospitalisation in the first year of life – Maternal vaccination, 25% coverage.** For each country, we report the 5th, 25th, 50th (median), 75th, and 95th percentiles of the simulation-level distribution across the 300 retained simulations.

| Country | Q05 | Q25 | Median | Q75 | Q95 |
|---|---|---|---|---|---|
| Austria | 19 | 34 | 52 | 79 | 269 |
| Belgium | 26 | 40 | 57 | 86 | 214 |
| Bulgaria | 15 | 24 | 38 | 69 | 318 |
| Croatia | 9 | 17 | 25 | 51 | 247 |
| Cyprus | 7 | 15 | 24 | 50 | 186 |
| Czechia | 14 | 30 | 43 | 71 | 210 |
| Denmark | 17 | 31 | 52 | 92 | 296 |
| Estonia | 7 | 19 | 34 | 69 | 354 |
| Finland | 12 | 23 | 38 | 69 | 277 |
| France | 20 | 31 | 41 | 59 | 115 |
| Germany | 27 | 36 | 49 | 69 | 131 |
| Greece | 15 | 29 | 43 | 71 | 249 |
| Hungary | 16 | 26 | 41 | 70 | 243 |
| Ireland | 12 | 22 | 36 | 65 | 289 |
| Italy | 26 | 36 | 49 | 67 | 142 |
| Latvia | 8 | 16 | 28 | 58 | 167 |
| Lithuania | 10 | 21 | 33 | 68 | 433 |
| Luxembourg | 9 | 18 | 36 | 66 | 241 |
| Malta | 12 | 22 | 37 | 58 | 174 |
| Netherlands | 33 | 59 | 78 | 126 | 399 |
| Norway | 22 | 42 | 64 | 109 | 446 |
| Poland | 19 | 37 | 54 | 87 | 235 |
| Portugal | 24 | 41 | 62 | 96 | 354 |
| Romania | 16 | 30 | 42 | 70 | 264 |
| Slovakia | 14 | 24 | 39 | 79 | 284 |
| Slovenia | 9 | 20 | 36 | 70 | 317 |
| Spain | 31 | 44 | 59 | 83 | 182 |
| Sweden | 17 | 30 | 45 | 87 | 423 |

#### S7.4 Maternal vaccination with seasonal catch-up

Table S91: **Country-level doses per averted hospitalisation in the first year of life – Maternal vaccination with catch-up,** 95% **coverage.** For each country, we report the 5th, 25th, 50th (median), 75th, and 95th percentiles of the simulation-level distribution across the 300 retained simulations.

| **Country** | **Q05** | **Q25** | **Median** | **Q75** | **Q95** |
|---|---|---|---|---|---|
| Austria | 43 | 55 | 65 | 76 | 96 |
| Belgium | 43 | 55 | 65 | 76 | 96 |
| Bulgaria | 34 | 43 | 51 | 59 | 77 |
| Croatia | 32 | 42 | 50 | 64 | 90 |
| Cyprus | 33 | 50 | 63 | 83 | 135 |
| Czechia | 37 | 45 | 52 | 60 | 77 |
| Denmark | 41 | 50 | 59 | 70 | 89 |
| Estonia | 32 | 49 | 65 | 83 | 137 |
| Finland | 35 | 46 | 54 | 65 | 86 |
| France | 33 | 38 | 42 | 48 | 59 |
| Germany | 40 | 47 | 54 | 63 | 78 |
| Greece | 35 | 44 | 51 | 59 | 76 |
| Hungary | 35 | 42 | 49 | 56 | 70 |
| Ireland | 35 | 43 | 51 | 59 | 74 |
| Italy | 38 | 45 | 52 | 59 | 75 |
| Latvia | 32 | 45 | 58 | 71 | 113 |
| Lithuania | 32 | 45 | 56 | 68 | 94 |
| Luxembourg | 36 | 54 | 72 | 97 | 197 |
| Malta | 47 | 69 | 99 | 130 | 276 |
| Netherlands | 63 | 79 | 91 | 107 | 141 |
| Norway | 48 | 61 | 72 | 85 | 111 |
| Poland | 41 | 51 | 62 | 77 | 114 |
| Portugal | 45 | 58 | 71 | 85 | 109 |
| Romania | 40 | 47 | 54 | 62 | 81 |
| Slovakia | 36 | 44 | 52 | 63 | 85 |
| Slovenia | 34 | 45 | 54 | 65 | 91 |
| Spain | 45 | 54 | 64 | 74 | 92 |
| Sweden | 42 | 51 | 58 | 68 | 83 |

Table S92: **Country-level doses per averted hospitalisation in the first year of life – Maternal vaccination with catch-up, 75% coverage.** For each country, we report the 5th, 25th, 50th (median), 75th, and 95th percentiles of the simulation-level distribution across the 300 retained simulations.

| **Country** | **Q05** | **Q25** | **Median** | **Q75** | **Q95** |
|---|---|---|---|---|---|
| Austria | 40 | 55 | 65 | 77 | 99 |
| Belgium | 43 | 55 | 65 | 78 | 100 |
| Bulgaria | 32 | 41 | 50 | 61 | 79 |
| Croatia | 29 | 41 | 52 | 64 | 104 |
| Cyprus | 30 | 46 | 61 | 83 | 138 |
| Czechia | 36 | 45 | 52 | 62 | 83 |
| Denmark | 41 | 50 | 58 | 72 | 95 |
| Estonia | 31 | 46 | 62 | 85 | 167 |
| Finland | 34 | 45 | 55 | 67 | 93 |
| France | 33 | 38 | 42 | 48 | 60 |
| Germany | 40 | 47 | 53 | 64 | 79 |
| Greece | 34 | 43 | 51 | 60 | 79 |
| Hungary | 34 | 41 | 49 | 58 | 71 |
| Ireland | 32 | 42 | 50 | 60 | 79 |
| Italy | 39 | 45 | 52 | 59 | 73 |
| Latvia | 28 | 44 | 57 | 73 | 134 |
| Lithuania | 32 | 44 | 55 | 68 | 106 |
| Luxembourg | 33 | 54 | 72 | 100 | 200 |
| Malta | 43 | 67 | 101 | 156 | 419 |
| Netherlands | 63 | 78 | 91 | 108 | 143 |
| Norway | 48 | 61 | 73 | 85 | 114 |
| Poland | 39 | 50 | 62 | 78 | 119 |
| Portugal | 44 | 58 | 70 | 86 | 108 |
| Romania | 38 | 46 | 53 | 63 | 80 |
| Slovakia | 34 | 43 | 52 | 63 | 92 |
| Slovenia | 32 | 43 | 51 | 66 | 92 |
| Spain | 45 | 55 | 65 | 74 | 95 |
| Sweden | 40 | 51 | 59 | 69 | 88 |

Table S93: **Country-level doses per averted hospitalisation in the first year of life – Maternal vaccination with catch-up, 50% coverage.** For each country, we report the 5th, 25th, 50th (median), 75th, and 95th percentiles of the simulation-level distribution across the 300 retained simulations.

| Country | Q05 | Q25 | Median | Q75 | Q95 |
|---|---|---|---|---|---|
| Austria | 38 | 54 | 65 | 77 | 109 |
| Belgium | 42 | 55 | 66 | 78 | 107 |
| Bulgaria | 30 | 40 | 52 | 65 | 95 |
| Croatia | 26 | 36 | 50 | 73 | 163 |
| Cyprus | 24 | 43 | 61 | 95 | 338 |
| Czechia | 32 | 44 | 53 | 62 | 86 |
| Denmark | 36 | 47 | 58 | 76 | 109 |
| Estonia | 26 | 42 | 60 | 97 | 246 |
| Finland | 30 | 43 | 54 | 68 | 108 |
| France | 31 | 38 | 43 | 50 | 62 |
| Germany | 38 | 46 | 54 | 64 | 81 |
| Greece | 31 | 42 | 52 | 63 | 87 |
| Hungary | 32 | 41 | 49 | 60 | 83 |
| Ireland | 29 | 41 | 49 | 63 | 91 |
| Italy | 37 | 45 | 52 | 59 | 74 |
| Latvia | 23 | 38 | 55 | 78 | 166 |
| Lithuania | 28 | 43 | 57 | 77 | 156 |
| Luxembourg | 27 | 48 | 66 | 117 | 276 |
| Malta | 36 | 61 | 87 | 149 | 515 |
| Netherlands | 57 | 77 | 90 | 111 | 148 |
| Norway | 43 | 60 | 73 | 89 | 132 |
| Poland | 35 | 49 | 63 | 80 | 118 |
| Portugal | 43 | 57 | 71 | 88 | 115 |
| Romania | 34 | 45 | 53 | 66 | 88 |
| Slovakia | 30 | 42 | 53 | 68 | 113 |
| Slovenia | 27 | 38 | 51 | 75 | 171 |
| Spain | 43 | 54 | 64 | 74 | 94 |
| Sweden | 37 | 49 | 58 | 73 | 103 |

Table S94: **Country-level doses per averted hospitalisation in the first year of life – Maternal vaccination with catch-up,** 25% **coverage.** For each country, we report the 5th, 25th, 50th (median), 75th, and 95th percentiles of the simulation-level distribution across the 300 retained simulations.

| Country | Q05 | Q25 | Median | Q75 | Q95 |
|---|---|---|---|---|---|
| Austria | 28 | 47 | 65 | 93 | 191 |
| Belgium | 35 | 52 | 65 | 84 | 140 |
| Bulgaria | 22 | 36 | 51 | 76 | 170 |
| Croatia | 16 | 28 | 47 | 78 | 201 |
| Cyprus | 15 | 26 | 48 | 85 | 339 |
| Czechia | 27 | 41 | 52 | 74 | 134 |
| Denmark | 28 | 42 | 58 | 85 | 190 |
| Estonia | 17 | 31 | 57 | 98 | 301 |
| Finland | 23 | 39 | 56 | 86 | 202 |
| France | 29 | 37 | 44 | 53 | 73 |
| Germany | 36 | 46 | 53 | 67 | 87 |
| Greece | 24 | 40 | 54 | 71 | 156 |
| Hungary | 24 | 36 | 50 | 67 | 128 |
| Ireland | 20 | 32 | 43 | 66 | 213 |
| Italy | 34 | 43 | 51 | 61 | 84 |
| Latvia | 14 | 27 | 48 | 79 | 187 |
| Lithuania | 18 | 36 | 54 | 87 | 409 |
| Luxembourg | 16 | 30 | 54 | 95 | 281 |
| Malta | 22 | 38 | 69 | 130 | 518 |
| Netherlands | 49 | 74 | 91 | 115 | 218 |
| Norway | 33 | 54 | 76 | 106 | 266 |
| Poland | 30 | 49 | 64 | 87 | 141 |
| Portugal | 35 | 54 | 74 | 99 | 180 |
| Romania | 28 | 40 | 54 | 72 | 126 |
| Slovakia | 24 | 33 | 48 | 77 | 159 |
| Slovenia | 17 | 30 | 44 | 80 | 314 |
| Spain | 40 | 54 | 65 | 81 | 109 |
| Sweden | 28 | 44 | 59 | 85 | 210 |

### S8 Sensitivity analysis to the waning immunity assumptions

This section provides the full results of the sensitivity analysis on the waning of treatment efficacy, conducted using Italy as an illustrative case study. We re-ran all intervention scenarios under five alternative specifications of the waning process: no waning, exponential decay, and Erlang distributions with 2, 3, and 4 stages. We retained the three-stage Erlang specification as the main analysis, in line with previous evidence supporting it as the most appropriate choice for modelling waning of RSV immunoprophylaxis [4]. For each coverage level, Table S95–S98 reports the percentage of hospitalisations averted in infants under one year of age under each combination of intervention and waning specification, summarised by the 5th, 25th, 50th (median), 75th, and 95th percentiles of the simulation-level distribution across the 300 retained simulations.

Table S95: **Sensitivity analysis to waning immunity assumptions – Italy,** 95% **coverage.** Percentage of hospitalisations averted in infants under one year of age for each combination of intervention strategy and waning specification, summarised by the 5th, 25th, 50th (median), 75th, and 95th percentiles of the simulation-level distribution across the 300 retained simulations. The three-stage Erlang specification corresponds to the main analysis.

| **Intervention** | **Waning specification** | **P5** | **P25** | **Median** | **P75** | **P95** |
|---|---|---|---|---|---|---|
| mAbs | No waning | 26.96% | 31.64% | 34.14% | 36.91% | 40.25% |
| | Exponential | 20.65% | 24.29% | 26.32% | 28.40% | 31.54% |
| | Erlang(2) | 23.44% | 27.38% | 29.83% | 32.38% | 35.44% |
| | *Erlang(3), main analysis* | 24.65% | 29.06% | 31.25% | 33.92% | 37.59% |
| | Erlang(4) | 25.36% | 29.52% | 31.89% | 34.13% | 37.76% |
| mAbs + Catch-up | No waning | 52.03% | 57.87% | 61.67% | 65.61% | 69.96% |
| | Exponential | 35.76% | 39.91% | 42.86% | 45.45% | 49.40% |
| | Erlang(2) | 41.62% | 45.95% | 49.40% | 53.01% | 56.63% |
| | *Erlang(3), main analysis* | 43.91% | 48.81% | 52.25% | 56.01% | 60.35% |
| | Erlang(4) | 44.96% | 50.70% | 53.91% | 57.82% | 60.81% |
| MV | No waning | 17.47% | 21.62% | 25.11% | 28.47% | 33.69% |
| | Exponential | 13.54% | 16.72% | 19.88% | 22.72% | 26.43% |
| | Erlang(2) | 14.31% | 19.12% | 22.29% | 25.35% | 30.29% |
| | *Erlang(3), main analysis* | 15.38% | 20.00% | 23.12% | 26.85% | 31.15% |
| | Erlang(4) | 16.30% | 20.52% | 23.68% | 27.30% | 32.27% |
| MV + Catch-up | No waning | 42.50% | 48.20% | 52.50% | 57.12% | 61.96% |
| | Exponential | 28.95% | 32.92% | 36.00% | 39.39% | 43.17% |
| | Erlang(2) | 32.79% | 38.36% | 41.54% | 45.39% | 49.75% |
| | *Erlang(3), main analysis* | 35.47% | 40.51% | 43.91% | 48.44% | 53.05% |
| | Erlang(4) | 36.35% | 41.83% | 45.42% | 49.78% | 54.54% |

Table S96: **Sensitivity analysis to waning immunity assumptions – Italy, 75% coverage.** Percentage of hospitalisations averted in infants under one year of age for each combination of intervention strategy and waning specification, summarised by the 5th, 25th, 50th (median), 75th, and 95th percentiles of the simulation-level distribution across the 300 retained simulations. The three-stage Erlang specification corresponds to the main analysis.

| **Intervention** | **Waning specification** | **P5** | **P25** | **Median** | **P75** | **P95** |
|---|---|---|---|---|---|---|
| mAbs | No waning | 20.72% | 24.82% | 26.89% | 28.83% | 32.41% |
| | Exponential | 15.72% | 18.88% | 20.60% | 22.55% | 24.84% |
| | Erlang(2) | 18.14% | 21.16% | 23.77% | 25.85% | 28.96% |
| | *Erlang(3), main analysis* | 19.60% | 22.73% | 24.79% | 26.80% | 29.74% |
| | Erlang(4) | 19.89% | 23.21% | 25.33% | 27.39% | 30.34% |
| mAbs + Catch-up | No waning | 41.02% | 45.51% | 48.64% | 51.67% | 55.66% |
| | Exponential | 28.23% | 31.41% | 33.92% | 36.59% | 39.75% |
| | Erlang(2) | 32.80% | 36.48% | 38.81% | 41.72% | 44.94% |
| | *Erlang(3), main analysis* | 34.32% | 38.35% | 41.20% | 44.63% | 47.77% |
| | Erlang(4) | 35.36% | 39.69% | 42.46% | 45.40% | 48.74% |
| MV | No waning | 13.04% | 16.64% | 19.58% | 23.17% | 26.76% |
| | Exponential | 10.15% | 13.37% | 15.37% | 17.90% | 21.69% |
| | Erlang(2) | 11.45% | 14.59% | 17.74% | 20.11% | 24.60% |
| | *Erlang(3), main analysis* | 11.36% | 15.62% | 18.35% | 20.94% | 24.67% |
| | Erlang(4) | 12.10% | 16.04% | 18.92% | 21.63% | 25.16% |
| MV + Catch-up | No waning | 33.29% | 38.17% | 41.34% | 45.07% | 49.27% |
| | Exponential | 22.60% | 26.17% | 28.51% | 31.18% | 35.07% |
| | Erlang(2) | 25.47% | 29.96% | 32.69% | 35.62% | 40.11% |
| | *Erlang(3), main analysis* | 27.72% | 31.80% | 34.52% | 38.22% | 42.04% |
| | Erlang(4) | 27.73% | 33.07% | 35.98% | 38.89% | 43.04% |

Table S97: **Sensitivity analysis to waning immunity assumptions – Italy, 50% coverage.** Percentage of hospitalisations averted in infants under one year of age for each combination of intervention strategy and waning specification, summarised by the 5th, 25th, 50th (median), 75th, and 95th percentiles of the simulation-level distribution across the 300 retained simulations. The three-stage Erlang specification corresponds to the main analysis.

| **Intervention** | **Waning specification** | **P5** | **P25** | **Median** | **P75** | **P95** |
|---|---|---|---|---|---|---|
| mAbs | No waning | 12.82% | 15.88% | 18.02% | 19.95% | 22.35% |
| | Exponential | 9.64% | 12.11% | 13.71% | 15.45% | 17.87% |
| | Erlang(2) | 11.07% | 13.78% | 15.74% | 17.54% | 20.32% |
| | *Erlang(3), main analysis* | 12.35% | 14.65% | 16.29% | 18.43% | 20.99% |
| | Erlang(4) | 11.95% | 15.11% | 16.86% | 18.66% | 21.23% |
| mAbs + Catch-up | No waning | 26.75% | 30.13% | 32.22% | 34.59% | 37.70% |
| | Exponential | 17.69% | 20.48% | 22.57% | 24.39% | 27.21% |
| | Erlang(2) | 20.67% | 23.66% | 25.73% | 28.26% | 30.84% |
| | *Erlang(3), main analysis* | 22.40% | 25.37% | 27.06% | 29.59% | 32.87% |
| | Erlang(4) | 23.23% | 26.26% | 28.51% | 30.87% | 33.77% |
| MV | No waning | 7.58% | 10.77% | 13.06% | 15.65% | 19.14% |
| | Exponential | 5.75% | 8.21% | 10.00% | 12.16% | 15.90% |
| | Erlang(2) | 6.34% | 9.52% | 11.70% | 13.96% | 16.98% |
| | *Erlang(3), main analysis* | 7.02% | 10.39% | 12.27% | 14.70% | 18.49% |
| | Erlang(4) | 6.81% | 10.39% | 12.64% | 14.71% | 17.57% |
| MV + Catch-up | No waning | 21.65% | 25.01% | 27.37% | 30.09% | 34.02% |
| | Exponential | 13.73% | 16.89% | 18.80% | 21.11% | 24.30% |
| | Erlang(2) | 16.24% | 19.45% | 21.41% | 24.08% | 27.30% |
| | *Erlang(3), main analysis* | 17.88% | 20.97% | 23.14% | 25.67% | 29.14% |
| | Erlang(4) | 18.48% | 21.98% | 24.05% | 26.02% | 29.23% |

Table S98: **Sensitivity analysis to waning immunity assumptions – Italy, 25% coverage.** Percentage of hospitalisations averted in infants under one year of age for each combination of intervention strategy and waning specification, summarised by the 5th, 25th, 50th (median), 75th, and 95th percentiles of the simulation-level distribution across the 300 retained simulations. The three-stage Erlang specification corresponds to the main analysis.

| **Intervention** | **Waning specification** | **P5** | **P25** | **Median** | **P75** | **P95** |
|---|---|---|---|---|---|---|
| mAbs | No waning | 4.87% | 7.40% | 8.91% | 10.35% | 12.58% |
| | Exponential | 2.35% | 5.45% | 6.84% | 8.27% | 10.10% |
| | Erlang(2) | 3.60% | 6.44% | 7.96% | 9.77% | 12.57% |
| | *Erlang(3), main analysis* | 4.05% | 6.77% | 8.31% | 9.63% | 12.34% |
| | Erlang(4) | 4.76% | 6.87% | 8.26% | 9.85% | 13.07% |
| mAbs + Catch-up | No waning | 12.43% | 14.40% | 16.09% | 17.74% | 20.69% |
| | Exponential | 7.21% | 9.49% | 11.18% | 12.70% | 15.56% |
| | Erlang(2) | 8.10% | 11.15% | 13.00% | 14.65% | 17.54% |
| | *Erlang(3), main analysis* | 9.37% | 12.13% | 13.72% | 15.68% | 18.58% |
| | Erlang(4) | 10.03% | 12.55% | 14.05% | 15.61% | 18.33% |
| MV | No waning | 2.82% | 4.80% | 6.55% | 8.16% | 11.36% |
| | Exponential | 1.14% | 3.69% | 5.23% | 6.70% | 9.29% |
| | Erlang(2) | 1.71% | 4.20% | 5.72% | 7.51% | 10.64% |
| | *Erlang(3), main analysis* | 2.16% | 4.21% | 5.96% | 7.64% | 10.19% |
| | Erlang(4) | 2.21% | 4.56% | 6.33% | 7.74% | 11.03% |
| MV + Catch-up | No waning | 9.21% | 12.00% | 13.77% | 15.56% | 18.47% |
| | Exponential | 5.05% | 7.71% | 9.30% | 11.02% | 13.77% |
| | Erlang(2) | 6.32% | 9.05% | 10.97% | 12.89% | 16.47% |
| | *Erlang(3), main analysis* | 7.55% | 10.16% | 11.77% | 13.54% | 16.38% |
| | Erlang(4) | 7.83% | 10.34% | 12.06% | 13.59% | 16.52% |

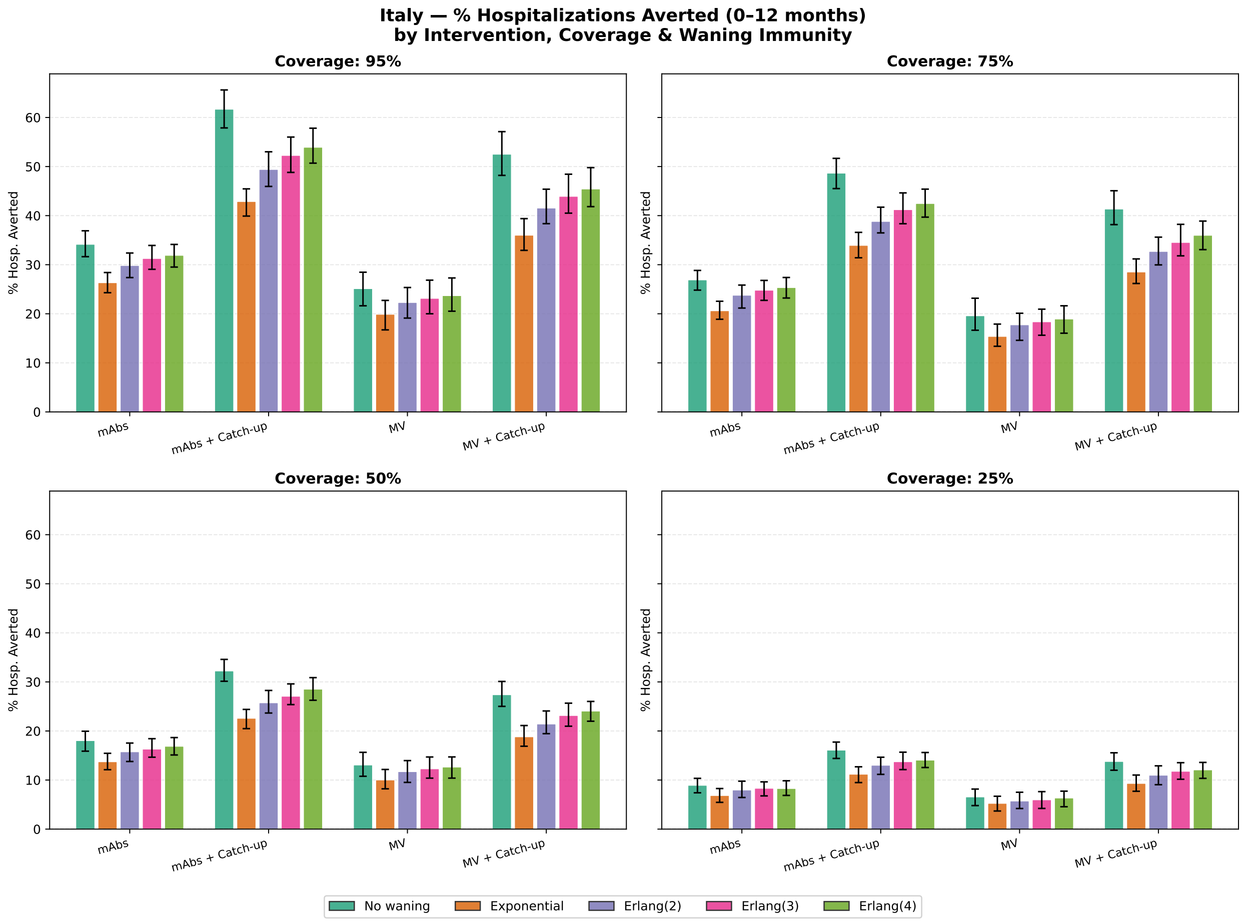

Figure S1: **Sensitivity analysis to waning immunity assumptions – Italy.** Percentage of hospitalisations averted in infants under one year of age, by intervention strategy and waning specification, at each coverage level. Bars show the median across the 300 retained simulations; error bars indicate the interquartile range (25th–75th percentile). Five waning specifications are compared: no waning (green), exponential decay (orange), and Erlang distributions with 2, 3, and 4 stages (purple, pink, and light green, respectively). The three-stage Erlang specification corresponds to the main analysis.

[4] David Hodgson, Neil Wilkins, Edwin van Leeuwen, Conall H Watson, Jonathan Crofts, Stefan Flasche, Mark Jit, and Katherine E Atkins. Protecting infants against rsv disease: an impact and cost-effectiveness comparison of long-acting monoclonal antibodies and maternal vaccination. *The Lancet Regional Health - sEurope*, 38, 2024.
